# Campus Sewage Water Surveillance based dynamics and infection trends of SARS-CoV-2 variants during third wave of COVID-19 in Pune, India

**DOI:** 10.1101/2023.03.02.23286683

**Authors:** Vinita Malik, Vinay Rajput, Rinka Pramanik, Rachel Samson, Rakesh kumar Yadav, Pradnya Kadam, Nikita Shah, Rutuja Sawant, Unnati Bhalerao, Manisha Tupekar, Soumen Khan, Priyanki Shah, LS Shashidhara, Sanjay Kamble, Syed Dastager, Krishanpal Karmodiya, Mahesh Dharne

## Abstract

The wastewater-based epidemiology (WBE) of SARS-CoV-2 is a quick and cost-effective method of tracking virus transmission. However, few studies reported on campus or in academic or residential settings worldwide. In this study, we demonstrated the WBE approach to detect, monitor, and evaluate genomic variants of SARS-CoV-2 fragments in a sewage treatment plant (STP) located on the campus of CSIR National Chemical Laboratory, Pune, India. Herein we describe the early warning capability of WBE, with viral load rise in campus sewage water up to 14 days before its clinical detection. This was supported further by a significant correlation between SARS-CoV-2 RNA concentration and clinically reported COVID-19 cases on campus. Additionally, we comprehended the probable targets missed by the quantitative qRT-PCR using amplicon-based sequencing due to low viral load. The analysis revealed the presence of signature mutations of the Omicron (S:N679K, S:N764K, S:D796Y, N:P13L, ORF1a:T3255I, ORF1a:K856R, ORF1a:P3395H, and N:S413R) before the lineage was first detected globally. Further, we used Lineage decomposition (LCS) tool to detect the Variant of Concern (VOC)/Variant of Interest (VOI) signals upto a month earlier in sewage water samples. The analysis also indicated the transition of lineage from Delta to Omicron in late Decemeber,2021. This is the first study in India highlighting the use of on-campus STP to evaluate the local spread of SARS-CoV-2, which could aid in preventing COVID-19 in academic institutes/universities. This study proves the usefulness of WBE as an early warning system for detecting, tracking and tracing VOCs using the sequencing approach. The current study could aid in taking critical decisions to tackle the pandemic scenario on campus.

**Highlights:**

- The first study on campus sewage water for SARS-CoV-2 surveillance in India
- Early detection of Omicron VOC signals during early November 2021
- Sequencing revealed the presence of Omicron VOC fragments prior to clinical cases reported on campus
- Genomic analysis indicated transition of Delta to Omicron lineage in late December 2021 and potentially led to the third wave
- Combining qRT-PCR and sequencing could be useful for on-campus tracking of variants using wastewater surveillance

## 1. Introduction

Following the detection of the first SARS-CoV-2 infection (Severe Acute Respiratory Syndrome Coronavirus 2) in late December 2019, several variants of this virus have been reported globally. The constantly changing genome of SARS-CoV-2 has challenged healthcare systems worldwide (Zhu et al., 2020). SARS-CoV-2 had infected over 44 million people and killed over 0.5 million people in India as of November 1, 2022 (Worldometer 2022). Despite administrating more than two million doses of the Covid-19 vaccine (https://www.mohfw.gov.in/), the government is still taking action to speed up the immunization campaign to contain the virus. However, the major obstacle for public health is the emergence of new variants of SARS-CoV-2 due to rapid mutations in its genome. The World Health Organization (WHO) has categorized SARS-CoV-2 variants into Variants of Interest (VOIs) and Variants of Concern (VOCs) based on their characteristics and impact on the global community. Monitoring the occurrence of the variants in the population is critical for timely public health interventions. The Alpha (B.1.1.7),B.1.351 (Beta), P.1(Gamma), B.1.617.2 (Delta), and B.1.1.529 (Omicron) are examples of notable VOCs of SARS-CoV-2 (Brief, 2021; Fujino et al., 2021; Galloway et al., 2020; Tegally et al., 2021; WHO, 2021). In October 2020, the Delta variant surfaced in India, resulting in the devastating second COVID-19 outbreak in the summer of 2021. During the Delta wave, India’s average death rate of 3 percent doubled over only three months (Jha et al., 2022). The Omicron variant caused the surge in COVID-19 infection, which was reported first in South Africa in late November 2021(He et al., 2021). Infectiousness and transmission rates for the Omicron variant were more than those of the earlier lineages (Duong et al., 2022; Wolter et al., 2022). Omicron is reportedly highly resistant to antibody-directed inactivation by vaccine-triggered sera and antibodies from both vaccinated and SARS-CoV-2 infected individuals (Hoffman et al., 2022; Tada et al., 2022; Wilhelm et al., 2021).

Primarily, genomic surveillance of SARS-CoV-2 is accomplished through clinical sample sequencing (Grimaldi et al., 2022). It has been observed and widely accepted that sequencing all clinically positive samples is not economically feasible (Tyson et al., 2020). Even though SARS-CoV-2 is primarily a respiratory virus, its genome can also be found in infected patients’ respiratory tract, stool, and urine(Wölfel et al., 2020). Both symptomatic and asymptomatic cases shed the virus in their stool, eventually reaching sewage treatment plants (STPs) (Mizumoto et al., 2020; Nishiura et al., 2020; Treibel et al., 2020; Wan et al., 2020). Based on the above findings, implementing WBE is an inexpensive virus surveillance approach that does not infringe on human rights (Carducci et al., 2020;O’Brien and Xagoraraki, 2019). In the recent past, WBE has been demonstrated to be instrumental during the H1N1 influenza outbreak and in detecting the prevalence of Poliovirus, Norovirus, Hepatitis A and E virus, adenovirus, and rotavirus (Ahmed et al., 2021; Hellmér et al., 2014; Lago et al., 2003). SARS-CoV-2 RNA is detected and quantified in wastewater using quantitative reverse transcription polymerase chain reaction (qRT-PCR) (Hillary et al., 2020). However, the qRT-PCR-based method for detecting variants in SARS-CoV-2 positive samples is only practical for already identified variants and cannot be utilized to identify novel variants (Dikdan et al., 2022; Harper et al., 2021; Shen et al., 2021). Wastewater surveillance is one of the most effective complementary epidemiological methods for detecting the presence and evolution of SARS-CoV-2 variants (Amman et al., 2022). Wastewater has been used to track SARS-CoV-2 quite early in the pandemic (Randazzo et al., 2020; Sherchan et al., 2020). Several researchers have asserted that SARS-CoV-2 can be found in wastewater from all over the world (Markt et al., 2022; Pecia et al., 2022; Wurtz et al., 2021; Wurtzer et al., 2021). WBE allows the detection of mutations before clinical testing is done (Bar-Or et al., 2021; Karthikeyen et al., 2022). The Centers for Disease Control and Prevention (CDC), Atlanta, USA, has also acknowledged the use of environmental wastewater surveillance techniques to track and identify circulating SARS-CoV-2 variants and issued guidelines on its use in the community National Wastewater Surveillance System, CDC, USA).

In this study, we present the applicability of WBE to test the presence of SARS-CoV-2 and its variants in sewage water from a research campus-based STP in India. We collected samples from the STP inlet of the CSIR-National Chemical Laboratory (CSIR-NCL) campus in Pune, India. To determine the SARS-CoV-2 frequency and genetic diversity of variants in campus sewage water, all samples were analyzed using high-throughput next-generation sequencing (NGS) technology. We also demonstrated that sewage water-based genomic surveillance could be a preventive measure for averting outbreaks and a reliable tool for identifying novel variants that would otherwise escape from variant-specific qRT-PCR assays.

## 2. Material and Methods

### 2.1 Sample collection and downstream processing

CSIR-National Chemical Laboratory, Pune, India (18°32’31.2”N; 73°48’43.2”E) is a federal research institution and home to a closed society of approximately 5,000 employees and students (Fig. 1). Domestic water is treated via a sewerage line connected to a Phytorid STP with a capacity of 0.15 million liters per day (MLD). The sample collection included a grab-sampling method, executed following the CDC, USA guidelines of COVID-19 wastewater sampling, and approved by the Institutional Biosafety Committee (IBSC). Between November 2021 and April 2022, one litre of each of the 38 samples was collected from the inlet in a sterile plastic container between 09:00 am and 10:00 am. All the samples were processed immediately after collection per CDC, USA guidelines of COVID-19 sewage water sampling. The sampling bottle was wiped with 70% ethanol and promptly subjected to 1h pasteurization at 60°C. Of the total 1L of the sample, an aliquot of 160 ml was used for the concentration of the RNA fragments of SARS-CoV-2 using the polyethylene glycol (PEG) precipitation method (Ahmed et al., 2020). Briefly, the sample was centrifuged at 5000xg (5910 R, Eppendorf, Germany) for 10 minutes at 4°C to collect the larger debris as a pellet. Filtration was done twice, first with Whatman qualitative filter paper grade 1 with a diameter of 47 mm (Merck, USA), and then with 0.22 µm PES filter membrane (diameter 47mm) using vacuum filtration assembly (Tarsons, India). The resulting filtrate was added to a 250ml HDPE bottle (HiMedia, India), containing 8% (w/v) PEG 8000 (HiMedia, India) and 1.7% of molecular biology grade NaCl (w/v) (HiMedia, India). This mixture was incubated for 16 h at 4°C in a rotating shaker water bath (Equitron medica, India) at 175 rpm. It was centrifuged at 10000xg for 1 h at 4°C to collect the viral pellet. The supernatant was removed, and the pellet was resuspended in 560 μL of TE buffer (pH 7.0). One part of the resuspended pellet (280 μL) was stored at -80°C for future use, and the remaining (280 μL) was used to extract viral RNA. QiagenQIAmp Viral RNA Minikit (Qiagen, Germany) was used for viral RNA extraction according to the manufacturer’s instructions. The RNA was eluted in 80 µL of elution buffer and stored at -80°C for downstream analysis. After obtaining permission from the Institutional Biosafety Committee (IBSC), the used materials were disposed off.

**Fig. 1:**
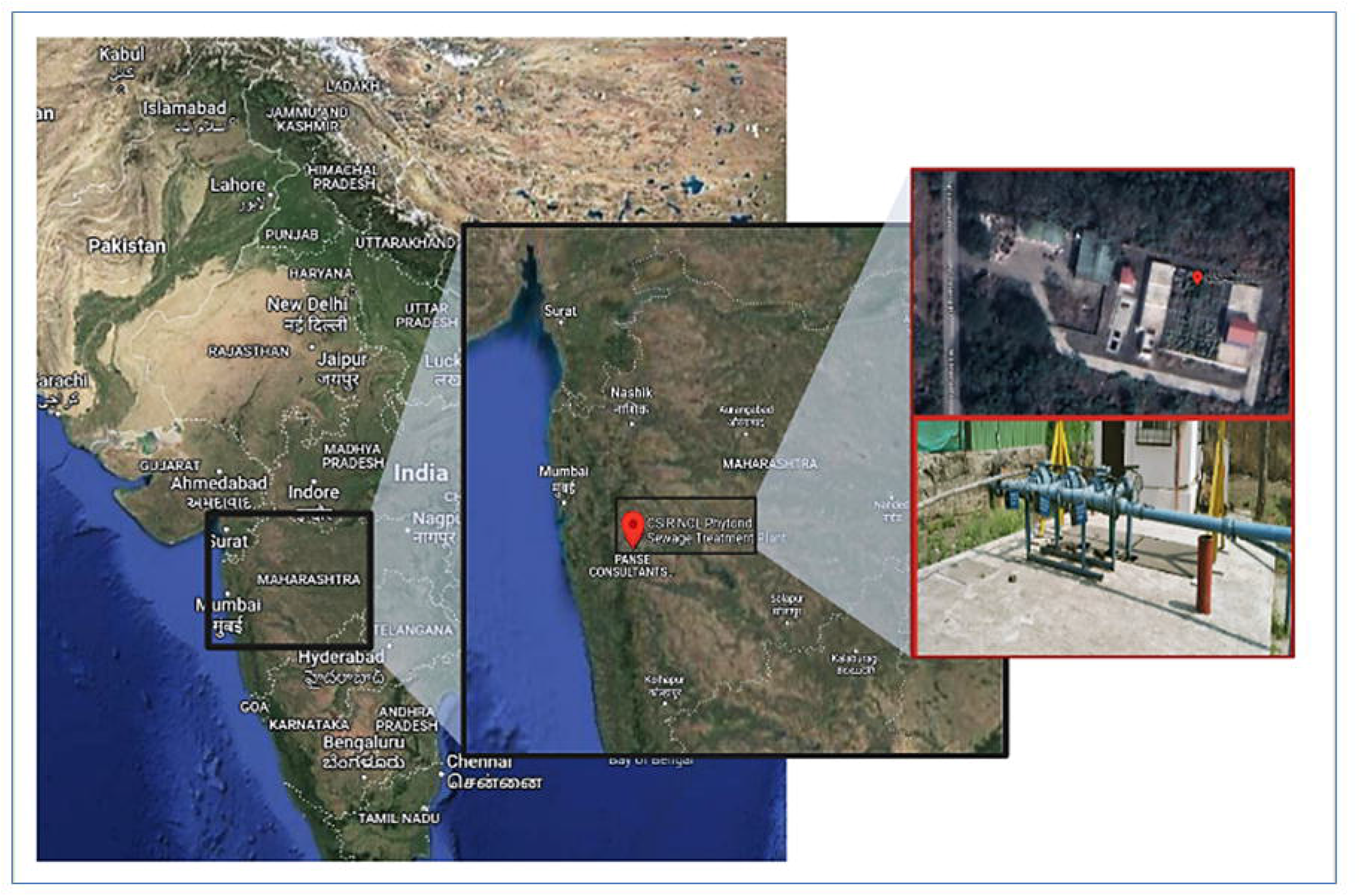
Sampling site-campus STP (18°32’31.2”N; 73°48’43.2”E)

### 2.2 Molecular detection

The qRT-PCR approach was used for SARS-CoV-2 RNA detection in sewage water samples. The experiment was conducted using an ICMR-approved GenePath Diagnostics qRT-PCR kit (GenePath DxCoViDx One v2.1.1TK-Quantitative). It is a one-tube multiplex qRT-PCR assay that includes E, N and RdRP genes of SARS-CoV-2, with the RNase P gene serving as a nucleic acid isolation control. Each sample was tested in triplicates, wherein the PCR cocktail for individual reaction contained 9.25 μL of master mix, 0.75 μL of a primer-probe mix, and 5 μL of template in a 15 μL total reaction volume. The thermal cycling protocol consisted of pre-incubation at 37°C for 5 min, reverse transcription for 7 min at 52°C, RT inactivation at 95°C for 3 min 30 sec and 40 amplification cycles of denaturation at 95°C for 5 sec, and extension at 58 °C for 35 sec. All qRT-PCR experiments were conducted using an Applied Biosystems 7500 Fast Real-Time PCR system (USA). Viral load in samples was quantified using an online Covid-19 Viral Load Calculation Tool provided by the kit’s manufacturer.

### 2.3 SARS-CoV-2 whole genome sequencing

All the samples were sequenced at the Next Generation Genomics facility at the Indian Institute of Science Education and Research (IISER), Pune, India. Sample libraries were prepared using Illumina Covid seq RUO kits (Illumina, USA). Post-annealing with random hexamers, the first strand synthesis reaction was carried out, followed by cDNA amplification in two separate multiplex PCR reactions, including primers designed to cover the entire genome of SARS-CoV-2. Using the Illumina PCR Indexes, the amplified products were further processed for tagmentation and adapter ligation, followed by enrichment and clean-up of amplicons as per the manufacturer’s instructions. Pooled libraries were quantified using the Qubit 4.0 fluorometer (Invitrogen, USA). All the pools were normalized to a final concentration of 4.0nM using the Illumina Pooling calculator. For sequencing, the final pooled library was denatured, neutralized, and adjusted to a final concentration of 1.4pM, and paired-end sequencing was performed using the Illumina NextSeq 550 sequencing platform (Illumina, USA).

### 2.4 Bioinformatic analysis

A dedicated in-house bioinformatics pipeline was used for analyzing raw data to identify the main circulating lineages in sewage water. In the first step, raw reads were checked for quality and filtered using fastp (Chen et al., 2018). After that, the reference genome of SARS-CoV-2 (MN908947.3) was used to align the filtered reads by BWA Mem (Li and Durbin, 2009).

Basic alignment and coverage statistics were determined by SAM tools coverage/bedcov (Li et al., 2009; Quinlan and Hall, 2010). iVar was used for single nucleotide variant (SNV) calling using a minimum base quality score of 20, minimum frequency 0.03, and minimum read depth of 10 (Grubaugh et al., 2019). To predict the lineage composition, we used LCS-a mixture model for determining SARS-CoV-2 variant composition in environmental samples (Valieris et al., 2022). Scripting was done in Python/Bash to manually check variant-associated mutations, followed by data plotting in R Studio.

## 3. Results

### 3.1 SARS-CoV-2 quantification

The goals of our study were to (i) detect and quantify SARS-CoV-2 genetic material in campus sewage water, (ii) monitor SARS-CoV-2 infection trend on campus, (iii) monitor viral variants of SARS-CoV-2 through NGS, and (iv) implement sewage water surveillance within the campus, incorporating it as a component of the pandemic response system. Overall, 38 sewage water samples collected from the campus STP inlet from November 2021 to April 2022 were investigated for the occurrence of SARS-CoV-2 RNA. Samples were considered positive when any two of three target genes for SARS-CoV-2 showed Ct value ≤ 35 as per kit instructions. Nineteen samples were positive, with Ct values ranging between 29.23 and 34.52, representing a 50% overall positive detection rate (Supplementary Table 1). The remaining 19 samples were negative.

Further, to understand the transmission dynamics of SARS-CoV-2 in sewage water at the community level, we calculated the viral load of each sample in copies/ml using the calculation tool provided by the qRT-PCR kit manufacturer (Fig. 2). The first significant increase in viral load was observed in sample PR-41 (111.44 copies/ml) collected on November 22, 2021 (Supplementary Table1), two weeks before the first positive case was reported on campus. Viral load was later observed to increase exponentially from January 03, 2022 (PR-50) and the highest viral load was detected on January 20, 2022 in sample PR-55 (526 copies/mL). SARS-CoV-2 RNA copies continued to be found from February 2022 to April 2022, with a relatively higher viral load in sample PR-62 (229 copies/mL). A total of 184 positive cases of COVID-19 were reported between November 2021 and April 2022 among the students, staff, and faculty, or their family members residing on the CSIR-NCL campus (Supplementary Table 2). However, the first case of COVID-19 on campus was reported to authorities on December 06, 2021. This correlates with our experimental data showing an increase in viral load 14 days before its first reporting. Pearson’s correlation for viral load showed a significant correlation (R²=0.68, p-value>7.9e^-11^) with COVID-19 cases reported on campus (Fig. 2B). Although no new COVID-19 cases were reported on campus after February 07, 2022, the SARS-CoV-2 RNA sustained with few spikes in the concentration of viral load during February 2022 to April 2022.

**Fig. 2:**
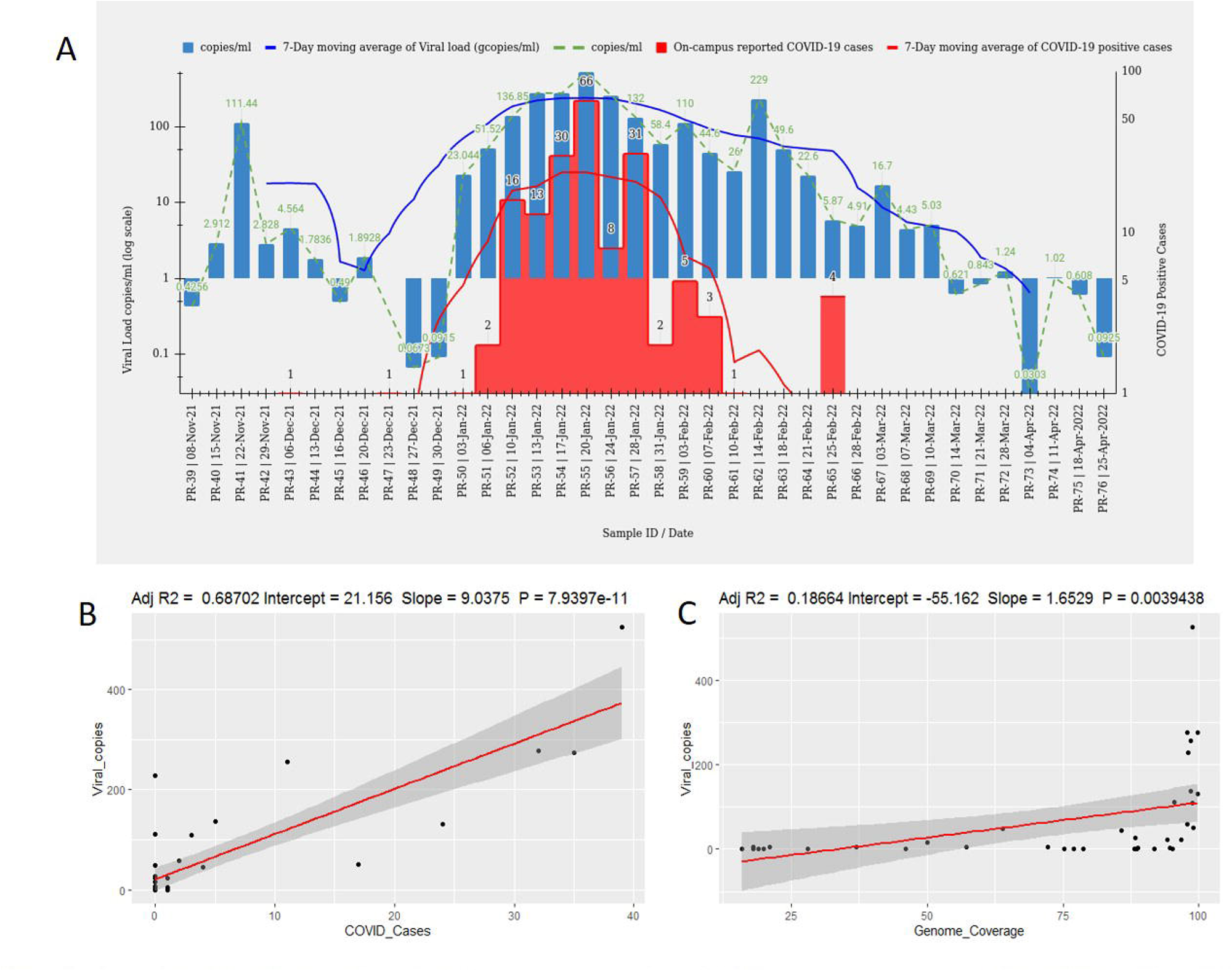
Analysis of SARS-CoV-2 viral load in WW against reported COVID-19 cases on campus. A. SARS-CoV-2 viral load dynamics in sewage water and reported COVID-19 cases on campus. B. Linear regression correlation plot of viral concentration in wastewater with reported COVID-19 cases on-campus. (Pearson correlation coefficient: R^2^ =0.68, p-value>7.9e^-11^). C. Linear regression correlation plot of viral concentration in wastewater with genome coverage (Pearson correlation coefficient: R^2^ =0.18, p-value>0.003)

### 3.2 Overview of SARS-CoV-2 sequencing, detected nucleotide substitutions and deletions

All 38 sewage water samples collected during the third wave of Omicron from November 2021 to April 2022 were sequenced using the Illumina Next Seq 500 platform. Samples were grouped into positive (n=19) and negative (n=19) categories based on SARS-CoV-2 qRT-PCR (Supplementary Table 1). Since the qRT-PCR method uses primer pairs that target specific viral fragments and the SARS-CoV-2 virus is susceptible to degradation in wastewater, that would eventually make capturing viral fragments with limited qRT-PCR primer pairs challenging. As a result, when wastewater is processed for qRT-PCR-based detection, the probability phenomenon may pose a challenge to the limited qRT-PCR primers. Currently, the whole genome NGS protocols use multiple primers targeting the entire genome, thus increasing the probability of detecting the minute amount of nucleic acid fragments in sewage water. Therefore, we sequenced all the samples regardless of qRT-PCR results to find the SARS-CoV-2 fragments and to understand the mutation profile of SARS-CoV-2 better.

A total of 124.16 million reads were generated from sequencing, of which 26.46 million reads were mapped to the SARS-CoV-2 reference genome (MN908947.3). Out of 38 sequenced samples, 27 showed mean genomic coverage above 50% ranging between 99.81% and 57.2%. At the same time, 16 out of 27 samples showed mean genome coverage uniformity above 90% (Supplementary Table1). Sequencing results of qRT-PCR positive (n=19) and negative samples (n=19) showed significant evidence for the presence of COVID-19 genomic material in the samples, supported by the linear regression analysis between viral concentration and reference genome coverage (Pearson correlation coefficient: R^2^ =0.18, p-value>0.003) (Fig. 2.C). Their reads mapped ranged between 1.7 million and 34 thousand (Fig. 3).

**Fig. 3:**
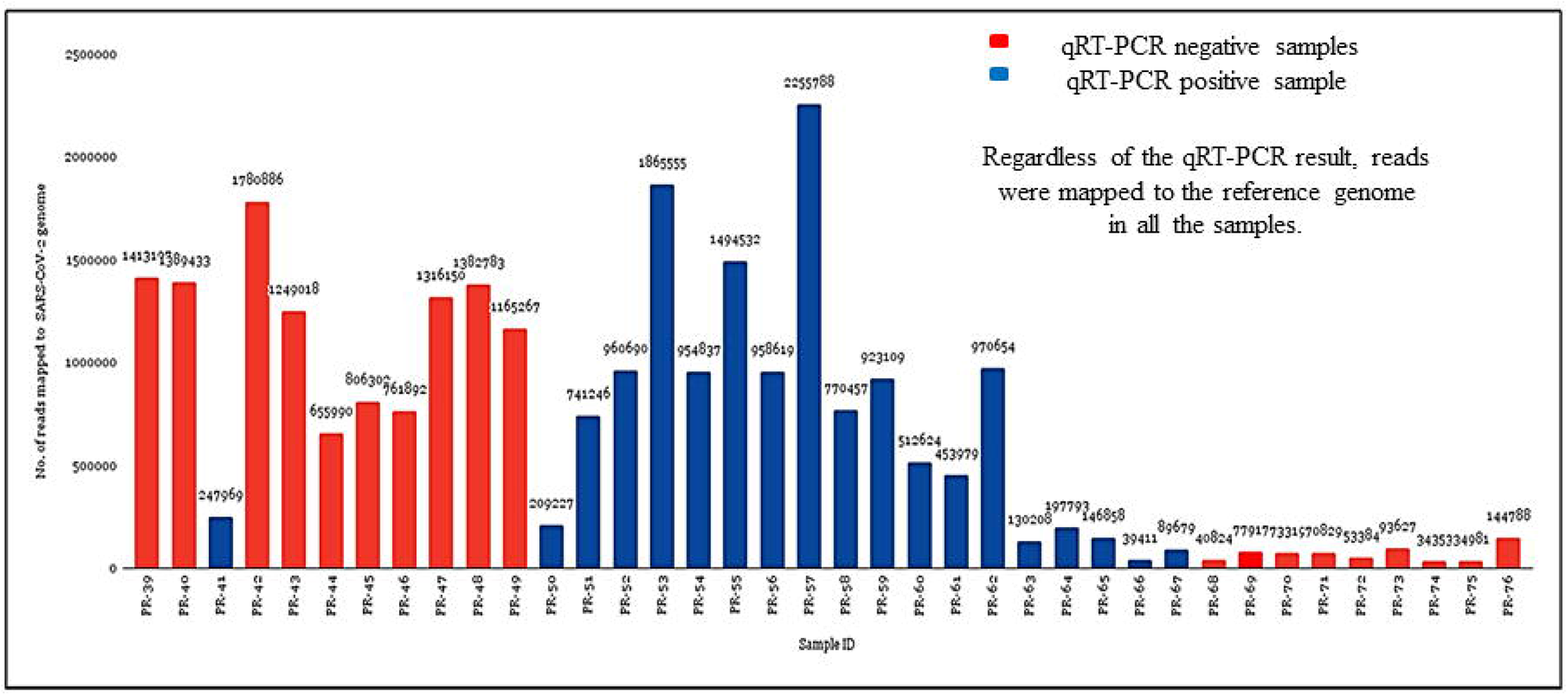
Distribution of the number of reads mapped to the SARS-CoV-2 reference genome per sample (MN908947.3)

Overall, 2108 unique nucleotide substitutions (Supplementary Fig.1) were found compared with the reference Wuhan genome of SARS-CoV-2 (MN908947.3). Among the nucleotide substitutions found: 941 mutations were in ORF1a polyprotein; 519 in ORF1ab polyprotein; 88 in ORF3a protein; 323 in spike protein; 19 in envelope protein; 48 in membrane protein; 10 in ORF6 protein; 19 in ORF7a protein; 15 in ORF7b protein; 17 in ORF8 protein; 101 in nucleocapsid phosphoprotein and 08 in ORF10 protein. A total of 119 deletions were detected in samples, 61 in ORF1a, 22 in ORF1ab, 6 in ORF3a, 22 in spike protein; 03 in membrane glycoprotein; 01 in ORF7a protein; 01 in ORF7b protein; and 03 in nucleocapsid phosphoprotein.

### 3.3 Early detection of Omicron signature mutations

Sequencing data were analyzed and searched for the presence of signature mutations of different lineages of SARS-CoV-2VOCs, Alpha (B.1.1.7), Beta (B.1.351), Gamma (P.1), Delta (B.1.617.2) and Omicron (B.1.1.529) and VOIs, Kappa (B.1.617.1), Epsilon (B.1.427, B.1.429), Eta (B.1.525), Iota (B.1.526) and Lambda (C.37) (Fig. 4). A total of 5214 signature mutations were detected, representing 16 highly transmissible variants of COVID-19. Samples collected from November to December 2021 were highly dominated by Delta lineage (Fig. 5A). The transition of Omicron lineage over delta lineage was observed in late December 2021, which is supported by the detection of a higher number of Omicron-associated signature mutations from January 2022 onwards (Fig. 5B), Omicron mutations ranging between 30-42 nucleotide substitutions per sample, followed by continuous detection till March 2022. The Omicron lineage of SARS-CoV-2 contains 62 characteristic deletions and non-synonymous nucleotide substitutions, including 30 substitutions on the spike region (Dejnirattisai et al., 2022). We detected the first signal of Omicron in sample PR-39 collected on November 08, 2021. A total of 11 signature mutations were detected, 09 from the spike gene (S:P681H, S:T478K, S:D614G, S:H655Y, S:N679K, S:N764K, S:L452R, S:D796Y, S:T478K;), 01 from N gene (N:P13L) and 01 from ORF1a gene (ORF1a:K856R). Similarly, 05 markers were detected in sample PR-40 (S:L452R, S:T478K, S:D614G, ORF1a:T3255I, ORF1a:P3395H), 06 in PR-41 (S:L452R, S:T478K, S:D614G, N:P13L, ORF1a:T3255I, ORF1a:P3395H) and 04 in PR-42 (S:L452R, S:T478K, S:D614G, ORF1a:T3255I) (Supplementary Tables 3 and 4).

**Fig. 4:**
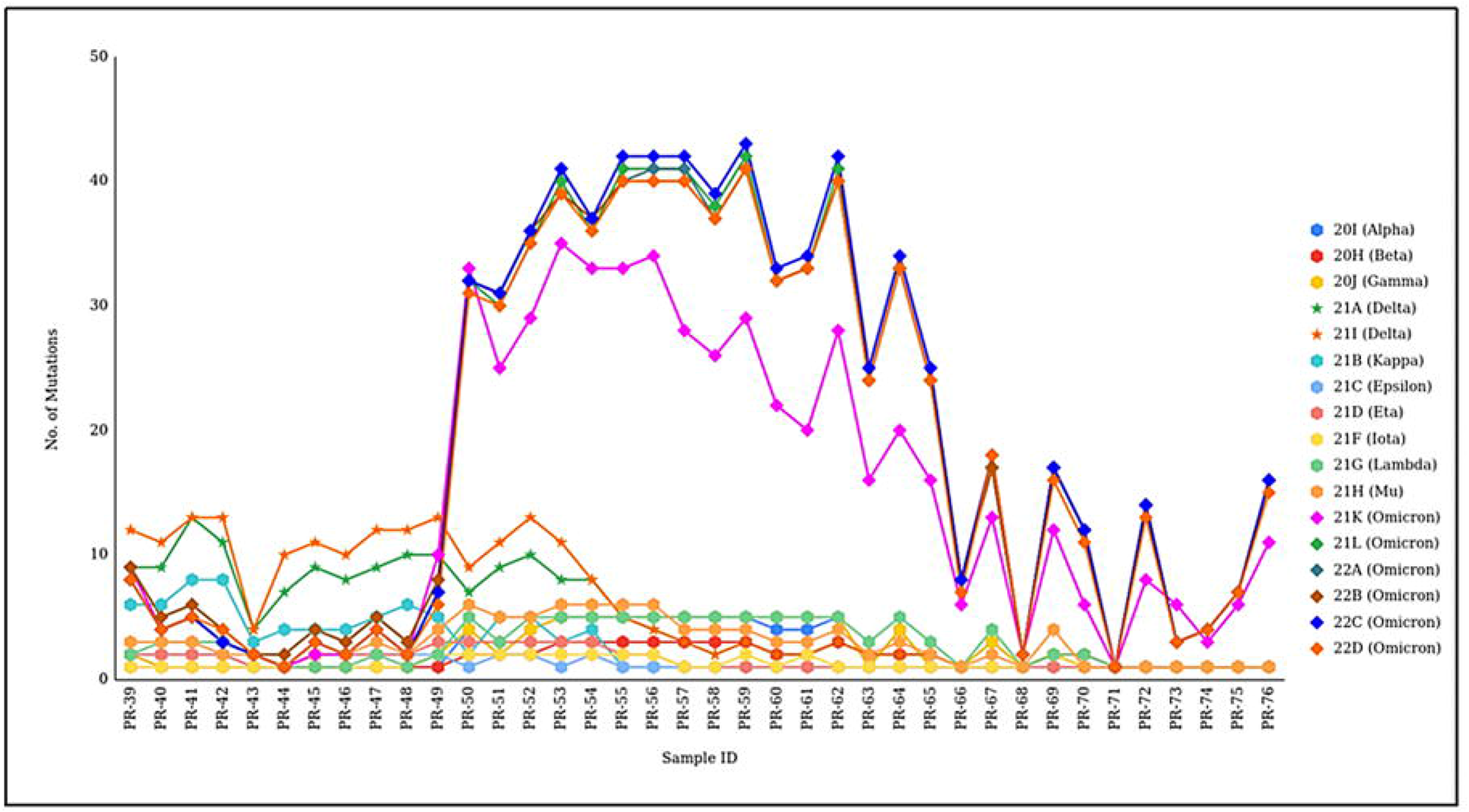
Detection of transition between SARS-CoV-2 VOC Delta and Omicron by sewage water genomic surveillance.

**Fig. 5:**
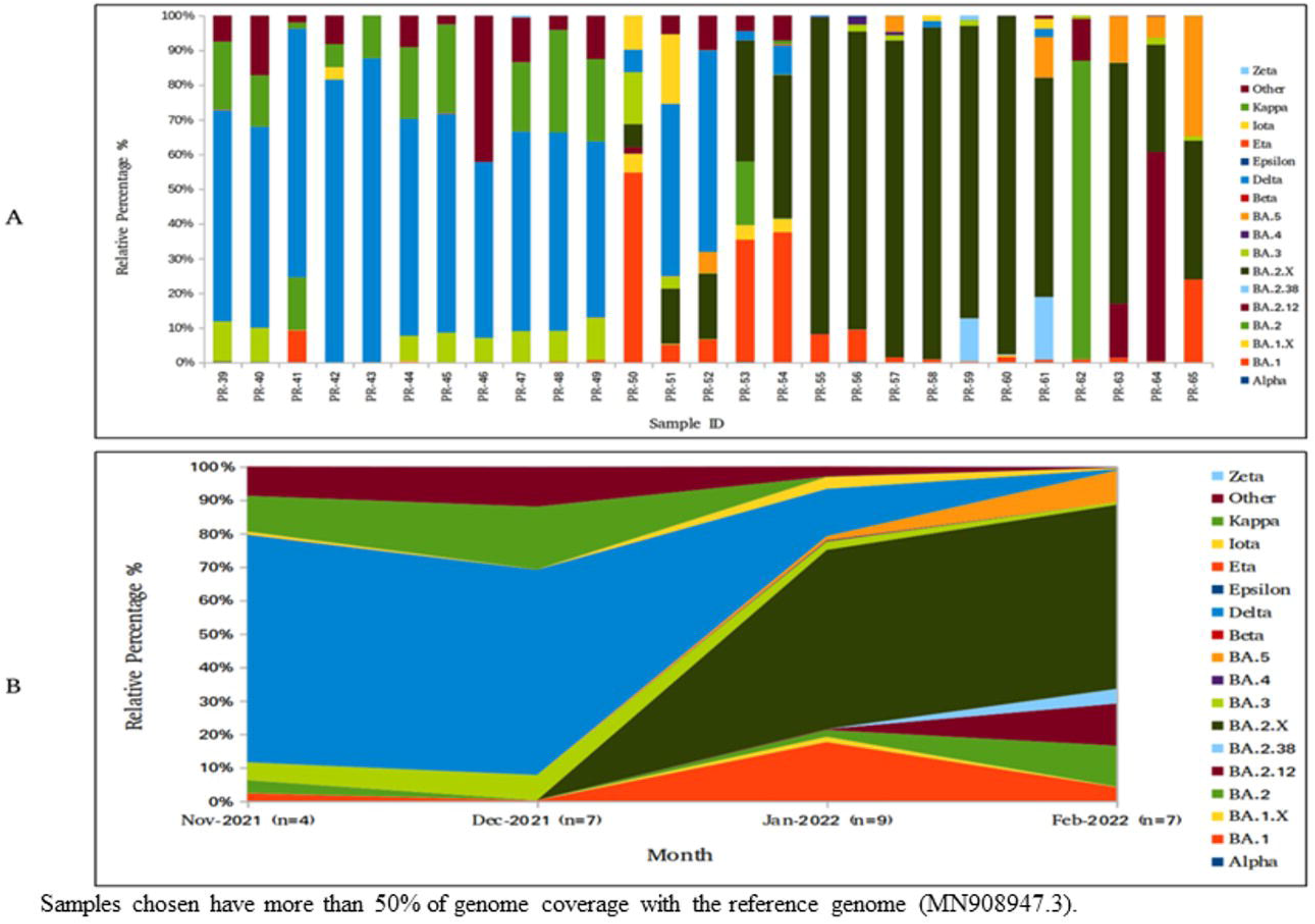
LCS analysis-based distribution of different lineages/sub-lineages of SARS-CoV-2. **A**. Relative abundance in the percentage of the identified SARS-CoV-2 lineages/sub-lineages**. B**. Proportion of SARS-CoV-2 lineages from November 2021-February, 2022.

### 3.4 SARS-CoV-2 prevalence and lineage trends in sewage water

LCS was used to ascertain the relative frequencies of SARS-CoV-2 variants in sewage water. A total of 426 lineages were detected, representing 16 highly transmissible variants of SARS-CoV-2 (VOCs and VOIs). We found that the Delta lineage dominated in (B.1.617.2) November 2021 (68%) and December 2021 (61%); after that Kappa (10% and 18%), BA.3 (5% and 7%), BA.2 (3%), BA.1 (2% and 0.1%) and others (8%) (Fig.5).The Omicron family lineages increased exponentially from January 2022 onwards and fully replaced Delta lineage by February 2022. BA.2.X was the most dominant lineage (53.6% and 55%), followed by BA.1 (17% and 4%), BA.2 (2% and 12%), BA.2.12 (0% and 12%), BA.5 (1% and 9%) and Iota (3% and 0.4%) respectively in January and February 2022. Finally, the linear-mixed effect model was employed to understand the lineage trend from November 2021 to April 2022. The Omicron lineages (BA.1, BA.2, BA.4, and BA.5) showed an increasing trend, whereas the Delta lineage (B.1.617.2) showed a decreasing trend from November 2021 to February 2022(Fig. 6). Linear mixed model based monthly proportion of SARS-CoV-2 lineages/sub-lineages.

**Fig. 6:**
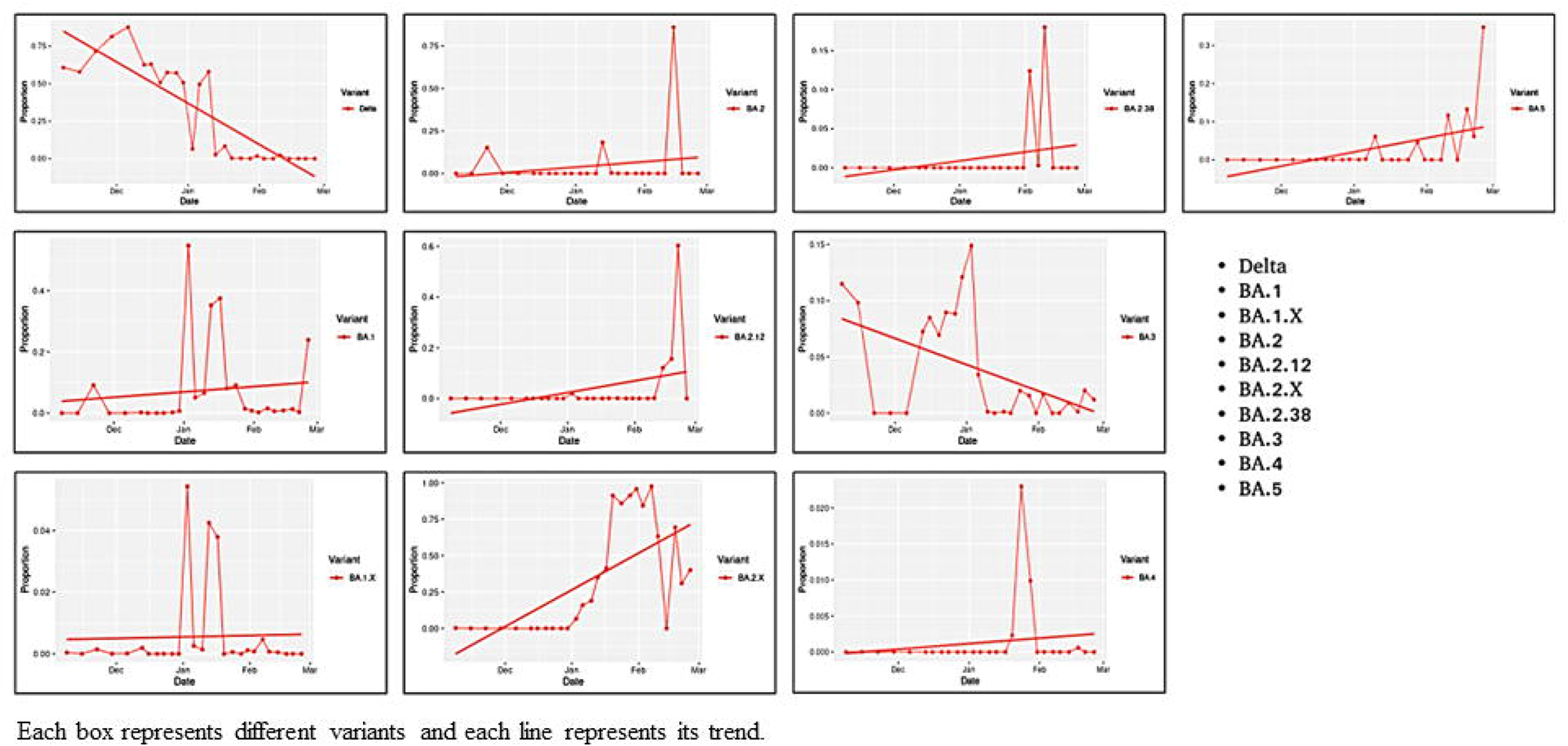
Linear mixed model based monthly proportion of SARS-CoV-2 lineages/sub-lineages.

## Discussion

Wastewater surveillance provides information on the viral load of SARS-CoV-2 and the introduction of new variants (VOI/VOC) in the community (Johnson et al., 2022; Oloye et al., 2022). WBE can be highly prioritised in India, especially with the emergence of new variants and limited COVID-19 testing. Our study demonstrates how genomic surveillance of sewage water from on-campus STP can be utilised for detection, monitoring, and evaluation of the spread of SARS-CoV-2, which effectively helps to take decisions for the protection against SARS-CoV-2 in academic institutes/universities as documented by Betancourt et al., 2021; Corchis-Scottet al., 2021; Gibas C et al., 2021; Kantor et al., 2022; Karthikeyan et al., 2021; Mangwana et al., 2022; Wang et al., 2022; and Vo et al., 2022 across the world. Several studies have revealed the detection of SARS-CoV-2 from wastewater in India (Arora et al., 2022; Dharmadhikari et al., 2022;Joshi et al., 2022). However, to the best of our knowledge, this is the first study from the country to use the WBE technique to prevent the spread of SARS-CoV-2 on an academic campus.

A higher viral load of SARS-CoV-2 is strongly linked to increased disease virulence, progression, and death rate (Fajnzylber et al., 2020; Kawasuji et al., 2022). Serosurveillance of COVID-19 in Pune city showed a rise in daily new cases from the last week of December, 2021 (Supplementary Fig.3). Whereas by the end of January, 2022 the pick of daily active and new cases were observed, which decreased by the end of March, 2022 (http://cms.unipune.ac.in/~bspujari/Covid19/Pune2/). The trend of daily active and new cases reported in the serosurveillance complements the clinical case reported on campus. We observed an increase in the viral load of SARS-CoV-2 in sewage water prior to the first clinical case reported on campus. Recent studies from the USA (Al-Falsity et al., 2022; Karthikeyan et al., 2021; Wu et al., 2022), Mexico(Padilla-Reyes et al., 2022); Greece (Galani et al., 2022), England (Martin et al., 2020; Morvan et al., 2022), South Africa (Johnson et al., 2022), Brazil (de Araújo et al., 2022); Japan (Malla et al., 2022), and India (Joshi et al., 2022) also revealed similar results which are consistent with our findings. The strong link between SARS-CoV-2 viral load in sewage water and reports of newly confirmed COVID-19 cases per day suggests that these infected people contribute substantial viral load to the sewage water (Hasan et al., 2021). These findings demonstrate WBE’s early warning capability (Kirby et al., 2022; Lastra et al., 2022; Wang et al., 2022).

In most locations, the epidemic caused by the Omicron variant had a lower impact on health than previous COVID-19 waves due to increased herd immunity and possibly diminished nature of SARS-CoV-2 infections (Nealon and Cowling, 2022). Nyberg and colleagues also found that the Omicron variant is less severe than the Delta lineage of SARS-CoV-2 in terms of mortality and hospitalization (Nyberg et al., 2022; Post and Lorenzo-Redondo, 2022). Although the viral load was higher and continuously detected in the on-campus STP sewage water, the reported COVID-19-positive cases were low. The aforementioned higher viral load could be correlated with the incidence of several asymptomatic COVID-19-positive individuals on campus who did not test themselves for the underlying condition. Another attribute could be the overestimation of the number of COVID-19-positive cases. Therefore, our study supports using the WBE approach as a realistic indicator for monitoring SARS-CoV-2 and future outbreaks.

The qRT-PCR method has been established as a standard for assessing viral nucleic acid from patients and has also been used for environmental studies with its utility in WBE studies (Ahmed et al., 2022; Johnson et al., 2022). Initially, researchers depended on qRT-PCR and Sanger sequencing to establish the existence of viral fragments of SARS-CoV-2 in wastewater. WBE studies are founded on SARS-CoV-2 fecal shedding (Crank et al., 2022; Li et al., 2022); however, the fecal matter would be vastly diluted by the addition of household sewage water and rain, establishing a probability framework for detecting viral fragments (Novoa et al., 2022; Wade et al., 2022). We detected SARS-CoV-2 in 100% of qRT-PCR negative samples, concluding that the amplicon sequencing protocol is much more sensitive in detecting the SARS-CoV-2 fragments in sewage water than qRT-PCR. Therefore, our study emphasizes the efforts of sequencing, irrespective of the qRT-PCR result, not only for viral detection but the monitoring of newly emerging variants, novel mutations, and evolutionary studies of SARS-CoV-2.

Variant tracking from wastewater cannot confirm the presence of a specific variant because COVID-19 genetic material in wastewater is often fragmented (Gregory et al., 2022; Wolfe et al., 2022); as a result, it poses a significant challenge to detect all characteristics mutations of a particular variant linked to a single genome. However, detecting multiple mutations associated with a variant or unique mutations absent in others increases the confidence of the presence of a specific variant in wastewater (Kirby et al., 2022; Zahmatkesh et al., 2022). Therefore, we also analyzed all the samples manually using the iVar SNP calling pipeline to gain in-depth insights into the mutation profile of low-sequenced samples. Surprisingly, we found Omicron signals in early November 2021, whereas Omicron was discovered for the first time in Botswana on November 11, 2021 (Rahmani and Rezaei, 2022). Later, WHO designated variant B.1.1529 as a VOC on November 26, 2021. India reported the first case of Omicron on December 02, 2021, in Karnataka state, followed by the first detection of Omicron in Maharashtra state on December 04, 2021. However, we detected several unique Omicron-associated mutations in our data in early November 2021 (S:N679K, S:N764K, S:D796Y, N:P13L, ORF1a:T3255I, ORF1a:K856R, ORF1a:P3395H, N:S413R) which provides substantial evidence that Omicron variant was likely present earlier on campus in India before it was first detected clinically across the globe. Similar reports from the USA and France also provide strong evidence for the presence of the Omicron variant in early November 2021 in community wastewater before the first detection from the clinical sample (Ferré et al., 2022; Kirby et al., 2022).

Wastewater testing captures contributions from all residents within a locality, which provides a less skewed outcome of viral genetic diversity and public health (Karthikeyan et al., 2021; Safford et al., 2022; Sims and Kasprzyk-Hordern, 2020). Clinical testing has limitations such as providing data only on those who agree to be tested, missing asymptomatic cases, resource allocation, and affordability concerns. We used LCS, a statistical model for estimating SARS-CoV-2 variants composition in pooled samples (Valieris et al., 2022). Our samples showed significant SARS-CoV-2 lineage diversity across months, including lineages that were not reported from clinical samples, highlighting the discrepancies in clinical testing and sequencing. We identified the shift around January and February 2022, defined by a sharp rise of Omicron lineages (BA.1, BA.2, BA.3, BA.4, BA.5) and a decrease of previously dominant lineages (Delta and Kappa).

Furthermore, BA.4 and BA.5 signals were detected in January and February 2022, two months earlier than their first report from a Telangana, India clinical sample in late March 2022. Hence, the emergence of variants in wastewater indicates that a sizeable number of people were infected with specific variants, contributing to the total viral load. One limitation of our study is the lack of sequencing data of clinically reported positive cases from campus, which could have aided in correlating with the emerging and prevailing variants of SARS-CoV-2 circulating on the campus.

Throughout the surveillance period, we informed decision makers (COVID-19 task force) about SARS-CoV-2 detection in sewage water of campus-STP approximately 48 h after sample collection, which helped in policy-making decisions to alert the community on campus. Last, our work illustrates the utility of WBE for monitoring COVID-19 on campus, and similar settings could help manage SARS-CoV-2 and other future emerging pathogens of concern.

## 4. Conclusions

This study concludes that the use of WBE at the community level would be an effective means of preventing SARS-CoV-2 outbreaks. An increase in viral load in the community detected early with the help qRT-PCR approach indicates its importance as an alarming feature of WBE. The amplicon sequencing protocol can be sensitive in detecting the fragments of SARS-CoV-2 in wastewater, which were failed to get caught in qRT-PCR. It can also be concluded that to provide comprehensive information about population infection dynamics; more emphasis should be on the amplicon sequencing-based approaches to gain information on the emerging variants. Overall, this work has shown that implementing real-time WBE on-campus and integrating it with the response system would effectively prevent and contain future outbreaks.

## Data availability

All data produced in the present study are available upon reasonable request to the authors. Clinical COVID-19 data was used from CMS dashboard (http://cms.unipune.ac.in/~bspujari/Covid19/Pune2/)

## Credit authorship contribution statement

**Vinita Malik:** Conceptualization, Data curation, Formal analysis, Investigation, Methodology, Validation, Roles/Writing – original draft

**Vinay Rajput**: Data curation, Formal analysis, Investigation, Methodology, Software, Visualization, Roles/Writing – original draft

**Rinka Pramanik**: Investigation, Methodology, Visualization, Roles/Writing – original draft

**Rachel Samson**: Writing – review & editing

**Rakeshkumar Yadav**: Writing – review & editing

**Pradnya Kadam**: Methodology

**Nikita Shah**: Methodology

**Rutuja Sawant**: Methodology

**Unnati Bhalerao**: Methodology

**Manisha Tupekar**: Methodology

**Soumen Khan**: Methodology

**Priyanki Shah**: Funding acquisition, Project administration, Supervision

**LS Shashidhara**: Funding acquisition, Project administration, Supervision

**Sanjay Kamble**: Investigation, Formal analysis

**Syed Dastager**: Investigation, Formal analysis

Krishanpal Karmodiya: Formal analysis

**Mahesh Dharne**: Conceptualization, Funding acquisition, Investigation, Project administration, Resources, Supervision, Writing – review & editing.

## Acknowledgments

This work is funded by a grant from Rockefeller Foundation, USA (Project code GAP 332926). The authors express their gratitude to the Director General, Council of Scientific and Industrial Research (CSIR), New Delhi; Directors of CSIR-NCL,, IISER-Pune and Pune Knowledge Cluster, for encouragement and support. Authors are indebted to Prof. Aurnab Ghose (IISER-Pune for grant administration support and discussions). Thanks to Dr. Narendra Kadoo (Chair, NCL Covid task force) and Dr. Chetan Gadgil (CSIR-NCL) for their valuable suggestions. Thanks to the Administrative office for providing on-campus case records and the Engineering and Civil services section of CSIR-NCL for all the support during study. The manuscript has been thoroughly checked for plagiarism content using iThenticate software.

## References

Ahmed, W., Angel, N., Edson, J., Bibby, K., Bivins, A., O’Brien, J.W., Choi, P.M., Kitajima, M., Simpson, S.L., Li, J., 2020. First confirmed detection of SARS-CoV-2 in untreated wastewater in Australia: a proof of concept for the wastewater surveillance of COVID-19 in the community. Science of the Total Environment 728, 138764.

Ahmed, W., Smith, W.J.M., Metcalfe, S., Jackson, G., Choi, P.M., Morrison, M., Field, D., Gyawali, P., Bivins, A., Bibby, K., 2022. Comparison of RT-qPCR and RT-dPCR platforms for the trace detection of SARS-CoV-2 RNA in wastewater. ACS ES&T Water.

Ahmed, W., Tscharke, B., Bertsch, P.M., Bibby, K., Bivins, A., Choi, P., Clarke, L., Dwyer, J., Edson, J., Nguyen, T.M.H., 2021. SARS-CoV-2 RNA monitoring in wastewater as a potential early warning system for COVID-19 transmission in the community: A temporal case study. Science of The Total Environment 761, 144216.

Al-Faliti, M., Kotlarz, N., McCall, C., Harris, A.R., Smith, A.L., Stadler, L.B., de los Reyes, F.L.I.I.I., Delgado Vela, J., 2022. Comparing Rates of Change in SARS-CoV-2 Wastewater Load and Clinical Cases in 19 Sewersheds Across Four Major Metropolitan Areas in the United States. ACS ES&T Water. https://doi.org/10.1021/acsestwater.2c00106

Amman, F., Markt, R., Endler, L., Hupfauf, S., Agerer, B., Schedl, A., Richter, L., Zechmeister, M., Bicher, M., Heiler, G., 2022. Viral variant-resolved wastewater surveillance of SARS-CoV-2 at national scale. Nat Biotechnol 1–9.

Bar-Or, I., Weil, M., Indenbaum, V., Bucris, E., Bar-Ilan, D., Elul, M., Levi, N., Aguvaev, I., Cohen, Z., Shirazi, R., 2021. Detection of SARS-CoV-2 variants by genomic analysis of wastewater samples in Israel. Science of The Total Environment 789, 148002.

Chen, R.E., Zhang, X., Case, J.B., Winkler, E.S., Liu, Y., VanBlargan, L.A., Liu, J., Errico, J.M., Xie, X., Suryadevara, N., 2021. Resistance of SARS-CoV-2 variants to neutralization by monoclonal and serum-derived polyclonal antibodies. Nat Med 27, 717–726.

Chen, S., Zhou, Y., Chen, Y., Gu, J., 2018. fastp: an ultra-fast all-in-one FASTQ preprocessor. Bioinformatics 34, i884–i890.

Clark, A.E., Wang, Z., Cantarel, B., Kanchwala, M., Xing, C., Chen, L., Irwin, P., Xu, Y., Oliver, D., Lee, F., 2021. Multiplex fragment analysis identifies SARS-CoV-2 variants. MedRxiv.

Crank, K., Chen, W., Bivins, A., Lowry, S., Bibby, K., 2022. Contribution of SARS-CoV-2 RNA shedding routes to RNA loads in wastewater. Science of The Total Environment 806, 150376.

Crits-Christoph, A., Kantor, R.S., Olm, M.R., Whitney, O.N., Al-Shayeb, B., Lou, Y.C., Flamholz, A., Kennedy, L.C., Greenwald, H., Hinkle, A., 2021. Genome sequencing of sewage detects regionally prevalent SARS-CoV-2 variants. mBio 12, e02703–20.

de Araújo, J.C., Mota, V.T., Teodoro, A., Leal, C., Leroy, D., Madeira, C., Machado, E.C., Dias, M.F., Souza, C.C., Coelho, G., Bressani, T., Morandi, T., Freitas, G.T.O., Duarte, A., Perdigão, C., Tröger, F., Ayrimoraes, S., de Melo, M.C., Laguardia, F., Reis, M.T.P., Mota, C., Chernicharo, C.A.L., 2022. Long-term monitoring of SARS-CoV-2 RNA in sewage samples from specific public places and STPs to track COVID-19 spread and identify potential hotspots. Science of The Total Environment 838, 155959. https://doi.org/10.1016/j.scitotenv.2022.155959

Dejnirattisai, W., Huo, J., Zhou, D., Zahradník, J., Supasa, P., Liu, C., Duyvesteyn, H.M.E., Ginn, H.M., Mentzer, A.J., Tuekprakhon, A., 2022. SARS-CoV-2 Omicron-B. 1.1. 529 leads to widespread escape from neutralizing antibody responses. Cell 185, 467–484.

Dharmadhikari, T., Rajput, V., Yadav, R., Boargaonkar, R., Patil, D., Kale, S., Kamble, S.P., Dastager, S.G., Dharne, M.S., 2022. High throughput sequencing based direct detection of SARS-CoV-2 fragments in wastewater of Pune, West India. Science of The Total Environment 807, 151038.

Duong, D.B., King, A.J., Grépin, K.A., Hsu, L.Y., Lim, J.F.Y., Phillips, C., Thai, T.T., Venkatachalam, I., Vogt, F., Yam, E.L.Y., 2022. Strengthening national capacities for pandemic preparedness: a cross-country analysis of COVID-19 cases and deaths. Health Policy Plan 37, 55–64.

Fajnzylber, J., Regan, J., Coxen, K., Corry, H., Wong, C., Rosenthal, A., Worrall, D., Giguel, F., Piechocka-Trocha, A., Atyeo, C., 2020. SARS-CoV-2 viral load is associated with increased disease severity and mortality. Nat Commun 11, 1–9.

Galani, A., Aalizadeh, R., Kostakis, M., Markou, A., Alygizakis, N., Lytras, T., Adamopoulos, P.G., Peccia, J., Thompson, D.C., Kontou, A., 2022. SARS-CoV-2 wastewater surveillance data can predict hospitalizations and ICU admissions. Science of The Total Environment 804, 150151.

Gregory, D.A., Trujillo, M., Rushford, C., Flury, A., Kannoly, S., San, K.M., Lyfoung, D., Wiseman, R.W., Bromert, K., Zhou, M.-Y., 2022. Genetic Diversity and Evolutionary Convergence of Cryptic SARS-CoV-2 Lineages Detected Via Wastewater Sequencing. medRxiv.

Grimaldi, A., Panariello, F., Annunziata, P., Giuliano, T., Daniele, M., Pierri, B., Colantuono, C., Salvi, M., Bouché, V., Manfredi, A., 2022. Improved SARS-CoV-2 sequencing surveillance allows the identification of new variants and signatures in infected patients. Genome Med 14, 1–15.

Grubaugh, N.D., Gangavarapu, K., Quick, J., Matteson, N.L., de Jesus, J.G., Main, B.J., Tan, A.L., Paul, L.M., Brackney, D.E., Grewal, S., 2019. An amplicon-based sequencing framework for accurately measuring intrahost virus diversity using PrimalSeq and iVar. Genome Biol 20, 1–19.

He, X., Hong, W., Pan, X., Lu, G., Wei, X., 2021. SARSLCoVL2 Omicron variant: characteristics and prevention. MedComm (Beijing) 2, 838–845.

Hillary, L.S., Malham, S.K., McDonald, J.E., Jones, D.L., 2020. Wastewater and public health: the potential of wastewater surveillance for monitoring COVID-19. Curr Opin Environ Sci Health 17, 14–20.

Izquierdo-Lara, R., Elsinga, G., Heijnen, L., Munnink, B.B.O., Schapendonk, C.M.E., Nieuwenhuijse, D., Kon, M., Lu, L., Aarestrup, F.M., Lycett, S., 2021. Monitoring SARS-CoV-2 circulation and diversity through community wastewater sequencing, the Netherlands and Belgium. Emerg Infect Dis 27, 1405.

Jha, P., Deshmukh, Y., Tumbe, C., Suraweera, W., Bhowmick, A., Sharma, S., Novosad, P., Fu, S.H., Newcombe, L., Gelband, H., 2022. COVID mortality in India: National survey data and health facility deaths. Science (1979) 375, 667–671.

Johnson, R., Sharma, J.R., Ramharack, P., Mangwana, N., Kinnear, C., Viraragavan, A., Glanzmann, B., Louw, J., Abdelatif, N., Reddy, T., 2022. Tracking the circulating SARS-CoV-2 variant of concern in South Africa using wastewater-based epidemiology. Sci Rep 12, 1–12.

Joshi, M., Kumar, M., Srivastava, V., Kumar, D., Rathore, D.S., Pandit, R., Graham, D.W., Joshi, C.G., 2022. Genetic sequencing detected the SARS-CoV-2 delta variant in wastewater a month prior to the first COVID-19 case in Ahmedabad (India). Environmental Pollution 310, 119757.

Karthikeyan, S., Nguyen, A., McDonald, D., Zong, Y., Ronquillo, N., Ren, J., Zou, J., Farmer, S., Humphrey, G., Henderson, D., 2021. Rapid, large-scale wastewater surveillance and automated reporting system enable early detection of nearly 85% of COVID-19 cases on a university campus. mSystems 6, e00793–21.

Kawasuji, H., Morinaga, Y., Tani, H., Yoshida, Y., Takegoshi, Y., Kaneda, M., Murai, Y., Kimoto, K., Ueno, A., Miyajima, Y., 2022. SARSLCoVL2 RNAemia with a higher nasopharyngeal viral load is strongly associated with disease severity and mortality in patients with COVIDL19. J Med Virol 94, 147–153.

Kirby, A.E., Welsh, R.M., Marsh, Z.A., Alexander, T.Y., Vugia, D.J., Boehm, A.B., Wolfe, M.K., White, B.J., Matzinger, S.R., Wheeler, A., 2022. Notes from the field: early evidence of the SARS-CoV-2 B. 1.1. 529 (Omicron) variant in community wastewater—United States, November–December 2021. Morbidity and Mortality Weekly Report 71, 103.

Lastra, A., Botello, J., Pinilla, A., Urrutia, J.I., Canora, J., Sánchez, J., Fernández, P., Candel, F.J., Zapatero, A., Ortega, M., 2022. SARS-CoV-2 detection in wastewater as an early warning indicator for COVID-19 pandemic. Madrid region case study. Environ Res 203, 111852.

Li, H., Durbin, R., 2009. Fast and accurate short read alignment with Burrows– Wheeler transform. bioinformatics 25, 1754–1760.

Li, H., Handsaker, B., Wysoker, A., Fennell, T., Ruan, J., Homer, N., Marth, G., Abecasis, G., Durbin, R., 2009. The sequence alignment/map format and SAMtools. Bioinformatics 25, 2078–2079.

Li, X., Kulandaivelu, J., Guo, Y., Zhang, S., Shi, J., O’Brien, J., Arora, S., Kumar, M., Sherchan, S.P., Honda, R., 2022. SARS-CoV-2 shedding sources in wastewater and implications for wastewater-based epidemiology. J Hazard Mater 432, 128667.

Malla, B., Thakali, O., Shrestha, S., Segawa, T., Kitajima, M., Haramoto, E., 2022. Application of a high-throughput quantitative PCR system for simultaneous monitoring of SARS-CoV-2 variants and other pathogenic viruses in wastewater. Science of the Total Environment 853, 158659.

Mangwana, N., Archer, E., Muller, C.J.F., Preiser, W., Wolfaardt, G., Kasprzyk-Hordern, B., Carstens, A., Brocker, L., Webster, C., McCarthy, D., 2022. Sewage surveillance of SARS-CoV-2 at student campus residences in the Western Cape, South Africa. Science of The Total Environment 851, 158028.

Markt, R., Endler, L., Amman, F., Schedl, A., Penz, T., Büchel-Marxer, M., Grünbacher, D., Mayr, M., Peer, E., Pedrazzini, M., 2022. Detection and abundance of SARS-CoV-2 in wastewater in Liechtenstein, and the estimation of prevalence and impact of the B. 1.1. 7 variant. J Water Health 20, 114–125.

Martin, J., Klapsa, D., Wilton, T., Zambon, M., Bentley, E., Bujaki, E., Fritzsche, M., Mate, R., Majumdar, M., 2020. Tracking SARS-CoV-2 in sewage: evidence of changes in virus variant predominance during COVID-19 pandemic. Viruses 12, 1144.

Mizumoto, K., Kagaya, K., Zarebski, A., Chowell, G., 2020. Estimating the asymptomatic proportion of coronavirus disease 2019 (COVID-19) cases on board the Diamond Princess cruise ship, Yokohama, Japan, 2020. Eurosurveillance 25, 2000180.

Morvan, M., Lojacomo, A., Souque, C., Wade, M., Hoffmann, T., Pouwels, K., Singer, A., Bunce, J., Engeli, A., Grimsley, J., 2022. Estimating SARS-CoV-2 prevalence from large-scale wastewater surveillance: insights from combined analysis of 44 sites in England. International Journal of Infectious Diseases 116, S24.

Nealon, J., Cowling, B.J., 2022. Omicron severity: milder but not mild. Lancet 399, 412.

Nishiura, H., Kobayashi, T., Miyama, T., Suzuki, A., Jung, S., Hayashi, K., Kinoshita, R., Yang, Y., Yuan, B., Akhmetzhanov, A.R., 2020. Estimation of the asymptomatic ratio of novel coronavirus infections (COVID-19). International journal of infectious diseases 94, 154–155.

Novoa, B., Ríos-Castro, R., Otero-Muras, I., Gouveia, S., Cabo, A., Saco, A., Rey-Campos, M., Pájaro, M., Fajar, N., Aranguren, R., 2022. Wastewater and marine bioindicators surveillance to anticipate COVID-19 prevalence and to explore SARS-CoV-2 diversity by next generation sequencing: One-year study. Science of the Total Environment 833, 155140.

Nyberg, T., Ferguson, N.M., Nash, S.G., Webster, H.H., Flaxman, S., Andrews, N., Hinsley, W., Bernal, J.L., Kall, M., Bhatt, S., 2022. Comparative analysis of the risks of hospitalisation and death associated with SARS-CoV-2 omicron (B. 1.1. 529) and delta (B. 1.617. 2) variants in England: a cohort study. The Lancet 399, 1303–1312.

Padilla-Reyes, D.A., Álvarez, M.M., Mora, A., Cervantes-Avilés, P.A., Kumar, M., Loge, F.J., Mahlknecht, J., 2022. Acquired insights from the long-term surveillance of SARS-CoV-2 RNA for COVID-19 monitoring: The case of Monterrey Metropolitan Area (Mexico). Environ Res 210, 112967. https://doi.org/10.1016/j.envres.2022.112967

Pérez-Cataluña, A., Chiner-Oms, Á., Cuevas-Ferrando, E., Díaz-Reolid, A., Falcó, I., Randazzo, W., Girón-Guzmán, I., Allende, A., Bracho, M.A., Comas, I., 2022. Spatial and temporal distribution of SARS-CoV-2 diversity circulating in wastewater. Water Res 211, 118007.

Post, L.A., Lorenzo-Redondo, R., 2022. Omicron: fewer adverse outcomes come with new dangers. The Lancet 399, 1280–1281.

Quinlan, A.R., Hall, I.M., 2010. BEDTools: a flexible suite of utilities for comparing genomic features. Bioinformatics 26, 841–842. https://doi.org/10.1093/bioinformatics/btq033

Rahmani, S., Rezaei, N., 2022. SARS-CoV-2 Omicron (B. 1.1. 529) Variant: No Time to Wait! Acta Bio Medica: Atenei Parmensis 93.

Randazzo, W., Cuevas-Ferrando, E., Sanjuán, R., Domingo-Calap, P., Sánchez, G., 2020. Metropolitan wastewater analysis for COVID-19 epidemiological surveillance. Int J Hyg Environ Health 230, 113621.

Safford, H.R., Shapiro, K., Bischel, H.N., 2022. Wastewater analysis can be a powerful public health tool—if it’s done sensibly. Proceedings of the National Academy of Sciences 119, e2119600119.

Scott, L.C., Aubee, A., Babahaji, L., Vigil, K., Tims, S., Aw, T.G., 2021. Targeted wastewater surveillance of SARS-CoV-2 on a university campus for COVID-19 outbreak detection and mitigation. Environ Res 200, 111374.

Sherchan, S.P., Shahin, S., Ward, L.M., Tandukar, S., Aw, T.G., Schmitz, B., Ahmed, W., Kitajima, M., 2020. First detection of SARS-CoV-2 RNA in wastewater in North America: a study in Louisiana, USA. Science of The Total Environment 743, 140621.

Sims, N., Kasprzyk-Hordern, B., 2020. Future perspectives of wastewater-based epidemiology: monitoring infectious disease spread and resistance to the community level. Environ Int 139, 105689.

Tada, T., Zhou, H., Dcosta, B.M., Samanovic, M.I., Chivukula, V., Herati, R.S., Hubbard, S.R., Mulligan, M.J., Landau, N.R., 2022. Increased resistance of SARS-CoV-2 Omicron variant to neutralization by vaccine-elicited and therapeutic antibodies. EBioMedicine 78, 103944.

Treibel, T.A., Manisty, C., Burton, M., McKnight, Á., Lambourne, J., Augusto, J.B., Couto-Parada, X., Cutino-Moguel, T., Noursadeghi, M., Moon, J.C., 2020. COVID-19: PCR screening of asymptomatic health-care workers at London hospital. The Lancet 395, 1608–1610.

Tyson, J.R., James, P., Stoddart, D., Sparks, N., Wickenhagen, A., Hall, G., Choi, J.H., Lapointe, H., Kamelian, K., Smith, A.D., 2020. Improvements to the ARTIC multiplex PCR method for SARS-CoV-2 genome sequencing using nanopore. BioRxiv.

Valieris, R., Drummond, R.D., Defelicibus, A., Dias-Neto, E., Rosales, R.A., Tojal da Silva, I., 2022. A mixture model for determining SARS-Cov-2 variant composition in pooled samples. Bioinformatics 38, 1809–1815.

Vo, V., Tillett, R.L., Chang, C.-L., Gerrity, D., Betancourt, W.Q., Oh, E.C., 2022. SARS-CoV-2 variant detection at a university dormitory using wastewater genomic tools. Science of The Total Environment 805, 149930.

Vogels, C.B.F., Breban, M.I., Ott, I.M., Alpert, T., Petrone, M.E., Watkins, A.E., Kalinich, C.C., Earnest, R., Rothman, J.E., Goes de Jesus, J., 2021. Multiplex qPCR discriminates variants of concern to enhance global surveillance of SARS-CoV-2. PLoS Biol 19, e3001236.

Wade, M.J., Jacomo, A. lo, Armenise, E., Brown, M.R., Bunce, J.T., Cameron, G.J., Fang, Z., Gilpin, D.F., Graham, D.W., Grimsley, J.M.S., 2022. Understanding and managing uncertainty and variability for wastewater monitoring beyond the pandemic: Lessons learned from the United Kingdom national COVID-19 surveillance programmes. J Hazard Mater 424, 127456.

Wan, Y., Shang, J., Graham, R., Baric, R.S., Li, F., 2020. Receptor recognition by the novel coronavirus from Wuhan: an analysis based on decade-long structural studies of SARS coronavirus. J Virol 94, e00127–20.

Wang, H., Miller, J.A., Verghese, M., Sibai, M., Solis, D., Mfuh, K.O., Jiang, B., Iwai, N., Mar, M., Huang, C., 2021. Multiplex SARS-CoV-2 genotyping PCR for population-level variant screening and epidemiologic surveillance. medRxiv.

Wang, Y., Liu, P., Zhang, H., Ibaraki, M., VanTassell, J., Geith, K., Cavallo, M., Kann, R., Saber, L., Kraft, C.S., 2022. Early warning of a COVID-19 surge on a university campus based on wastewater surveillance for SARS-CoV-2 at residence halls. Science of The Total Environment 821, 153291.

Wilhelm, A., Widera, M., Grikscheit, K., Toptan, T., Schenk, B., Pallas, C., Metzler, M., Kohmer, N., Hoehl, S., Helfritz, F.A., 2021. Reduced neutralization of SARS-CoV-2 Omicron variant by vaccine sera and monoclonal antibodies. MedRxiv.

Wolfe, M., Hughes, B., Duong, D., Chan-Herur, V., Wigginton, K.R., White, B.J., Boehm, A.B., 2022. Detection of SARS-CoV-2 Variants Mu, Beta, Gamma, Lambda, Delta, Alpha, and Omicron in Wastewater Settled Solids Using Mutation-Specific Assays Is Associated with Regional Detection of Variants in Clinical Samples. Appl Environ Microbiol 88, e00045–22.

Wölfel, R., Corman, V.M., Guggemos, W., Seilmaier, M., Zange, S., Müller, M.A., Niemeyer, D., Jones, T.C., Vollmar, P., Rothe, C., 2020. Author Correction: Virological assessment of hospitalized patients with COVID-2019. Nature 588, E35–E35.

Wolter, N., Jassat, W., Walaza, S., Welch, R., Moultrie, H., Groome, M., Amoako, D.G., Everatt, J., Bhiman, J.N., Scheepers, C., 2022. Early assessment of the clinical severity of the SARS-CoV-2 omicron variant in South Africa: a data linkage study. The Lancet 399, 437–446.

Wu, F., Xiao, A., Zhang, J., Moniz, K., Endo, N., Armas, F., Bonneau, R., Brown, M.A., Bushman, M., Chai, P.R., 2022. SARS-CoV-2 RNA concentrations in wastewater foreshadow dynamics and clinical presentation of new COVID-19 cases. Science of The Total Environment 805, 150121.

Wurtz, N., Penant, G., Jardot, P., Duclos, N., la Scola, B., 2021. Culture of SARS-CoV-2 in a panel of laboratory cell lines, permissivity, and differences in growth profile. European Journal of Clinical Microbiology & Infectious Diseases 40, 477–484.

Xagoraraki, I., O’Brien, E., 2020. Wastewater-based epidemiology for early detection of viral outbreaks, in: Women in Water Quality. Springer, pp. 75–97.

Zahmatkesh, S., Sillanpaa, M., Rezakhani, Y., Wang, C., 2022. Review of Concerned SARS-CoV-2 Variants Like Alpha (B. 1.1. 7), Beta (B. 1.351), Gamma (P. 1), Delta (B. 1.617. 2), and Omicron (B. 1.1. 529), as well as Novel Methods for Reducing and Inactivating SARS-CoV-2 Mutants in Wastewater Treatment Facilities. Journal of Hazardous Materials Advances 100140.

Zhu, N., Zhang, D., Wang, W., Li, X., Yang, B., Song, J., Zhao, X., Huang, B., Shi, W., Lu, R., 2020. A novel coronavirus from patients with pneumonia in China, 2019. New England journal of medicine.

