## Supplementary Information for "Campus Sewage Water Surveillance based dynamics and infection trends of SARS-CoV-2 variants during third wave of COVID-19 in Pune, India"

**Supplementary Table 1:** Details of sewage water sampling, qRT-PCR results (N, RdRp and E gene of SARS-CoV-2 and human control as internal control) and NGS data. Raw reads were quality checked and filtered using *fastp.* **HC**: Human control; **Ct:** Cycle threshold; **Ref**: Reference genome of SARS-CoV-2 (MN908947.3)

| **Sam-ple ID** | **Samp-ling Date** | **qRT-PCR Result** | **Ct value** | | | | **Viral load (copies/ml)** | **Genome Cover-age** | **Total No of Reads Million (M)** | **Reads After Filtra-tion (M)** | **Reads mapped to Ref. genome** | **% of reads mapped to Ref. genome** |
| --- | --- | --- | --- | --- | --- | --- | --- | --- | --- | --- | --- | --- |
|  |  |  | **N** | **RdRp** | **E** | **HC** |  |  |  |  |  |  |
| PR-39 | 08-Nov-21 | Negative | 0 | 0 | 38.21 | 31.94 | 0.4256 | 88.57 | 5.611624 | 5.434132 | 1413197 | 26.00 |
| PR-40 | 15-Nov-21 | Negative | 38.35 | 38.64 | 37.6 | 34.62 | 2.912 | 88.79 | 5.338325 | 5.186424 | 1389433 | 26.78 |
| PR-41 | 22-Nov-21 | Positive | 29.23 | 30.55 | 30.92 | 34.52 | 111.44 | 95.48 | 1.443459 | 1.365927 | 247969 | 18.14 |
| PR-42 | 29-Nov-21 | Negative | 32.55 | 38.25 | 36.82 | 32.71 | 2.828 | 94.64 | 5.844010 | 5.515828 | 1780886 | 32.25 |
| PR-43 | 06-Dec-21 | Negative | 31.46 | 35.75 | 36.73 | 31.39 | 4.564 | 57.2 | 5.133580 | 4.772402 | 1249018 | 26.15 |
| PR-44 | 13-Dec-21 | Negative | 35.49 | 35.86 | 37.78 | 32.16 | 1.7836 | 76.96 | 4.388206 | 4.057792 | 655990 | 16.16 |
| PR-45 | 16-Dec-21 | Negative | 35.89 | 40.28 | 39.32 | 36.35 | 0.49 | 75.23 | 5.079211 | 4.703773 | 806302 | 17.14 |
| PR-46 | 20-Dec-21 | Negative | 35.39 | 36.48 | 36.46 | 31.65 | 1.8928 | 78.78 | 5.038525 | 4.606785 | 761892 | 16.53 |
| PR-47 | 23-Dec-21 | Negative | 0 | 0 | 0 | 34.33 | 0 | 91.83 | 4.825359 | 4.687523 | 1316150 | 28.07 |
| PR-48 | 27-Dec-21 | Negative | 38.76 | 0 | 0 | 35.54 | 0.0673 | 88.2 | 5.139689 | 4.991671 | 1382783 | 27.69 |
| PR-49 | 30-Dec-21 | Negative | 37.61 | 39.07 | 0 | 33.87 | 0.0915 | 95.22 | 7.935566 | 7.531358 | 1165267 | 15.47 |
| PR-50 | 03-Jan-22 | Positive | 30.24 | 32.75 | 33.12 | 33.69 | 23.044 | 96.69 | 1.280912 | 1.199468 | 209227 | 17.44 |
| PR-51 | 06-Jan-22 | Positive | 29.65 | 32.4 | 32.89 | 33.58 | 51.52 | 98.93 | 4.774020 | 4.598473 | 741246 | 16.11 |
| PR-52 | 10-Jan-22 | Positive | 28.86 | 31.45 | 31.09 | 33.32 | 136.85 | 98.42 | 5.101415 | 4.927306 | 960690 | 19.48 |
| PR-53 | 13-Jan-22 | Positive | 26.54 | 28.94 | 28.99 | 31.04 | 277 | 99.76 | 4.397424 | 4.276766 | 1865555 | 43.60 |
| PR-54 | 17-Jan-22 | Positive | 25.46 | 28.46 | 28.71 | 28.63 | 275 | 97.81 | 3.339500 | 3.163125 | 954837 | 30.18 |
| PR-55 | 20-Jan-22 | Positive | 25.15 | 27.83 | 26.78 | 29.37 | 526 | 98.75 | 3.643429 | 3.518642 | 1494532 | 42.47 |
| PR-56 | 24-Jan-22 | Positive | 27.78 | 29.66 | 28.81 | 33.57 | 256 | 98.46 | 4.039068 | 3.877571 | 958619 | 24.72 |
| PR-57 | 28-Jan-22 | Positive | 29.3 | 30.63 | 29.75 | 31.32 | 132 | 99.72 | 3.885318 | 3.846160 | 2255788 | 58.64 |
| PR-58 | 31-Jan-22 | Positive | 28.27 | 32.84 | 31.4 | 28.62 | 58.4 | 97.87 | 3.258752 | 3.099596 | 770457 | 24.85 |
| PR-59 | 03-Feb-22 | Positive | 26.96 | 31.97 | 30.58 | 30.45 | 110 | 98.82 | 2.544216 | 2.456155 | 923109 | 37.57 |
| PR-60 | 07-Feb-22 | Positive | 30.82 | 33.39 | 32.5 | 33.39 | 44.6 | 85.74 | 2.305476 | 2.213425 | 512624 | 23.15 |
| PR-61 | 10-Feb-22 | Positive | 30.96 | 34.81 | 33.36 | 26.51 | 26 | 88.27 | 2.330473 | 2.224737 | 453979 | 20.39 |
| PR-62 | 14-Feb-22 | Positive | 26.82 | 28.95 | 27.11 | 31.12 | 229 | 98.03 | 3.896496 | 3.749742 | 970654 | 25.87 |
| PR-63 | 18-Feb-22 | Positive | 28.8 | 31.34 | 29.57 | 31.24 | 49.6 | 63.96 | 2.074386 | 1.971915 | 130208 | 6.60 |
| PR-64 | 21-Feb-22 | Positive | 30.77 | 34.44 | 33.31 | 31.31 | 22.6 | 94.16 | 1.499933 | 1.416394 | 197793 | 13.96 |
| PR-65 | 25-Feb-22 | Positive | 32.07 | 35.96 | 34.28 | 30.3 | 5.87 | 72.14 | 1.008602 | 0.9441 | 146858 | 15.55 |
| PR-66 | 28-Feb-22 | Positive | 33.67 | 36.05 | 34.98 | 31.34 | 4.91 | 37 | 1.633056 | 1.537163 | 39411 | 2.56 |
| PR-67 | 03-Mar-22 | Positive | 30.18 | 32.33 | 30.42 | 30.43 | 16.7 | 50 | 1.569485 | 1.462009 | 89679 | 6.13 |
| PR-68 | 07-Mar-22 | Negative | 34.29 | 37.76 | 36.38 | 31.91 | 4.43 | 18 | 1.780636 | 1.720377 | 40824 | 2.37 |
| PR-69 | 10-Mar-22 | Negative | 34.13 | 37.84 | 35.82 | 31.75 | 5.03 | 21 | 1.797309 | 1.684330 | 77917 | 4.62 |
| PR-70 | 14-Mar-22 | Negative | 35.9 | 40.02 | 38.85 | 29.54 | 0.621 | 20 | 2.124956 | 1.984329 | 73319 | 3.69 |
| PR-71 | 21-Mar-22 | Negative | 36.93 | 0 | 38.55 | 29.07 | 0.843 | 16 | 2.419227 | 2.358400 | 70829 | 3.00 |
| PR-72 | 28-Mar-22 | Negative | 38 | 40.59 | 39.39 | 29.98 | 1.24 | 19 | 1.165791 | 1.111143 | 53384 | 4.80 |
| PR-73 | 04-Apr-22 | Negative | 36.77 | 0 | 0 | 32.65 | 0.0303 | 18 | 2.018268 | 1.917459 | 93627 | 4.88 |
| PR-74 | 11-Apr-22 | Negative | 35.5 | 0 | 0 | 29.93 | 1.02 | 20 | 1.004969 | 0.9265 | 34353 | 3.57 |
| PR-75 | 18-Apr-22 | Negative | 38.16 | 0 | 40 | 30.48 | 0.608 | 28 | 1.116263 | 1.022465 | 34981 | 3.42 |
| PR-76 | 25-Apr-22 | Negative | 39.67 | 0 | 40.25 | 34.78 | 0.0925 | 46 | 2.373484 | 2.120135 | 144788 | 6.83 |

**Supplementary Table 2:** Clinical Data comprehending date and number of cases reported on campus

| **S. No.** | **Case** | **Reporting date** |
| --- | --- | --- |
| 1 | Case 1 | 06-12-2021 |
| 2 | Case 2 | 25-12-2021 |
| 3 | Case 3 | 04-01-2022 |
| 4 | Case 4 | 05-01-2022 |
| 5 | Case 5 | 05-01-2022 |
| 6 | Case 6 | 06-01-2022 |
| 7 | Case 7 | 07-01-2022 |
| 8 | Case 8 | 07-01-2022 |
| 9 | Case 9 | 08-01-2022 |
| 10 | Case 10 | 08-01-2022 |
| 11 | Case 11 | 08-01-2022 |
| 12 | Case 12 | 08-01-2022 |
| 13 | Case 13 | 08-01-2022 |
| 14 | Case 14 | 08-01-2022 |
| 15 | Case 15 | 08-01-2022 |
| 16 | Case 16 | 08-01-2022 |
| 17 | Case 17 | 08-01-2022 |
| 18 | Case 18 | 09-01-2022 |
| 19 | Case 19 | 09-01-2022 |
| 20 | Case 20 | 09-01-2022 |
| 21 | Case 21 | 09-01-2022 |
| 22 | Case 22 | 11-01-2022 |
| 23 | Case 23 | 11-01-2022 |
| 24 | Case 24 | 11-01-2022 |
| 25 | Case 25 | 11-01-2022 |
| 26 | Case 26 | 11-01-2022 |
| 27 | Case 27 | 12-01-2022 |
| 28 | Case 28 | 12-01-2022 |
| 29 | Case 29 | 12-01-2022 |
| 30 | Case 30 | 12-01-2022 |
| 31 | Case 31 | 12-01-2022 |
| 32 | Case 32 | 12-01-2022 |
| 33 | Case 33 | 12-01-2022 |
| 34 | Case 34 | 12-01-2022 |
| 35 | Case 35 | 13-01-2022 |
| 36 | Case 36 | 13-01-2022 |
| 37 | Case 37 | 13-01-2022 |
| 38 | Case 38 | 13-01-2022 |
| 39 | Case 39 | 13-01-2022 |
| 40 | Case 40 | 13-01-2022 |
| 41 | Case 41 | 13-01-2022 |
| 42 | Case 42 | 13-01-2022 |
| 43 | Case 43 | 13-01-2022 |
| 44 | Case 44 | 13-01-2022 |
| 45 | Case 45 | 13-01-2022 |
| 46 | Case 46 | 13-01-2022 |
| 47 | Case 47 | 13-01-2022 |
| 48 | Case 48 | 14-01-2022 |
| 49 | Case 49 | 14-01-2022 |
| 50 | Case 50 | 14-01-2022 |
| 51 | Case 51 | 14-01-2022 |
| 52 | Case 52 | 14-01-2022 |
| 53 | Case 53 | 14-01-2022 |
| 54 | Case 54 | 14-01-2022 |
| 55 | Case 55 | 14-01-2022 |
| 56 | Case 56 | 14-01-2022 |
| 57 | Case 57 | 14-01-2022 |
| 58 | Case 58 | 14-01-2022 |
| 59 | Case 59 | 14-01-2022 |
| 60 | Case 60 | 14-01-2022 |
| 61 | Case 61 | 14-01-2022 |
| 62 | Case 62 | 14-01-2022 |
| 63 | Case 63 | 14-01-2022 |
| 64 | Case 64 | 15-01-2022 |
| 65 | Case 65 | 17-01-2022 |
| 66 | Case 66 | 17-01-2022 |
| 67 | Case 67 | 17-01-2022 |
| 68 | Case 68 | 17-01-2022 |
| 69 | Case 69 | 17-01-2022 |
| 70 | Case 70 | 17-01-2022 |
| 71 | Case 71 | 17-01-2022 |
| 72 | Case 72 | 17-01-2022 |
| 73 | Case 73 | 17-01-2022 |
| 74 | Case 74 | 17-01-2022 |
| 75 | Case 75 | 17-01-2022 |
| 76 | Case 76 | 17-01-2022 |
| 77 | Case 77 | 17-01-2022 |
| 78 | Case 78 | 17-01-2022 |
| 79 | Case 79 | 17-01-2022 |
| 80 | Case 80 | 17-01-2022 |
| 81 | Case 81 | 17-01-2022 |
| 82 | Case 82 | 17-01-2022 |
| 83 | Case 83 | 17-01-2022 |
| 84 | Case 84 | 17-01-2022 |
| 85 | Case 85 | 17-01-2022 |
| 86 | Case 86 | 17-01-2022 |
| 87 | Case 87 | 17-01-2022 |
| 88 | Case 88 | 17-01-2022 |
| 89 | Case 89 | 17-01-2022 |
| 90 | Case 90 | 17-01-2022 |
| 91 | Case 91 | 17-01-2022 |
| 92 | Case 92 | 17-01-2022 |
| 93 | Case 93 | 17-01-2022 |
| 94 | Case 94 | 17-01-2022 |
| 95 | Case 95 | 17-01-2022 |
| 96 | Case 96 | 17-01-2022 |
| 97 | Case 97 | 17-01-2022 |
| 98 | Case 98 | 17-01-2022 |
| 99 | Case 99 | 17-01-2022 |
| 100 | Case 100 | 18-01-2022 |
| 101 | Case 101 | 18-01-2022 |
| 102 | Case 102 | 18-01-2022 |
| 103 | Case 103 | 18-01-2022 |
| 104 | Case 104 | 18-01-2022 |
| 105 | Case 105 | 18-01-2022 |
| 106 | Case 106 | 19-01-2022 |
| 107 | Case 107 | 19-01-2022 |
| 108 | Case 108 | 19-01-2022 |
| 109 | Case 109 | 19-01-2022 |
| 110 | Case 110 | 19-01-2022 |
| 111 | Case 111 | 19-01-2022 |
| 112 | Case 112 | 19-01-2022 |
| 113 | Case 113 | 19-01-2022 |
| 114 | Case 114 | 19-01-2022 |
| 115 | Case 115 | 19-01-2022 |
| 116 | Case 116 | 19-01-2022 |
| 117 | Case 117 | 19-01-2022 |
| 118 | Case 118 | 19-01-2022 |
| 119 | Case 119 | 19-01-2022 |
| 120 | Case 120 | 19-01-2022 |
| 121 | Case 121 | 19-01-2022 |
| 122 | Case 122 | 19-01-2022 |
| 123 | Case 123 | 20-01-2022 |
| 124 | Case 124 | 20-01-2022 |
| 125 | Case 125 | 20-01-2022 |
| 126 | Case 126 | 20-01-2022 |
| 127 | Case 127 | 20-01-2022 |
| 128 | Case 128 | 20-01-2022 |
| 129 | Case 129 | 20-01-2022 |
| 130 | Case 130 | 20-01-2022 |
| 131 | Case 131 | 21-01-2022 |
| 132 | Case 132 | 21-01-2022 |
| 133 | Case 133 | 21-01-2022 |
| 134 | Case 134 | 21-01-2022 |
| 135 | Case 135 | 21-01-2022 |
| 136 | Case 136 | 21-01-2022 |
| 137 | Case 137 | 21-01-2022 |
| 138 | Case 138 | 21-01-2022 |
| 139 | Case 139 | 24-01-2022 |
| 140 | Case 140 | 24-01-2022 |
| 141 | Case 141 | 24-01-2022 |
| 142 | Case 142 | 24-01-2022 |
| 143 | Case 143 | 24-01-2022 |
| 144 | Case 144 | 24-01-2022 |
| 145 | Case 145 | 24-01-2022 |
| 146 | Case 146 | 24-01-2022 |
| 147 | Case 147 | 24-01-2022 |
| 148 | Case 148 | 24-01-2022 |
| 149 | Case 149 | 24-01-2022 |
| 150 | Case 150 | 24-01-2022 |
| 151 | Case 151 | 24-01-2022 |
| 152 | Case 152 | 25-01-2022 |
| 153 | Case 153 | 25-01-2022 |
| 154 | Case 154 | 27-01-2022 |
| 155 | Case 155 | 27-01-2022 |
| 156 | Case 156 | 27-01-2022 |
| 157 | Case 157 | 27-01-2022 |
| 158 | Case 158 | 27-01-2022 |
| 159 | Case 159 | 27-01-2022 |
| 160 | Case 160 | 27-01-2022 |
| 161 | Case 161 | 27-01-2022 |
| 162 | Case 162 | 27-01-2022 |
| 163 | Case 163 | 27-01-2022 |
| 164 | Case 164 | 27-01-2022 |
| 165 | Case 165 | 27-01-2022 |
| 166 | Case 166 | 27-01-2022 |
| 167 | Case 167 | 27-01-2022 |
| 168 | Case 168 | 27-01-2022 |
| 169 | Case 169 | 27-01-2022 |
| 170 | Case 170 | 28-01-2022 |
| 171 | Case 171 | 28-01-2022 |
| 172 | Case 172 | 31-01-2022 |
| 173 | Case 173 | 31-01-2022 |
| 174 | Case 174 | 01-02-2022 |
| 175 | Case 175 | 02-02-2022 |
| 176 | Case 176 | 02-02-2022 |
| 177 | Case 177 | 04-02-2022 |
| 178 | Case 178 | 04-02-2022 |
| 179 | Case 179 | 04-02-2022 |
| 180 | Case 180 | 07-02-2022 |
| 181 | Case 181 | 27-02-2022 |
| 182 | Case 182 | 27-02-2022 |
| 183 | Case 183 | 27-02-2022 |
| 184 | Case 184 | 27-02-2022 |

**Supplementary Table 3:** Non-synonymous and synonymous nucleotide substitutions and deletionsdetected in the spike glycoprotein region as compared to the SARS-CoV-2 reference genome (MN908947.3). Referenceand alternative depth relateto the percentage of the total depth that corresponded to the nucleotide present in the reference genome and the alternative nucleotide, respectively.

| **Position** | **Amino Acid change** | **Reference depth** | **Alternative depth** | **Sample ID** |
| --- | --- | --- | --- | --- |
| **21575** | S:L5F | 18 | 1 | PR-59 |
|  |  | 45 | 2 | PR-61 |
| **21586** | S:L8L | 22 | 1 | PR-50 |
| **21588** | S:P9L | 10 | 1 | PR-65 |
| **21617** | S:T19S | 31 | 1 | PR-54 |
| **21618** | S:T19R | 130 | 129 | PR-45 |
|  |  | 2 | 2 | PR-48 |
|  |  | 236 | 79 | PR-52 |
|  |  | 51 | 51 | PR-41 |
|  | S:T19I | 236 | 152 | PR-52 |
|  |  | 115 | 53 | PR-53 |
|  |  | 32 | 1 | PR-54 |
|  |  | 493 | 445 | PR-57 |
|  |  | 147 | 44 | PR-56 |
|  |  | 51 | 51 | PR-55 |
|  |  | 8 | 8 | PR-59 |
|  |  | 82 | 82 | PR-62 |
|  |  | 57 | 57 | PR-61 |
|  |  | 43 | 43 | PR-58 |
|  |  | 12 | 12 | PR-65 |
|  |  | 44 | 44 | PR-64 |
| **21623** | S:R21G | 12 | 1 | PR-50 |
| **21632** | S:DEL:21632:10 | 239 | 0 | PR-52 |
|  |  | 117 | 0 | PR-53 |
|  |  | 485 | 0 | PR-57 |
|  |  | 177 | 0 | PR-56 |
|  |  | 48 | 0 | PR-55 |
|  |  | 6 | 0 | PR-59 |
|  |  | 77 | 0 | PR-62 |
|  |  | 58 | 0 | PR-61 |
|  |  | 39 | 0 | PR-58 |
|  |  | 14 | 0 | PR-65 |
|  |  | 45 | 0 | PR-64 |
| **21633** | S:L24S | 107 | 4 | PR-52 |
|  |  | 59 | 7 | PR-57 |
|  |  | 1 | 1 | PR-62 |
|  |  | 2 | 2 | PR-61 |
|  |  | 1 | 1 | PR-58 |
|  |  | 1 | 1 | PR-64 |
| **21635** | S:P25T | 16 | 1 | PR-51 |
| **21643** | S:A27A | 12 | 1 | PR-51 |
| **21664** | S:R34R | 2 | 1 | PR-60 |
|  |  | 20 | 1 | PR-59 |
| **21665** | S:G35C | 26 | 1 | PR-51 |
|  | S:G35S | 30 | 2 | PR-59 |
| **21667** | S:G35G | 50 | 3 | PR-50 |
|  |  | 313 | 21 | PR-55 |
|  |  | 1104 | 35 | PR-59 |
|  |  | 334 | 11 | PR-62 |
|  |  | 218 | 11 | PR-58 |
|  |  | 17 | 4 | PR-67 |
|  |  | 10 | 1 | PR-63 |
| **21682** | S:DEL:21682:2 | 41 | 0 | PR-63 |
| **21741** | S:S60C | 17 | 1 | PR-65 |
| **21745** | S:N61N | 27 | 1 | PR-64 |
|  |  | 8 | 3 | PR-49 |
| **21762** | S:A67V | 8 | 3 | PR-49 |
|  |  | 177 | 17 | PR-52 |
|  |  | 91 | 36 | PR-51 |
|  |  | 5140 | 1067 | PR-53 |
|  |  | 2645 | 455 | PR-54 |
|  |  | 93 | 43 | PR-50 |
|  |  | 426 | 29 | PR-56 |
|  |  | 1039 | 59 | PR-55 |
|  |  | 1 | 1 | PR-73 |
| **21764** | S:DEL:21764:7 | 8 | 0 | PR-49 |
|  |  | 175 | 0 | PR-52 |
|  |  | 92 | 0 | PR-51 |
|  |  | 5281 | 4 | PR-53 |
|  |  | 2753 | 1 | PR-54 |
|  |  | 96 | 0 | PR-50 |
|  |  | 461 | 0 | PR-56 |
|  |  | 1113 | 0 | PR-55 |
|  |  | 1 | 0 | PR-73 |
| **21846** | S:T95I | 15 | 8 | PR-49 |
|  |  | 179 | 31 | PR-52 |
|  |  | 115 | 78 | PR-51 |
|  |  | 6407 | 2222 | PR-53 |
|  |  | 3076 | 897 | PR-54 |
|  |  | 102 | 54 | PR-50 |
|  |  | 447 | 54 | PR-56 |
|  |  | 1220 | 143 | PR-55 |
|  |  | 4 | 4 | PR-73 |
|  |  | 14 | 2 | PR-69 |
| **21850** | S:E96E | 29 | 1 | PR-64 |
| **21863** | S:I101L | 14 | 1 | PR-69 |
| **21868** | S:R102R | 1106 | 131 | PR-62 |
| **21870** | S:G103V | 486 | 37 | PR-56 |
| **21895** | S:D111D | 3003 | 155 | PR-54 |
| **21920** | S:DEL:21920:2 | 11 | 0 | PR-49 |
| **21924** | S:N121I | 11 | 1 | PR-49 |
| **21945** | S:I128N | 32 | 1 | PR-64 |
| **21947** | S:K129* | 31 | 1 | PR-64 |
| **21952** | S:V130V | 33 | 1 | PR-64 |
| **21954** | S:C131F | 2109 | 64 | PR-54 |
|  |  | 706 | 63 | PR-58 |
| **21980** | S:F140I | 17 | 1 | PR-40 |
| **21986** | S:DEL:21986:10 | 6 | 0 | PR-49 |
|  |  | 128 | 0 | PR-52 |
|  |  | 104 | 0 | PR-51 |
|  |  | 4055 | 2 | PR-53 |
|  |  | 1949 | 1 | PR-54 |
|  |  | 66 | 1 | PR-50 |
|  |  | 383 | 0 | PR-56 |
|  |  | 1054 | 0 | PR-55 |
|  |  | 2 | 0 | PR-73 |
|  | S:G142C | 11 | 1 | PR-69 |
| **21987** | S:G142D | 81 | 77 | PR-47 |
|  |  | 2 | 2 | PR-49 |
|  |  | 107 | 98 | PR-52 |
|  |  | 79 | 45 | PR-51 |
|  |  | 2821 | 2812 | PR-53 |
|  |  | 1466 | 1462 | PR-54 |
|  |  | 39 | 39 | PR-50 |
|  |  | 1498 | 1358 | PR-57 |
|  |  | 348 | 337 | PR-56 |
|  |  | 992 | 927 | PR-55 |
|  |  | 1719 | 1715 | PR-60 |
|  |  | 3584 | 3573 | PR-59 |
|  |  | 841 | 837 | PR-62 |
|  |  | 237 | 194 | PR-61 |
|  |  | 707 | 703 | PR-58 |
|  |  | 34 | 34 | PR-67 |
|  |  | 10 | 9 | PR-65 |
|  |  | 51 | 51 | PR-63 |
|  |  | 31 | 28 | PR-64 |
|  |  | 10 | 10 | PR-69 |
| **21990** | S:DEL:21990:4 | 21 | 0 | PR-40 |
|  |  | 102 | 0 | PR-52 |
|  |  | 84 | 0 | PR-51 |
|  |  | 5 | 0 | PR-41 |
|  | S:H146Q | 28 | 1 | PR-40 |
| **22016** | S:W152R | 275 | 58 | PR-42 |
| **22022** | UTR:22022 | 23 | 1 | PR-63 |
| **22028** | S:DEL:22028:7 | 284 | 0 | PR-42 |
|  |  | 26 | 0 | PR-47 |
|  |  | 34 | 0 | PR-40 |
|  |  | 47 | 0 | PR-52 |
|  |  | 75 | 0 | PR-51 |
|  |  | 9 | 0 | PR-41 |
|  |  | 2 | 0 | PR-39 |
| **22029** | S:E156G | 2 | 1 | PR-42 |
| **22038** | S:V159A | 19 | 1 | PR-52 |
|  |  | 29 | 2 | PR-56 |
|  |  | 128 | 5 | PR-55 |
| **22039** | S:V159V | 28 | 1 | PR-56 |
| **22061** | S:T167A | 30 | 1 | PR-40 |
| **22077** | S:S172Y | 16 | 1 | PR-53 |
| **22113** | S:DEL:22113:2 | 1 | 0 | PR-54 |
| **22152** | S:I197T | 24 | 1 | PR-41 |
| **22153** | S:I197I | 16 | 1 | PR-52 |
| **22156** | S:D198D | 33 | 1 | PR-56 |
| **22158** | S:G199V | 31 | 1 | PR-51 |
| **22170** | S:I203K | 248 | 8 | PR-57 |
| **22181** | S:H207N | 28 | 1 | PR-41 |
| **22187** | S:P209T | 81 | 9 | PR-55 |
| **22193** | S:DEL:22193:4 | 36 | 0 | PR-53 |
|  |  | 5 | 0 | PR-50 |
| **22194** | S:N211I | 17 | 2 | PR-53 |
| **22199** | S:V213L | 13 | 1 | PR-52 |
| **22200** | S:V213G | 12 | 1 | PR-52 |
|  |  | 48 | 29 | PR-51 |
|  |  | 29 | 15 | PR-53 |
|  |  | 1 | 1 | PR-54 |
|  |  | 5 | 4 | PR-50 |
|  |  | 250 | 250 | PR-57 |
|  |  | 28 | 28 | PR-56 |
|  |  | 94 | 94 | PR-55 |
|  |  | 1 | 1 | PR-60 |
|  |  | 8 | 8 | PR-59 |
|  |  | 8 | 8 | PR-59 |
|  |  | 2 | 2 | PR-62 |
|  |  | 45 | 45 | PR-61 |
|  |  | 11 | 11 | PR-58 |
|  |  | 1 | 1 | PR-64 |
| **22204** | UTR:22204 | 27 | 7 | PR-53 |
|  |  | 4 | 1 | PR-50 |
| **22208** | S:L216I | 9 | 8 | PR-39 |
| **22211** | S:P217T | 27 | 1 | PR-56 |
| **22215** | S:Q218P | 28 | 1 | PR-41 |
| **22218** | S:DEL:22218:2 | 48 | 0 | PR-51 |
|  |  | 4 | 0 | PR-50 |
| **22227** | S:A222V | 5 | 5 | PR-47 |
| **22227** | S:DEL:22227:2 | 13 | 0 | PR-52 |
| **22228** | S:A222A | 18 | 11 | PR-46 |
| **22262** | S:N234D | 307 | 14 | PR-42 |
|  |  | 27 | 1 | PR-41 |
| **22286** | S:L242F | 39 | 14 | PR-40 |
| **22289** | S:A243S | 28 | 1 | PR-56 |
| **22305** | S:DEL:22305:2 | 28 | 0 | PR-56 |
| **22322** | S:S254T | 16 | 1 | PR-41 |
| **22339** | S:T259T | 33 | 1 | PR-53 |
|  |  | 25 | 1 | PR-70 |
| **22340** | S:A260P | 10 | 1 | PR-49 |
|  |  | 2 | 1 | PR-41 |
| **22342** | S:A260A | 9 | 1 | PR-42 |
| **22344** | S:G261D | 11 | 6 | PR-55 |
| **22347** | S:A262G | 1 | 1 | PR-42 |
| **22438** | S:A292A | 1 | 1 | PR-52 |
| **22449** | S:L296P | 31 | 2 | PR-53 |
| **22492** | S:K310K | 10 | 1 | PR-62 |
| **22512** | S:N317S | 21 | 1 | PR-57 |
| **22522** | S:V320V | 17 | 1 | PR-62 |
| **22523** | S:Q321E | 16 | 1 | PR-56 |
|  |  | 23 | 1 | PR-55 |
|  | S:Q321* | 11 | 1 | PR-58 |
| **22578** | S:G339D | 89 | 20 | PR-49 |
|  |  | 2887 | 1321 | PR-52 |
|  |  | 1148 | 126 | PR-51 |
|  |  | 1342 | 1230 | PR-53 |
|  |  | 119 | 110 | PR-54 |
|  |  | 8 | 8 | PR-50 |
|  |  | 5276 | 5276 | PR-57 |
|  |  | 2025 | 2025 | PR-56 |
|  |  | 2839 | 2837 | PR-55 |
|  |  | 17 | 17 | PR-60 |
|  |  | 359 | 359 | PR-59 |
|  |  | 2228 | 2228 | PR-62 |
|  |  | 667 | 667 | PR-61 |
|  |  | 2068 | 2068 | PR-58 |
|  |  | 2 | 2 | PR-66 |
|  |  | 207 | 207 | PR-65 |
|  |  | 371 | 371 | PR-63 |
|  |  | 1007 | 1006 | PR-64 |
|  |  | 303 | 303 | PR-70 |
|  |  | 40 | 40 | PR-69 |
|  |  | 309 | 309 | PR-72 |
| **22582** | S:E340D | 17 | 1 | PR-60 |
| **22583** | S:DEL:22583:3 | 11 | 0 | PR-50 |
| **22590** | S:N343S | 747 | 61 | PR-61 |
| **22599** | S:R346I | 2863 | 311 | PR-52 |
|  | S:R346K | 8 | 1 | PR-50 |
| **22605** | S:A348E | 32 | 1 | PR-69 |
| **22661** | S:V367F | 3479 | 357 | PR-52 |
| **22673** | S:S371P | 149 | 12 | PR-54 |
|  |  | 10 | 6 | PR-50 |
| **22674** | S:S371F | 3484 | 1369 | PR-52 |
|  |  | 1458 | 143 | PR-51 |
|  |  | 1810 | 1632 | PR-53 |
|  |  | 152 | 137 | PR-54 |
|  |  | 10 | 6 | PR-50 |
|  |  | 6520 | 6519 | PR-57 |
|  |  | 2555 | 2555 | PR-56 |
|  |  | 3379 | 3378 | PR-55 |
|  |  | 23 | 23 | PR-60 |
|  |  | 365 | 365 | PR-59 |
|  |  | 365 | 365 | PR-59 |
|  |  | 2605 | 2605 | PR-62 |
|  |  | 765 | 765 | PR-61 |
|  |  | 2356 | 2356 | PR-58 |
|  |  | 206 | 206 | PR-65 |
|  |  | 345 | 345 | PR-63 |
|  |  | 1191 | 1191 | PR-64 |
|  |  | 344 | 344 | PR-70 |
|  |  | 28 | 28 | PR-69 |
|  |  | 376 | 376 | PR-72 |
| **22676** | S:A372S | 147 | 14 | PR-54 |
| **22677** | S:A372E | 227 | 72 | PR-39 |
| **22679** | S:S373P | 3497 | 1338 | PR-52 |
|  |  | 1449 | 145 | PR-51 |
|  |  | 1779 | 1603 | PR-53 |
|  |  | 150 | 136 | PR-54 |
|  |  | 13 | 7 | PR-50 |
|  |  | 6320 | 6318 | PR-57 |
|  |  | 2497 | 2497 | PR-56 |
|  |  | 3320 | 3320 | PR-55 |
|  |  | 22 | 22 | PR-60 |
|  |  | 366 | 366 | PR-59 |
|  |  | 366 | 366 | PR-59 |
|  |  | 2510 | 2509 | PR-62 |
|  |  | 763 | 763 | PR-61 |
|  |  | 2287 | 2287 | PR-58 |
|  |  | 207 | 206 | PR-65 |
|  |  | 349 | 349 | PR-63 |
|  |  | 1194 | 1190 | PR-64 |
|  |  | 326 | 325 | PR-70 |
|  |  | 26 | 26 | PR-69 |
|  |  | 377 | 377 | PR-72 |
| **22685** | S:DEL:22685:2 | 23 | 0 | PR-69 |
| **22686** | S:S375Y | 1531 | 258 | PR-51 |
|  | S:S375F | 3596 | 1352 | PR-52 |
|  |  | 1815 | 1625 | PR-53 |
|  |  | 158 | 145 | PR-54 |
|  |  | 14 | 7 | PR-50 |
|  |  | 6304 | 6302 | PR-57 |
|  |  | 2521 | 2521 | PR-56 |
|  |  | 3268 | 3266 | PR-55 |
|  |  | 25 | 25 | PR-60 |
|  |  | 383 | 383 | PR-59 |
|  |  | 2556 | 2554 | PR-62 |
|  |  | 793 | 792 | PR-61 |
|  |  | 2318 | 2317 | PR-58 |
|  |  | 187 | 187 | PR-65 |
|  |  | 335 | 335 | PR-63 |
|  |  | 1186 | 1186 | PR-64 |
|  |  | 334 | 334 | PR-70 |
|  |  | 24 | 24 | PR-69 |
|  |  | 380 | 380 | PR-72 |
| **22688** | S:T376A | 3529 | 1304 | PR-52 |
|  |  | 1514 | 139 | PR-51 |
|  |  | 1755 | 1571 | PR-53 |
|  |  | 151 | 119 | PR-54 |
|  |  | 6057 | 6054 | PR-57 |
|  |  | 2446 | 2441 | PR-56 |
|  |  | 3165 | 3165 | PR-55 |
|  |  | 25 | 25 | PR-60 |
|  |  | 365 | 364 | PR-59 |
|  |  | 2491 | 2488 | PR-62 |
|  |  | 777 | 776 | PR-61 |
|  |  | 2261 | 2260 | PR-58 |
|  |  | 184 | 184 | PR-65 |
|  |  | 326 | 326 | PR-63 |
|  |  | 1155 | 1154 | PR-64 |
|  |  | 321 | 321 | PR-70 |
|  |  | 26 | 26 | PR-69 |
|  |  | 372 | 372 | PR-72 |
| **22691** | S:F377L | 3712 | 413 | PR-52 |
| **22713** | S:P384H | 35 | 35 | PR-69 |
| **22770** | S:R403T | 7928 | 859 | PR-43 |
| **22775** | S:D405N | 2956 | 1221 | PR-52 |
|  |  | 1225 | 116 | PR-51 |
|  |  | 1479 | 1293 | PR-53 |
|  |  | 112 | 81 | PR-54 |
|  |  | 8 | 1 | PR-50 |
|  |  | 4800 | 4798 | PR-57 |
|  |  | 2060 | 2060 | PR-56 |
|  |  | 2477 | 2476 | PR-55 |
|  |  | 9 | 9 | PR-60 |
|  |  | 247 | 234 | PR-59 |
|  |  | 2304 | 2304 | PR-62 |
|  |  | 664 | 664 | PR-61 |
|  |  | 1943 | 1943 | PR-58 |
|  |  | 165 | 165 | PR-65 |
|  |  | 323 | 323 | PR-63 |
|  |  | 981 | 979 | PR-64 |
|  |  | 288 | 288 | PR-70 |
|  |  | 23 | 23 | PR-69 |
|  |  | 320 | 319 | PR-72 |
| **22786** | S:R408S | 2783 | 1133 | PR-52 |
|  |  | 1189 | 96 | PR-51 |
|  |  | 1438 | 1283 | PR-53 |
|  |  | 122 | 88 | PR-54 |
|  |  | 9 | 1 | PR-50 |
|  |  | 4518 | 4514 | PR-57 |
|  |  | 1945 | 1943 | PR-56 |
|  |  | 2321 | 2320 | PR-55 |
|  |  | 8 | 8 | PR-60 |
|  |  | 239 | 239 | PR-59 |
|  |  | 2128 | 2125 | PR-62 |
|  |  | 635 | 635 | PR-61 |
|  |  | 1880 | 1880 | PR-58 |
|  |  | 172 | 172 | PR-65 |
|  |  | 305 | 304 | PR-63 |
|  |  | 959 | 958 | PR-64 |
|  |  | 293 | 293 | PR-70 |
|  |  | 21 | 21 | PR-69 |
|  |  | 308 | 307 | PR-72 |
| **22794** | S:A411G | 221 | 59 | PR-39 |
| **22796** | S:P412T | 31 | 1 | PR-69 |
| **22813** | S:K417N | 3674 | 1509 | PR-52 |
|  |  | 1568 | 142 | PR-51 |
|  |  | 1936 | 1723 | PR-53 |
|  |  | 155 | 145 | PR-54 |
|  |  | 9 | 7 | PR-50 |
|  |  | 6579 | 6568 | PR-57 |
|  |  | 2694 | 2678 | PR-56 |
|  |  | 3349 | 3337 | PR-55 |
|  |  | 19 | 19 | PR-60 |
|  |  | 423 | 419 | PR-59 |
|  |  | 2727 | 2721 | PR-62 |
|  |  | 875 | 872 | PR-61 |
|  |  | 2516 | 2513 | PR-58 |
|  |  | 210 | 209 | PR-65 |
|  |  | 356 | 356 | PR-63 |
|  |  | 1206 | 1200 | PR-64 |
|  |  | 368 | 367 | PR-70 |
|  |  | 31 | 31 | PR-69 |
|  |  | 377 | 374 | PR-72 |
| **22817** | S:A419S | 2492 | 568 | PR-42 |
|  |  | 8518 | 2385 | PR-43 |
| **22844** | S:D428Y | 2906 | 220 | PR-58 |
| **22848** | S:F429Y | 31 | 1 | PR-60 |
| **22868** | S:W436G | 12 | 1 | PR-45 |
| **22882** | S:N440K | 233 | 12 | PR-59 |
| **22917** | S:L452R | 6 | 6 | PR-42 |
|  |  | 14 | 14 | PR-45 |
|  |  | 18 | 18 | PR-44 |
|  |  | 6 | 18 | PR-46 |
|  |  | 54 | 54 | PR-47 |
|  |  | 52 | 52 | PR-40 |
|  |  | 97 | 97 | PR-48 |
|  |  | 7 | 7 | PR-49 |
|  |  | 2 | 1 | PR-52 |
|  |  | 5 | 4 | PR-51 |
|  |  | 2 | 1 | PR-54 |
|  |  | 1 | 1 | PR-41 |
|  |  | 97 | 97 | PR-39 |
| **22933** | S:R457S | 84 | 5 | PR-48 |
| **22939** | S:S459S | 16 | 1 | PR-44 |
| **22984** | S:Q474H | 25 | 1 | PR-44 |
|  |  | 108 | 7 | PR-48 |
| **22988** | S:G476C | 6 | 1 | PR-46 |
|  |  | 109 | 5 | PR-39 |
| **22992** | S:S477N | 1 | 1 | PR-53 |
|  |  | 1 | 1 | PR-50 |
|  |  | 10 | 10 | PR-57 |
|  |  | 4 | 4 | PR-56 |
|  |  | 2 | 2 | PR-55 |
|  |  | 6 | 6 | PR-59 |
|  |  | 6 | 6 | PR-59 |
|  |  | 15 | 15 | PR-62 |
| **22995** | S:T478K | 2 | 2 | PR-42 |
|  |  | 11 | 11 | PR-45 |
|  |  | 21 | 21 | PR-44 |
|  |  | 13 | 6 | PR-46 |
|  |  | 46 | 46 | PR-47 |
|  |  | 81 | 81 | PR-40 |
|  |  | 93 | 93 | PR-48 |
|  |  | 7 | 7 | PR-49 |
|  |  | 2 | 2 | PR-52 |
|  |  | 5 | 5 | PR-51 |
|  |  | 1 | 1 | PR-53 |
|  |  | 1 | 1 | PR-50 |
|  |  | 9 | 9 | PR-57 |
|  |  | 4 | 4 | PR-56 |
|  |  | 1 | 1 | PR-55 |
|  |  | 6 | 6 | PR-59 |
|  |  | 15 | 15 | PR-62 |
|  |  | 1 | 1 | PR-41 |
|  |  | 91 | 91 | PR-39 |
| **23013** | S:E484A | 2 | 2 | PR-53 |
|  |  | 1 | 1 | PR-50 |
|  |  | 9 | 9 | PR-57 |
|  |  | 5 | 5 | PR-56 |
|  |  | 3 | 3 | PR-55 |
|  |  | 6 | 6 | PR-59 |
|  |  | 16 | 16 | PR-62 |
| **23025** | S:C488F | 13 | 2 | PR-46 |
| **23040** | S:Q493R | 1 | 1 | PR-53 |
|  |  | 9 | 9 | PR-57 |
|  |  | 5 | 5 | PR-56 |
|  |  | 2 | 2 | PR-55 |
|  |  | 7 | 7 | PR-59 |
|  |  | 18 | 18 | PR-62 |
|  |  | 2 | 2 | PR-58 |
| **23047** | S:DEL:23047:2 | 4 | 0 | PR-52 |
| **23055** | S:Q498R | 5 | 1 | PR-52 |
|  |  | 1 | 1 | PR-53 |
|  |  | 2 | 2 | PR-54 |
|  |  | 13 | 13 | PR-57 |
|  |  | 4 | 4 | PR-56 |
|  |  | 4 | 4 | PR-55 |
|  |  | 8 | 8 | PR-59 |
|  |  | 17 | 17 | PR-62 |
|  |  | 3 | 3 | PR-58 |
| **23058** | S:P499R | 2 | 2 | PR-41 |
| **23063** | S:N501Y | 1 | 1 | PR-53 |
|  |  | 2 | 2 | PR-54 |
|  |  | 13 | 13 | PR-57 |
|  |  | 3 | 3 | PR-56 |
|  |  | 3 | 3 | PR-55 |
|  |  | 7 | 7 | PR-59 |
|  |  | 12 | 12 | PR-62 |
|  |  | 3 | 3 | PR-58 |
| **23071** | S:V503V | 48 | 2 | PR-47 |
| **23075** | S:Y505H | 1 | 1 | PR-54 |
|  |  | 15 | 15 | PR-57 |
|  |  | 2 | 2 | PR-56 |
|  |  | 4 | 4 | PR-55 |
|  |  | 8 | 8 | PR-59 |
|  |  | 10 | 10 | PR-62 |
|  |  | 3 | 3 | PR-58 |
| **23078** | S:Q506K | 5 | 1 | PR-51 |
| **23093** | S:V511L | 5 | 1 | PR-55 |
| **23106** | S:F515S | 4 | 1 | PR-52 |
| **23121** | S:A520G | 4 | 1 | PR-52 |
| **23122** | S:A520A | 29 | 12 | PR-42 |
|  |  | 83 | 3 | PR-48 |
|  |  | 14 | 4 | PR-49 |
|  |  | 21 | 16 | PR-52 |
|  |  | 26 | 17 | PR-51 |
|  |  | 7 | 5 | PR-53 |
|  |  | 1 | 1 | PR-54 |
|  |  | 5 | 3 | PR-56 |
|  |  | 12 | 3 | PR-59 |
|  |  | 29 | 14 | PR-62 |
|  |  | 3 | 3 | PR-61 |
|  |  | 6 | 3 | PR-58 |
|  |  | 1 | 1 | PR-63 |
|  |  | 1 | 1 | PR-64 |
|  |  | 4 | 4 | PR-70 |
|  |  | 1 | 1 | PR-69 |
|  |  | 3 | 2 | PR-41 |
| **23126** | S:A522S | 3039 | 1 | PR-46 |
| **23127** | S:A522E | 21 | 1 | PR-44 |
| **23128** | S:A522A | 50 | 2 | PR-57 |
|  |  | 44 | 2 | PR-55 |
| **23202** | S:T547K | 761 | 203 | PR-49 |
|  |  | 7816 | 1797 | PR-52 |
|  |  | 5834 | 351 | PR-51 |
|  |  | 7212 | 4091 | PR-53 |
|  |  | 3409 | 2344 | PR-54 |
|  |  | 1359 | 1041 | PR-50 |
|  |  | 24249 | 1417 | PR-57 |
|  |  | 14018 | 2376 | PR-56 |
|  |  | 22987 | 1771 | PR-55 |
|  |  | 3500 | 134 | PR-59 |
|  |  | 11333 | 686 | PR-62 |
|  |  | 11601 | 622 | PR-58 |
|  |  | 3143 | 1090 | PR-65 |
|  |  | 749 | 747 | PR-69 |
| **23213** | S:V551F | 20949 | 632 | PR-57 |
| **23230** | S:DEL:23230:2 | 11745 | 1 | PR-62 |
| **23285** | S:A575S | 6698 | 264 | PR-53 |
|  |  | 1455 | 44 | PR-50 |
| **23294** | S:D578Y | 6576 | 419 | PR-51 |
| **23294** | S:D578H | 6576 | 288 | PR-51 |
| **23295** | S:D578G | 474 | 474 | PR-45 |
| **23298** | S:P579Q | 6476 | 502 | PR-51 |
| **23302** | S:Q580H | 1502 | 184 | PR-60 |
| **23371** | UTR:23371 | 263 | 14 | PR-39 |
| **23381** | S:Q607K | 8344 | 556 | PR-61 |
| **23387** | S:A609T | 500 | 500 | PR-45 |
| **23403** | S:D614G | 26524 | 26511 | PR-42 |
|  |  | 14301 | 14298 | PR-43 |
|  |  | 506 | 505 | PR-45 |
|  |  | 2446 | 3038 | PR-46 |
|  |  | 35 | 35 | PR-47 |
|  |  | 322 | 321 | PR-40 |
|  |  | 359 | 359 | PR-48 |
|  |  | 1184 | 1184 | PR-49 |
|  |  | 9373 | 9372 | PR-52 |
|  |  | 6552 | 6549 | PR-51 |
|  |  | 6862 | 6861 | PR-53 |
|  |  | 3291 | 3291 | PR-54 |
|  |  | 1460 | 1460 | PR-50 |
|  |  | 25325 | 25318 | PR-57 |
|  |  | 14467 | 14459 | PR-56 |
|  |  | 23963 | 23950 | PR-55 |
|  |  | 1706 | 1706 | PR-60 |
|  |  | 3010 | 3009 | PR-59 |
|  |  | 12206 | 12200 | PR-62 |
|  |  | 8380 | 8379 | PR-61 |
|  |  | 12603 | 12599 | PR-58 |
|  |  | 1026 | 1026 | PR-74 |
|  |  | 3218 | 3217 | PR-65 |
|  |  | 2708 | 2706 | PR-63 |
|  |  | 1213 | 1211 | PR-64 |
|  |  | 1885 | 1884 | PR-70 |
|  |  | 874 | 873 | PR-69 |
|  |  | 3476 | 3473 | PR-41 |
|  |  | 629 | 629 | PR-39 |
| **23415** | S:T618R | 27476 | 1618 | PR-42 |
|  |  | 1112 | 456 | PR-49 |
| **23419** | S:E619E | 28859 | 9311 | PR-42 |
| **23434** | S:I624I | 2817 | 270 | PR-41 |
| **23443** | S:D627D | 1 | 1 | PR-44 |
| **23466** | S:V635G | 19 | 1 | PR-67 |
|  |  | 28 | 1 | PR-72 |
| **23478** | S:G639V | 9393 | 352 | PR-52 |
| **23492** | S:Q644K | 28 | 1 | PR-67 |
| **23517** | S:G652A | 373 | 14 | PR-42 |
|  |  | 13 | 1 | PR-63 |
| **23525** | S:H655Y | 327 | 119 | PR-52 |
|  |  | 402 | 171 | PR-51 |
|  |  | 2022 | 1777 | PR-53 |
|  |  | 1246 | 883 | PR-54 |
|  |  | 36 | 5 | PR-50 |
|  |  | 474 | 474 | PR-57 |
|  |  | 551 | 551 | PR-56 |
|  |  | 438 | 438 | PR-55 |
|  |  | 1646 | 1645 | PR-60 |
|  |  | 1459 | 1458 | PR-59 |
|  |  | 353 | 353 | PR-62 |
|  |  | 95 | 95 | PR-61 |
|  |  | 763 | 763 | PR-58 |
|  |  | 44 | 43 | PR-67 |
|  |  | 45 | 45 | PR-64 |
|  |  | 26 | 26 | PR-69 |
|  |  | 76 | 76 | PR-72 |
|  |  | 4456 | 196 | PR-39 |
| **23547** | S:C662F | 380 | 16 | PR-51 |
| **23548** | S:C662W | 28 | 1 | PR-41 |
| **23551** | S:D663E | 30 | 1 | PR-50 |
| **23554** | S:DEL:23554:2 | 1248 | 0 | PR-54 |
|  | S:I664M | 24 | 1 | PR-69 |
| **23565** | S:A668E | 31 | 1 | PR-41 |
| **23587** | S:Q675H | 608 | 40 | PR-57 |
|  | S:Q675Q | 30 | 1 | PR-41 |
| **23591** | S:Q677K | 878 | 96 | PR-58 |
| **23599** | S:N679K | 319 | 124 | PR-52 |
|  |  | 384 | 165 | PR-51 |
|  |  | 2512 | 2236 | PR-53 |
|  |  | 1444 | 1053 | PR-54 |
|  |  | 37 | 2 | PR-50 |
|  |  | 585 | 585 | PR-57 |
|  |  | 639 | 639 | PR-56 |
|  |  | 559 | 559 | PR-55 |
|  |  | 1739 | 1738 | PR-60 |
|  |  | 1462 | 1462 | PR-59 |
|  |  | 401 | 401 | PR-62 |
|  |  | 80 | 80 | PR-61 |
|  |  | 864 | 864 | PR-58 |
|  |  | 64 | 64 | PR-67 |
|  |  | 56 | 56 | PR-64 |
|  |  | 21 | 21 | PR-69 |
|  |  | 103 | 103 | PR-72 |
|  |  | 4568 | 229 | PR-39 |
| **23604** | S:P681R | 200 | 200 | PR-42 |
|  |  | 2776 | 2776 | PR-43 |
|  |  | 2928 | 2928 | PR-45 |
|  |  | 1819 | 1819 | PR-44 |
|  |  | 60 | 2445 | PR-46 |
|  |  | 3866 | 3865 | PR-47 |
|  |  | 5817 | 5816 | PR-40 |
|  |  | 3809 | 3710 | PR-48 |
|  |  | 1595 | 1582 | PR-49 |
|  |  | 302 | 183 | PR-52 |
|  |  | 302 | 119 | PR-52 |
|  |  | 367 | 163 | PR-51 |
|  |  | 2426 | 2178 | PR-53 |
|  |  | 1381 | 1012 | PR-54 |
|  |  | 33 | 2 | PR-50 |
|  |  | 104 | 104 | PR-72 |
|  | S:P681H | 4451 | 233 | PR-39 |
|  |  | 367 | 204 | PR-51 |
|  |  | 2426 | 248 | PR-53 |
|  |  | 1381 | 368 | PR-54 |
|  |  | 33 | 30 | PR-50 |
|  |  | 560 | 560 | PR-57 |
|  |  | 612 | 612 | PR-56 |
|  |  | 540 | 539 | PR-55 |
|  |  | 1679 | 1679 | PR-60 |
|  |  | 1403 | 1402 | PR-59 |
|  |  | 386 | 386 | PR-62 |
|  |  | 77 | 77 | PR-61 |
|  |  | 820 | 820 | PR-58 |
|  |  | 61 | 61 | PR-67 |
|  |  | 54 | 54 | PR-64 |
|  |  | 20 | 20 | PR-69 |
|  | S:P681H | 27 | 27 | PR-41 |
|  | S:P681H | 4451 | 4218 | PR-39 |
| **23638** | S:I692I | 14 | 14 | PR-69 |
| **23725** | S:S721S | 30 | 1 | PR-69 |
| **23730** | S:T723I | 29 | 1 | PR-69 |
| **23760** | S:K733R | 4135 | 140 | PR-48 |
| **23767** | S:S735S | 3653 | 151 | PR-45 |
| **23780** | S:M740V | 33 | 1 | PR-50 |
| **23786** | S:I742F | 2 | 2 | PR-63 |
| **23801** | S:T747S | 22 | 1 | PR-65 |
| **23803** | S:T747T | 23 | 1 | PR-65 |
|  | S:DEL:23803:2 | 23 | 1 | PR-65 |
| **23809** | S:C749* | 14 | 1 | PR-63 |
| **23818** | S:L752L | 29 | 1 | PR-65 |
| **23835** | S:S758I | 154 | 24 | PR-52 |
| **23843** | S:T761A | 21 | 1 | PR-61 |
| **23846** | S:Q762K | 187 | 24 | PR-51 |
| **23848** | S:Q762H | 29 | 1 | PR-61 |
| **23854** | S:N764K | 107 | 26 | PR-52 |
|  |  | 182 | 62 | PR-51 |
|  |  | 645 | 605 | PR-53 |
|  |  | 332 | 315 | PR-54 |
|  |  | 51 | 51 | PR-50 |
|  |  | 723 | 722 | PR-57 |
|  |  | 209 | 209 | PR-56 |
|  |  | 291 | 291 | PR-55 |
|  |  | 10 | 10 | PR-60 |
|  |  | 56 | 56 | PR-59 |
|  |  | 81 | 81 | PR-62 |
|  |  | 27 | 27 | PR-61 |
|  |  | 140 | 140 | PR-58 |
|  |  | 17 | 17 | PR-67 |
|  |  | 40 | 40 | PR-65 |
|  |  | 22 | 22 | PR-63 |
|  |  | 95 | 95 | PR-64 |
|  |  | 23 | 12 | PR-39 |
| **23862** | S:L767* | 28 | 1 | PR-49 |
| **23864** | S:T768A | 21 | 1 | PR-63 |
| **23865** | S:T768I | 58 | 2 | PR-44 |
|  |  | 30 | 2 | PR-61 |
|  |  | 50 | 2 | PR-41 |
| **23873** | S:A771S | 165 | 6 | PR-51 |
| **23874** | S:A771D | 775 | 36 | PR-42 |
| **23876** | S:V772F | 759 | 36 | PR-42 |
| **23887** | S:D775E | 22 | 1 | PR-63 |
| **23893** | S:N777K | 64 | 33 | PR-44 |
| **23894** | S:T778P | 22 | 1 | PR-63 |
| **23899** | S:Q779Q | 8 | 1 | PR-40 |
| **23902** | S:E780D | 306 | 11 | PR-54 |
| **23908** | S:F782L | 20 | 1 | PR-63 |
| **23936** | S:P792T | 22 | 1 | PR-63 |
| **23937** | S:P792Q | 20 | 1 | PR-63 |
| **23939** | S:P793T | 22 | 1 | PR-63 |
| **23941** | S:P793P | 23 | 1 | PR-49 |
| **23948** | S:D796Y | 22 | 2 | PR-49 |
|  |  | 80 | 21 | PR-52 |
|  |  | 147 | 75 | PR-51 |
|  |  | 534 | 505 | PR-53 |
|  |  | 231 | 222 | PR-54 |
|  |  | 35 | 35 | PR-50 |
|  |  | 672 | 672 | PR-57 |
|  |  | 209 | 209 | PR-56 |
|  |  | 213 | 213 | PR-55 |
|  |  | 1 | 1 | PR-60 |
|  |  | 49 | 49 | PR-59 |
|  |  | 77 | 77 | PR-62 |
|  |  | 46 | 46 | PR-61 |
|  |  | 163 | 163 | PR-58 |
|  |  | 15 | 15 | PR-67 |
|  |  | 29 | 29 | PR-65 |
|  |  | 18 | 18 | PR-63 |
|  |  | 58 | 58 | PR-64 |
|  |  | 14 | 5 | PR-39 |
| **23955** | S:G798D | 74 | 9 | PR-52 |
|  |  | 494 | 34 | PR-53 |
|  |  | 75 | 32 | PR-62 |
|  |  | 46 | 24 | PR-61 |
|  |  | 149 | 38 | PR-58 |
| **23956** | S:G798G | 569 | 47 | PR-42 |
|  | S:DEL:23956:2 | 24 | 0 | PR-65 |
| **23973** | S:Q804P | 34 | 0 | PR-59 |
|  |  | 37 | 2 | PR-59 |
|  |  | 29 | 1 | PR-65 |
| **23975** | S:I805L | 20 | 1 | PR-63 |
| **23984** | S:D808Y | 29 | 1 | PR-65 |
| **23986** | S:D808D | 666 | 38 | PR-42 |
| **23987** | S:P809T | 7 | 3 | PR-40 |
| **23988** | S:P809R | 7 | 1 | PR-40 |
| **23998** | S:P812P | 22 | 1 | PR-63 |
| **24006** | S:R815M | 180 | 11 | PR-51 |
| **24019** | S:E819D | 57 | 2 | PR-41 |
| **24023** | S:L821I | 7 | 1 | PR-40 |
| **24028** | S:L822L | 24 | 1 | PR-63 |
| **24029** | S:F823I | 16 | 1 | PR-39 |
| **24031** | S:F823L | 23 | 1 | PR-67 |
| **24040** | S:V826V | 6 | 1 | PR-60 |
| **24048** | S:A829E | 11 | 1 | PR-40 |
| **24068** | S:Q836* | 26 | 1 | PR-63 |
| **24074** | S:G838S | 9 | 1 | PR-60 |
| **24075** | S:G838A | 6 | 1 | PR-45 |
|  |  | 6 | 1 | PR-47 |
| **24076** | S:G838G | 25 | 9 | PR-45 |
|  |  | 91 | 4 | PR-46 |
|  |  | 44 | 5 | PR-47 |
|  |  | 20 | 2 | PR-40 |
|  |  | 26 | 10 | PR-48 |
|  |  | 629 | 26 | PR-53 |
|  |  | 9 | 2 | PR-60 |
|  |  | 35 | 7 | PR-39 |
| **24077** | S:DEL:24077:2 | 140 | 0 | PR-45 |
|  |  | 83 | 0 | PR-44 |
|  |  | 82 | 0 | PR-48 |
|  |  | 33 | 0 | PR-49 |
|  |  | 81 | 0 | PR-39 |
| **24078** | S:D839V | 139 | 17 | PR-45 |
|  |  | 83 | 4 | PR-44 |
|  |  | 1313 | 5 | PR-46 |
|  |  | 139 | 19 | PR-47 |
|  |  | 75 | 9 | PR-40 |
|  |  | 83 | 10 | PR-48 |
|  |  | 83 | 3 | PR-48 |
|  |  | 32 | 1 | PR-49 |
|  |  | 800 | 30 | PR-53 |
|  |  | 44 | 2 | PR-60 |
|  |  | 23 | 1 | PR-63 |
|  |  | 81 | 11 | PR-39 |
| **24102** | S:R847I | 179 | 9 | PR-50 |
| **24127** | S:DEL:24127:20 | 402 | 0 | PR-62 |
| **24130** | S:N856K | 445 | 80 | PR-51 |
|  |  | 3358 | 1043 | PR-53 |
|  |  | 972 | 365 | PR-54 |
|  |  | 186 | 34 | PR-50 |
|  |  | 715 | 169 | PR-56 |
| **24140** | S:V860I | 21 | 1 | PR-65 |
| **24150** | S:P863L | 324 | 16 | PR-52 |
| **24151** | S:P863P | 4 | 1 | PR-67 |
| **24158** | S:T866S | 27 | 1 | PR-64 |
| **24160** | S:T866T | 20 | 1 | PR-64 |
| **24170** | S:I870V | 31 | 1 | PR-42 |
| **24172** | S:I870M | 2 | 2 | PR-42 |
| **24197** | S:A879P | 365 | 17 | PR-51 |
| **24202** | S:G880G | 595 | 101 | PR-58 |
| **24234** | S:G891V | 376 | 16 | PR-51 |
| **24237** | S:A892D | 1960 | 67 | PR-48 |
| **24241** | S:A893A | 22 | 1 | PR-64 |
| **24243** | S:L894S | 94 | 3 | PR-41 |
| **24244** | S:L894L | 2112 | 100 | PR-39 |
| **24305** | S:V915F | 419 | 20 | PR-56 |
| **24351** | S:A930V | 1028 | 37 | PR-39 |
| **24353** | S:I931F | 16 | 1 | PR-64 |
| **24377** | S:S939P | 1180 | 37 | PR-44 |
| **24400** | S:G946G | 12 | 1 | PR-43 |
| **24401** | S:K947E | 29 | 1 | PR-64 |
| **24410** | S:D950N | 2187 | 2183 | PR-45 |
|  |  | 1277 | 1275 | PR-44 |
|  |  | 121 | 1271 | PR-46 |
|  |  | 2553 | 2529 | PR-47 |
|  |  | 2498 | 2489 | PR-40 |
|  |  | 1810 | 1809 | PR-48 |
|  |  | 402 | 334 | PR-49 |
|  |  | 523 | 284 | PR-52 |
|  |  | 485 | 283 | PR-51 |
|  |  | 4058 | 227 | PR-53 |
|  |  | 1065 | 247 | PR-54 |
|  |  | 227 | 91 | PR-50 |
|  |  | 826 | 101 | PR-56 |
|  |  | 1864 | 69 | PR-55 |
|  |  | 170 | 67 | PR-41 |
|  |  | 1712 | 1691 | PR-39 |
| **24424** | S:Q954H | 700 | 303 | PR-52 |
|  |  | 512 | 203 | PR-51 |
|  |  | 4325 | 4113 | PR-53 |
|  |  | 1221 | 992 | PR-54 |
|  |  | 287 | 199 | PR-50 |
|  |  | 4444 | 4440 | PR-57 |
|  |  | 1075 | 972 | PR-56 |
|  |  | 3368 | 3301 | PR-55 |
|  |  | 1097 | 1096 | PR-60 |
|  |  | 3864 | 3861 | PR-59 |
|  |  | 739 | 739 | PR-62 |
|  |  | 584 | 584 | PR-61 |
|  |  | 1567 | 1565 | PR-58 |
|  |  | 44 | 44 | PR-66 |
|  |  | 22 | 22 | PR-74 |
|  |  | 111 | 111 | PR-65 |
|  |  | 298 | 298 | PR-63 |
|  |  | 56 | 56 | PR-64 |
| **24456** | S:Q965L | 1040 | 48 | PR-42 |
| **24461** | S:S967R | 4 | 1 | PR-44 |
| **24469** | S:N969K | 779 | 490 | PR-52 |
|  |  | 348 | 232 | PR-51 |
|  |  | 2987 | 2987 | PR-53 |
|  |  | 1037 | 1036 | PR-54 |
|  |  | 310 | 282 | PR-50 |
|  |  | 6327 | 6326 | PR-57 |
|  |  | 1141 | 1141 | PR-56 |
|  |  | 5285 | 5284 | PR-55 |
|  |  | 186 | 186 | PR-60 |
|  |  | 607 | 607 | PR-59 |
|  |  | 636 | 636 | PR-62 |
|  |  | 15 | 15 | PR-61 |
|  |  | 1736 | 1736 | PR-58 |
|  |  | 92 | 92 | PR-66 |
|  |  | 42 | 42 | PR-74 |
|  |  | 210 | 210 | PR-65 |
|  |  | 483 | 483 | PR-63 |
|  |  | 70 | 70 | PR-64 |
| **24481** | S:I973I | 2634 | 85 | PR-53 |
|  |  | 271 | 19 | PR-50 |
| **24497** | S:D979Y | 4475 | 181 | PR-57 |
| **24503** | S:L981F | 640 | 208 | PR-52 |
|  |  | 279 | 30 | PR-51 |
|  |  | 2428 | 1678 | PR-53 |
|  |  | 871 | 647 | PR-54 |
|  |  | 238 | 212 | PR-50 |
|  |  | 886 | 144 | PR-56 |
|  |  | 3675 | 347 | PR-55 |
|  |  | 135 | 18 | PR-60 |
|  |  | 395 | 134 | PR-63 |
| **24506** | S:S982P | 10 | 1 | PR-61 |
| **24550** | S:L996F | 40 | 10 | PR-39 |
| **24552** | S:I997T | 62 | 3 | PR-64 |
| **24610** | S:A1016A | 30 | 1 | PR-74 |
| **24638** | S:A1026S | 619 | 61 | PR-52 |
| **24642** | S:T1027I | 77 | 30 | PR-49 |
| **24665** | S:G1035* | 635 | 635 | PR-43 |
| **24694** | S:G1044G | 170 | 11 | PR-60 |
|  |  | 534 | 31 | PR-59 |
|  |  | 23 | 2 | PR-61 |
|  |  | 77 | 4 | PR-64 |
|  |  | 2 | 2 | PR-69 |
| **24699** | S:G1046V | 35 | 2 | PR-68 |
| **24701** | S:Y1047N | 21 | 1 | PR-40 |
| **24703** | S:Y1047* | 32 | 1 | PR-45 |
|  |  | 32 | 2 | PR-45 |
|  |  | 32 | 7 | PR-45 |
|  |  | 27 | 8 | PR-44 |
|  |  | 1 | 5 | PR-46 |
|  |  | 144 | 14 | PR-47 |
|  |  | 138 | 18 | PR-40 |
|  |  | 78 | 10 | PR-48 |
|  |  | 206 | 16 | PR-49 |
|  |  | 141 | 7 | PR-39 |
| **24739** | S:G1059G | 2665 | 159 | PR-58 |
| **24745** | S:V1061V | 219 | 19 | PR-61 |
| **24755** | S:V1065L | 9577 | 386 | PR-45 |
|  |  | 3296 | 162 | PR-51 |
| **24757** | S:V1065V | 2830 | 144 | PR-52 |
| **24763** | S:Y1067Y | 17661 | 548 | PR-48 |
| **24770** | S:A1070P | 15 | 1 | PR-66 |
| **24797** | S:P1079T | 71 | 21 | PR-41 |
| **24860** | S:T1100A | 1652 | 67 | PR-50 |
| **24902** | S:I1114V | 2262 | 147 | PR-52 |
|  |  | 3156 | 740 | PR-51 |
|  |  | 6959 | 631 | PR-54 |
| **24942** | S:D1127V | 24 | 1 | PR-65 |
| **24982** | S:P1140P | 2561 | 145 | PR-59 |
| **24991** | S:P1143P | 2965 | 146 | PR-42 |
| **25000** | S:D1146D | 2705 | 278 | PR-52 |
|  |  | 3441 | 317 | PR-51 |
|  |  | 9414 | 8549 | PR-53 |
|  |  | 7021 | 4083 | PR-54 |
|  |  | 1833 | 226 | PR-50 |
|  |  | 4238 | 3492 | PR-57 |
|  |  | 1969 | 1681 | PR-56 |
|  |  | 3422 | 3409 | PR-55 |
|  |  | 746 | 746 | PR-60 |
|  |  | 3246 | 3244 | PR-59 |
|  |  | 1463 | 1207 | PR-62 |
|  |  | 170 | 170 | PR-61 |
|  |  | 1349 | 1330 | PR-58 |
|  |  | 954 | 953 | PR-68 |
|  |  | 5 | 5 | PR-74 |
|  |  | 46 | 19 | PR-65 |
|  |  | 51 | 26 | PR-63 |
|  |  | 147 | 146 | PR-64 |
|  |  | 47 | 47 | PR-69 |
|  |  | 642 | 25 | PR-41 |
| **25003** | S:S1147S | 7147 | 433 | PR-42 |
| **25019** | S:D1153Y | 38 | 18 | PR-63 |
| **25037** | S:H1159N | 31 | 1 | PR-63 |
| **25048** | S:P1162P | 3330 | 155 | PR-44 |
| **25051** | S:D1163E | 22 | 1 | PR-63 |
| **25070** | S:S1170P | 18 | 2 | PR-44 |
| **25073** | S:G1171S | 28 | 1 | PR-65 |
| **25077** | S:I1172S | 2292 | 1 | PR-46 |
|  |  | 139 | 7 | PR-40 |
|  |  | 3 | 2 | PR-48 |
|  |  | 20 | 5 | PR-49 |
|  |  | 5 | 4 | PR-59 |
|  |  | 14 | 1 | PR-58 |
|  |  | 3 | 1 | PR-64 |
| **25079** | S:N1173D | 1 | 1 | PR-48 |
|  |  | 21 | 2 | PR-55 |
| **25084** | S:A1174A | 24 | 1 | PR-65 |
| **25094** | S:N1178Y | 26 | 1 | PR-65 |
| **25118** | S:L1186V | 20 | 1 | PR-65 |
| **25132** | S:A1190A | 21 | 1 | PR-65 |
| **25151** | S:L1197I | 14 | 1 | PR-55 |
|  |  | 23 | 1 | PR-65 |
| **25162** | S:L1200L | 15 | 14 | PR-49 |
| **25183** | S:E1207D | 24 | 1 | PR-65 |
| **25230** | S:G1223V | 288 | 70 | PR-51 |
| **25234** | S:L1224F | 290 | 121 | PR-51 |
| **25244** | S:V1228L | 3 | 1 | PR-59 |
| **25276** | S:T1238T | 479 | 121 | PR-57 |
| **25285** | S:C1241W | 32 | 1 | PR-59 |
| **25286** | S:S1242R | 64 | 3 | PR-48 |
|  | S:S1242G | 15 | 1 | PR-60 |
|  |  | 51 | 2 | PR-59 |
| **25297** | S:K1245N | 1490 | 69 | PR-40 |
| **25299** | S:G1246V | 441 | 29 | PR-51 |
| **25302** | S:C1247F | 2529 | 132 | PR-42 |
| **25305** | S:C1248F | 3073 | 196 | PR-48 |
| **25307** | S:S1249T | 1 | 1 | PR-68 |
|  |  | 2 | 2 | PR-71 |
| **25314** | S:G1251V | 470 | 78 | PR-51 |
|  |  | 27 | 5 | PR-68 |
|  |  | 93 | 6 | PR-73 |
|  |  | 17 | 17 | PR-71 |
| **25317** | S:S1252C | 550 | 80 | PR-51 |
|  |  | 33 | 5 | PR-68 |
|  |  | 92 | 6 | PR-73 |
|  |  | 19 | 19 | PR-71 |
| **25320** | S:C1253F | 4672 | 173 | PR-59 |
|  |  | 614 | 63 | PR-58 |
| **25324** | S:C1254* | 1501 | 110 | PR-42 |
|  |  | 489 | 37 | PR-43 |
|  |  | 526 | 20 | PR-52 |
|  |  | 6 | 1 | PR-74 |
|  |  | 51 | 2 | PR-65 |
| **25327** | S:K1255N | 30 | 1 | PR-68 |
| **25329** | S:F1256S | 7 | 1 | PR-73 |
| **25330** | S:F1256F | 1463 | 65 | PR-42 |
| **25331** | S:D1257Y | 3859 | 130 | PR-48 |
| **25336** | S:E1258E | 31 | 1 | PR-68 |
| **25336** | S:E1258D | 92 | 13 | PR-41 |
| **25337** | S:D1259H | 1151 | 56 | PR-42 |
|  |  | 6 | 1 | PR-74 |
|  |  | 93 | 16 | PR-41 |
|  |  | 57 | 2 | PR-72 |
| **25340** | S:D1260H | 3869 | 168 | PR-49 |
| **25344** | S:S1261Y | 1745 | 92 | PR-46 |
| **25346** | S:E1262* | 612 | 40 | PR-51 |
| **25347** | S:E1262G | 51 | 2 | PR-66 |
| **25348** | S:E1262D | 1532 | 97 | PR-44 |
| **25353** | S:V1264A | 1580 | 63 | PR-44 |
| **25362** | S:G1267V | 3622 | 132 | PR-54 |

**Supplementary Table 4:** Distribution of SARS-CoV- 2 lineages specific mutations in each sample. **S**: Sample ID; **DOC**: Date of collection; **20I**: Alpha, V1; **20H**: Beta, V2; **20J**: Gamma, V3; **21A**: Delta; **21 I**: Delta; **21B**: Kappa; **21C**: Epsilon; **21 D**: Eta; **21F**: Iota; **21G**: Lambda; **21 K21 L, 22A, 22B, 22C,22D**: Omicron

| **S** | **DOC** | **20I** | **20H** | **20J** | **21A** | **21I** | **21B** | **21C** | **21D** | **21F** | **21G** | **21K** | **21L** | **22A** | **22B** | **22C** | **22D** |
| --- | --- | --- | --- | --- | --- | --- | --- | --- | --- | --- | --- | --- | --- | --- | --- | --- | --- |
| PR-39 | 08-Nov-21 | S:  D614G, S:P681H | S:  D614G | S:  D614G, S:  H655Y | S:L452R, S:T478K, S:D614G, S:P681R, S:D950N, M:I82T, N:D63G, N:R203M, ORF3a:S26L | S:L452R, S:T478K, S:D614G, S:P681R, S:D950N, ORF1a:P1640L, ORF1a:V3718A, ORF1a:T3750I, M:I82T, N:D63G, N:R203M, ORF3a:S26L | S:L452R, S:D614G, S:P681R, N:R203M, ORF3a:S26L, ORF1a:T3646A | S:  L452R,  S:  D614G | S:  D614G, M:  I82T | S:  D614G | S:  D614G, N:  P13L | S:T478K, S:D614G, S:H655Y, S:N679K, S:P681H, S:N764K, S:D796Y, N:P13L, ORF1a:K856R | S:T478K, S:D614G, S:H655Y, S:N679K, S:P681H, S:N764K, S:D796Y, N:P13L | S:L452R, S:T478K, S:D614G, S:H655Y, S:N679K, S:P681H, S:N764K, S:D796Y, N:P13L | S:L452R, S:T478K, S:D614G, S:H655Y, S:N679K, S:P681H, S:N764K, S:D796Y, N:P13L | S:T478K, S:D614G, S:H655Y, S:N679K, S:P681H, S:N764K, S:D796Y, N:P13L | S:T478K, S:D614G, S:H655Y, S:N679K, S:P681H, S:N764K, S:D796Y, N:P13L |
| PR-40 | 15-Nov-21 | S:D614G | S:D614G | S:D614G | S:L452R, S:T478K, S:D614G, S:P681R, S:D950N, M:I82T, N:D63G, N:R203M, ORF3a:S26L | S:L452R, S:T478K, S:D614G, S:P681R, S:D950N, ORF1a:V3718A, ORF1a:T3750I, M:I82T, N:D63G, N:R203M, ORF3a:S26L | S:L452R, S:D614G, S:P681R, N:R203M, ORF3a:S26L, ORF1a:T3646A | S:L452R, S:D614G | S:D614G, M:I82T | S:D614G | S:D614G, ORF1a:P2287S, ORF1a:T3255I | S:T478K, S:D614G, ORF1a:T3255I, ORF1a:P3395H | S:T478K, S:D614G, ORF1a:T3255I, ORF1a:P3395H | S:L452R, S:T478K, S:D614G, ORF1a:T3255I, ORF1a:P3395H | S:L452R, S:T478K, S:D614G, ORF1a:T3255I, ORF1a:P3395H | S:T478K, S:D614G, ORF1a:T3255I, ORF1a:P3395H | S:T478K, S:D614G, ORF1a:T3255I, ORF1a:P3395H |
| PR-41 | 22-Nov-21 | S:D614G | S:D614G | S:D614G | S:T19R, S:L452R, S:T478K, S:D614G, S:P681R, S:D950N, M:I82T, N:D63G, N:R203M, N:D377Y, ORF3a:S26L, ORF7a:V82A, ORF7a:T120I | S:T19R, S:L452R, S:T478K, S:D614G, S:P681R, S:D950N, M:I82T, N:D63G, N:R203M, N:D377Y, ORF3a:S26L, ORF7a:V82A, ORF7a:T120I | S:L452R, S:D614G, S:P681R, N:R203M, N:D377Y, ORF3a:S26L, ORF1a:T3646A, ORF7a:V82A | S:L452R, S:D614G | S:D614G, M:I82T | S:D614G | S:D614G, ORF1a:T3255I, N:P13L | S:T478K, S:D614G, N:P13L, ORF1a:T3255I, ORF1a:P3395H | S:T478K, S:D614G, N:P13L, ORF1a:T3255I, ORF1a:P3395H | S:L452R, S:T478K, S:D614G, N:P13L, ORF1a:T3255I, ORF1a:P3395H | S:L452R, S:T478K, S:D614G, N:P13L, ORF1a:T3255I, ORF1a:P3395H | S:T478K, S:D614G, N:P13L, ORF1a:T3255I, ORF1a:P3395H | S:T478K, S:D614G, N:P13L, ORF1a:T3255I, ORF1a:P3395H |
| PR-42 | 29-Nov-21 | S:D614G | S:D614G | S:D614G | S:L452R, S:T478K, S:D614G, S:P681R, M:I82T, N:D63G, N:R203M, N:D377Y, ORF3a:S26L, ORF7a:V82A, ORF7a:T120I | S:L452R, S:T478K, S:D614G, S:P681R, ORF1a:P1640L, ORF1a:A3209V, M:I82T, N:D63G, N:R203M, N:D377Y, ORF3a:S26L, ORF7a:V82A, ORF7a:T120I | S:L452R, S:D614G, S:P681R, N:R203M, N:D377Y, ORF3a:S26L, ORF1a:T3646A, ORF7a:V82A | S:L452R, S:D614G | S:D614G, M:I82T | S:D614G | S:D614G, ORF1a:P2287S, ORF1a:T3255I | S:T478K, S:D614G, ORF1a:T3255I | S:T478K, S:D614G, ORF1a:T3255I | S:L452R, S:T478K, S:D614G, ORF1a:T3255I | S:L452R, S:T478K, S:D614G, ORF1a:T3255I | S:T478K, S:D614G, ORF1a:T3255I | S:W152R, S:T478K, S:D614G, ORF1a:T3255I |
| PR-43 | 06-Dec-21 | S:D614G | S:D614G | S:D614G | S:D614G, S:P681R, N:D63G, N:R203M | S:D614G, S:P681R, N:D63G, N:R203M | S:D614G, S:P681R, N:R203M | S:D614G | S:D614G | S:D614G | S:D614G, ORF1a:T3255I | S:D614G, ORF1a:T3255I | S:D614G, ORF1a:T3255I | S:D614G, ORF1a:T3255I | S:D614G, ORF1a:T3255I | S:D614G, ORF1a:T3255I | S:D614G, ORF1a:T3255I |
| PR-44 | 13-Dec-21 |  |  |  | S:L452R, S:T478K, S:P681R, S:D950N, M:I82T, N:D63G, ORF3a:S26L | S:L452R, S:T478K, S:P681R, S:D950N, ORF1a:P1640L, ORF1a:V3718A, ORF1a:T3750I, M:I82T, N:D63G, ORF3a:S26L | S:L452R, S:P681R, ORF3a:S26L, ORF1a:T3646A | S:L452R | M:I82T |  |  | S:T478K | S:T478K | S:L452R, S:T478K | S:L452R, S:T478K | S:T478K | S:T478K |
| PR-45 | 16-Dec-21 | S:D614G | S:D614G | S:D614G | S:T19R, S:L452R, S:T478K, S:D614G, S:P681R, S:D950N, M:I82T, N:D63G, ORF3a:S26L | S:T19R, S:L452R, S:T478K, S:D614G, S:P681R, S:D950N, ORF1a:V3718A, ORF1a:T3750I, M:I82T, N:D63G, ORF3a:S26L | S:L452R, S:D614G, S:P681R, ORF3a:S26L | S:L452R, S:D614G | S:D614G, M:I82T | S:D614G | S:D614G | S:T478K, S:D614G | S:T478K, S:D614G, N:S413R | S:L452R, S:T478K, S:D614G, N:S413R | S:L452R, S:T478K, S:D614G, N:S413R | S:T478K, S:D614G, N:S413R | S:T478K, S:D614G, N:S413R |
| PR-46 | 20-Dec-21 | S:D614G | S:D614G | S:D614G | S:L452R, S:T478K, S:D614G, S:P681R, S:D950N, M:I82T, N:D63G, ORF3a:S26L | S:L452R, S:T478K, S:D614G, S:P681R, S:D950N, ORF1a:V3718A, ORF1a:T3750I, M:I82T, N:D63G, ORF3a:S26L | S:L452R, S:D614G, S:P681R, ORF3a:S26L | S:L452R, S:D614G | S:D614G, M:I82T | S:D614G | S:D614G | S:T478K, S:D614G | S:T478K, S:D614G | S:L452R, S:T478K, S:D614G | S:L452R, S:T478K, S:D614G | S:T478K, S:D614G | S:T478K, S:D614G |
| PR-47 | 23-Dec-21 | S:D614G | S:D614G | S:D614G | S:G142D, S:L452R, S:T478K, S:D614G, S:P681R, S:D950N, M:I82T, N:D63G, ORF3a:S26L | S:G142D, S:A222V, S:L452R, S:T478K, S:D614G, S:P681R, S:D950N, ORF1a:V3718A, ORF1a:T3750I, M:I82T, N:D63G, ORF3a:S26L | S:L452R, S:D614G, S:P681R, ORF3a:S26L, ORF1a:T1567I | S:L452R, S:D614G | S:D614G, M:I82T | S:D614G | S:D614G, ORF1a:T3255I | S:T478K, S:D614G, ORF1a:A2710T, ORF1a:T3255I | S:G142D, S:T478K, S:D614G, ORF1a:T3255I | S:G142D, S:L452R, S:T478K, S:D614G, ORF1a:T3255I | S:G142D, S:L452R, S:T478K, S:D614G, ORF1a:T3255I | S:G142D, S:T478K, S:D614G, ORF1a:T3255I | S:G142D, S:T478K, S:D614G, ORF1a:T3255I |
| PR-48 | 27-Dec-21 | S:D614G | S:D614G | S:D614G | S:T19R, S:L452R, S:T478K, S:D614G, S:P681R, S:D950N, M:I82T, N:D63G, N:R203M, ORF3a:S26L | S:T19R, S:L452R, S:T478K, S:D614G, S:P681R, S:D950N, ORF1a:V3718A, ORF1a:T3750I, M:I82T, N:D63G, N:R203M, ORF3a:S26L | S:L452R, S:D614G, S:P681R, N:R203M, ORF3a:S26L, ORF1a:T3646A | S:L452R, S:D614G | S:D614G, M:I82T | S:D614G | S:D614G | S:T478K, S:D614G | S:T478K, S:D614G | S:L452R, S:T478K, S:D614G | S:L452R, S:T478K, S:D614G | S:T478K, S:D614G | S:T478K, S:D614G |
| PR-49 | 30-Dec-21 | S:D614G | S:D614G | S:D614G, S:T1027I | S:G142D, S:L452R, S:T478K, S:D614G, S:P681R, S:D950N, M:I82T, N:D63G, ORF3a:S26L, ORF7a:T120I | S:G142D, S:L452R, S:T478K, S:D614G, S:P681R, S:D950N, ORF1a:P1640L, ORF1a:V3718A, ORF1a:T3750I, M:I82T, N:D63G, ORF3a:S26L, ORF7a:T120I | S:L452R, S:D614G, S:P681R, ORF3a:S26L, ORF1a:T3646A | S:L452R, S:D614G | S:A67V, S:D614G, M:I82T | S:T95I, S:D614G | S:D614G, ORF1a:T3255I | S:A67V, S:T95I, S:G339D, S:T478K, S:T547K, S:D614G, S:D796Y, ORF1a:K856R, ORF1a:T3255I, ORF1a:P3395H | S:G142D, S:G339D, S:T478K, S:D614G, S:D796Y, ORF1a:T3255I, ORF1a:P3395H | S:G142D, S:G339D, S:L452R, S:T478K, S:D614G, S:D796Y, ORF1a:T3255I, ORF1a:P3395H | S:G142D, S:G339D, S:L452R, S:T478K, S:D614G, S:D796Y, ORF1a:T3255I, ORF1a:P3395H | S:G142D, S:G339D, S:T478K, S:D614G, S:D796Y, ORF1a:T3255I, ORF1a:P3395H | S:G142D, S:T478K, S:D614G, S:D796Y, ORF1a:T3255I, ORF1a:P3395H |
| PR-50 | 03-Jan-22 | S:D614G, S:P681H, N:R203K, N:G204R | S:K417N, S:D614G | S:D614G, S:H655Y, N:R203K, N:G204R | S:G142D, S:T478K, S:D614G, S:P681R, S:D950N, M:I82T, N:D63G | S:G142D, S:T478K, S:D614G, S:P681R, S:D950N, ORF1a:V3718A, ORF1a:T3750I, M:I82T, N:D63G | S:D614G, S:P681R | S:D614G | S:A67V, S:D614G, M:I82T | S:T95I, S:D614G | S:D614G, ORF1a:T3255I, N:P13L, N:R203K, N:G204R | S:A67V, S:T95I, S:G339D, S:S373P, S:S375F, S:K417N, S:S477N, S:T478K, S:E484A, S:T547K, S:D614G, S:H655Y, S:N679K, S:P681H, S:N764K, S:D796Y, S:N856K, S:Q954H, S:N969K, S:L981F, N:P13L, N:R203K, N:G204R, ORF1a:K856R, ORF1a:L2084I, ORF1a:A2710T, ORF1a:T3255I, ORF1a:P3395H, ORF1a:I3758V, E:T9I, M:D3G, M:Q19E, M:A63T | S:G142D, S:V213G, S:G339D, S:S371F, S:S373P, S:S375F, S:D405N, S:R408S, S:K417N, S:S477N, S:T478K, S:E484A, S:D614G, S:H655Y, S:N679K, S:P681H, S:N764K, S:D796Y, S:Q954H, S:N969K, N:P13L, N:R203K, N:G204R, ORF1a:T842I, ORF1a:G1307S, ORF1a:T3090I, ORF1a:T3255I, ORF1a:P3395H, ORF3a:T223I, E:T9I, M:Q19E, M:A63T | S:G142D, S:V213G, S:G339D, S:S371F, S:S373P, S:S375F, S:D405N, S:R408S, S:K417N, S:S477N, S:T478K, S:E484A, S:D614G, S:H655Y, S:N679K, S:P681H, S:N764K, S:D796Y, S:Q954H, S:N969K, N:P13L, N:R203K, N:G204R, ORF1a:T842I, ORF1a:G1307S, ORF1a:T3090I, ORF1a:T3255I, ORF1a:P3395H, ORF3a:T223I, E:T9I, M:Q19E, M:A63T | S:G142D, S:V213G, S:G339D, S:S371F, S:S373P, S:S375F, S:D405N, S:R408S, S:K417N, S:S477N, S:T478K, S:E484A, S:D614G, S:H655Y, S:N679K, S:P681H, S:N764K, S:D796Y, S:Q954H, S:N969K, N:P13L, N:R203K, N:G204R, ORF1a:T842I, ORF1a:G1307S, ORF1a:T3090I, ORF1a:T3255I, ORF1a:P3395H, ORF3a:T223I, E:T9I, M:Q19E, M:A63T | S:G142D, S:V213G, S:G339D, S:S371F, S:S373P, S:S375F, S:D405N, S:R408S, S:K417N, S:S477N, S:T478K, S:E484A, S:D614G, S:H655Y, S:N679K, S:P681H, S:N764K, S:D796Y, S:Q954H, S:N969K, N:P13L, N:R203K, N:G204R, ORF1a:T842I, ORF1a:G1307S, ORF1a:T3090I, ORF1a:T3255I, ORF1a:P3395H, ORF3a:T223I, E:T9I, M:Q19E, M:A63T | S:G142D, S:V213G, S:S371F, S:S373P, S:S375F, S:D405N, S:R408S, S:K417N, S:S477N, S:T478K, S:E484A, S:D614G, S:H655Y, S:N679K, S:P681H, S:N764K, S:D796Y, S:Q954H, S:N969K, N:P13L, N:R203K, N:G204R, ORF1a:T842I, ORF1a:G1307S, ORF1a:T3090I, ORF1a:T3255I, ORF1a:P3395H, ORF3a:T223I, E:T9I, M:Q19E, M:A63T |
| PR-51 | 06-Jan-22 | S:D614G, S:P681H | S:K417N, S:D614G | S:D614G, S:H655Y | S:G142D, S:L452R, S:T478K, S:D614G, S:P681R, S:D950N, M:I82T, N:D63G, ORF3a:S26L | S:G142D, S:L452R, S:T478K, S:D614G, S:P681R, S:D950N, ORF1a:V3718A, ORF1a:T3750I, M:I82T, N:D63G, ORF3a:S26L | S:L452R, S:D614G, S:P681R, ORF3a:S26L, ORF1a:T3646A | S:L452R, S:D614G | S:A67V, S:D614G, M:I82T | S:T95I, S:D614G | S:D614G, ORF1a:T3255I, N:P13L | S:A67V, S:T95I, S:G339D, S:S373P, S:S375F, S:K417N, S:T478K, S:T547K, S:D614G, S:H655Y, S:N679K, S:P681H, S:N764K, S:D796Y, S:N856K, S:Q954H, S:N969K, S:L981F, N:P13L, ORF1a:T3255I, ORF1a:P3395H, ORF1a:I3758V, E:T9I, M:Q19E, M:A63T | S:G142D, S:V213G, S:G339D, S:S371F, S:S373P, S:S375F, S:T376A, S:D405N, S:R408S, S:K417N, S:T478K, S:D614G, S:H655Y, S:N679K, S:P681H, S:N764K, S:D796Y, S:Q954H, S:N969K, N:P13L, ORF1a:T842I, ORF1a:G1307S, ORF1a:L3027F, ORF1a:T3090I, ORF1a:T3255I, ORF1a:P3395H, ORF3a:T223I, E:T9I, M:Q19E, M:A63T | S:G142D, S:V213G, S:G339D, S:S371F, S:S373P, S:S375F, S:T376A, S:D405N, S:R408S, S:K417N, S:L452R, S:T478K, S:D614G, S:H655Y, S:N679K, S:P681H, S:N764K, S:D796Y, S:Q954H, S:N969K, N:P13L, ORF1a:T842I, ORF1a:G1307S, ORF1a:L3027F, ORF1a:T3090I, ORF1a:T3255I, ORF1a:P3395H, ORF3a:T223I, E:T9I, M:Q19E, M:A63T | S:G142D, S:V213G, S:G339D, S:S371F, S:S373P, S:S375F, S:T376A, S:D405N, S:R408S, S:K417N, S:L452R, S:T478K, S:D614G, S:H655Y, S:N679K, S:P681H, S:N764K, S:D796Y, S:Q954H, S:N969K, N:P13L, ORF1a:T842I, ORF1a:G1307S, ORF1a:L3027F, ORF1a:T3090I, ORF1a:T3255I, ORF1a:P3395H, ORF3a:T223I, E:T9I, M:Q19E, M:A63T | S:G142D, S:V213G, S:G339D, S:S371F, S:S373P, S:S375F, S:T376A, S:D405N, S:R408S, S:K417N, S:T478K, S:D614G, S:H655Y, S:N679K, S:P681H, S:N764K, S:D796Y, S:Q954H, S:N969K, N:P13L, ORF1a:T842I, ORF1a:G1307S, ORF1a:L3027F, ORF1a:T3090I, ORF1a:L3201F, ORF1a:T3255I, ORF1a:P3395H, ORF3a:T223I, E:T9I, M:Q19E, M:A63T | S:G142D, S:V213G, S:S371F, S:S373P, S:S375F, S:T376A, S:D405N, S:R408S, S:K417N, S:T478K, S:D614G, S:H655Y, S:N679K, S:P681H, S:N764K, S:D796Y, S:Q954H, S:N969K, N:P13L, ORF1a:T842I, ORF1a:G1307S, ORF1a:L3027F, ORF1a:T3090I, ORF1a:L3201F, ORF1a:T3255I, ORF1a:P3395H, ORF3a:T223I, E:T9I, M:Q19E, M:A63T |
| PR-52 | 10-Jan-22 | S:D614G, S:P681H, N:R203K, N:G204R | S:K417N, S:D614G | S:D614G, S:H655Y, N:R203K, N:G204R | S:T19R, S:G142D, S:L452R, S:T478K, S:D614G, S:P681R, S:D950N, M:I82T, N:D63G, ORF3a:S26L | S:T19R, S:G142D, S:L452R, S:T478K, S:D614G, S:P681R, S:D950N, ORF1a:P1640L, ORF1a:V3718A, ORF1a:T3750I, M:I82T, N:D63G, ORF3a:S26L | S:L452R, S:D614G, S:P681R, ORF3a:S26L, ORF1a:T3646A | S:L452R, S:D614G | S:A67V, S:D614G, M:I82T | S:T95I, S:D614G | S:D614G, ORF1a:T3255I, N:P13L, N:R203K, N:G204R | S:A67V, S:T95I, S:G339D, S:S373P, S:S375F, S:K417N, S:T478K, S:Q498R, S:T547K, S:D614G, S:H655Y, S:N679K, S:P681H, S:N764K, S:D796Y, S:Q954H, S:N969K, S:L981F, N:P13L, N:R203K, N:G204R, ORF1a:K856R, ORF1a:A2710T, ORF1a:T3255I, ORF1a:P3395H, ORF1a:I3758V, E:T9I, M:Q19E, M:A63T | S:T19I, S:G142D, S:V213G, S:G339D, S:S371F, S:S373P, S:S375F, S:T376A, S:D405N, S:R408S, S:K417N, S:T478K, S:Q498R, S:D614G, S:H655Y, S:N679K, S:P681H, S:N764K, S:D796Y, S:Q954H, S:N969K, N:P13L, N:R203K, N:G204R, N:S413R, ORF1a:S135R, ORF1a:T842I, ORF1a:L3027F, ORF1a:T3090I, ORF1a:T3255I, ORF1a:P3395H, ORF3a:T223I, E:T9I, M:Q19E, M:A63T | S:T19I, S:G142D, S:V213G, S:G339D, S:S371F, S:S373P, S:S375F, S:T376A, S:D405N, S:R408S, S:K417N, S:L452R, S:T478K, S:Q498R, S:D614G, S:H655Y, S:N679K, S:P681H, S:N764K, S:D796Y, S:Q954H, S:N969K, N:P13L, N:R203K, N:G204R, N:S413R, ORF1a:S135R, ORF1a:T842I, ORF1a:L3027F, ORF1a:T3090I, ORF1a:T3255I, ORF1a:P3395H, ORF3a:T223I, E:T9I, M:Q19E, M:A63T | S:T19I, S:G142D, S:V213G, S:G339D, S:S371F, S:S373P, S:S375F, S:T376A, S:D405N, S:R408S, S:K417N, S:L452R, S:T478K, S:Q498R, S:D614G, S:H655Y, S:N679K, S:P681H, S:N764K, S:D796Y, S:Q954H, S:N969K, N:P13L, N:R203K, N:G204R, N:S413R, ORF1a:S135R, ORF1a:T842I, ORF1a:L3027F, ORF1a:T3090I, ORF1a:T3255I, ORF1a:P3395H, ORF3a:T223I, E:T9I, M:Q19E, M:A63T | S:T19I, S:G142D, S:V213G, S:G339D, S:S371F, S:S373P, S:S375F, S:T376A, S:D405N, S:R408S, S:K417N, S:T478K, S:Q498R, S:D614G, S:H655Y, S:N679K, S:P681H, S:N764K, S:D796Y, S:Q954H, S:N969K, N:P13L, N:R203K, N:G204R, N:S413R, ORF1a:S135R, ORF1a:T842I, ORF1a:L3027F, ORF1a:T3090I, ORF1a:L3201F, ORF1a:T3255I, ORF1a:P3395H, ORF3a:T223I, E:T9I, M:Q19E, M:A63T | S:T19I, S:G142D, S:V213G, S:S371F, S:S373P, S:S375F, S:T376A, S:D405N, S:R408S, S:K417N, S:T478K, S:Q498R, S:D614G, S:H655Y, S:N679K, S:P681H, S:N764K, S:D796Y, S:Q954H, S:N969K, N:P13L, N:R203K, N:G204R, N:S413R, ORF1a:S135R, ORF1a:T842I, ORF1a:L3027F, ORF1a:T3090I, ORF1a:L3201F, ORF1a:T3255I, ORF1a:P3395H, ORF3a:T223I, E:T9I, M:Q19E, M:A63T |
| PR-53 | 13-Jan-22 | S:N501Y, S:D614G, S:P681H, N:R203K, N:G204R | S:K417N, S:N501Y, S:D614G | S:N501Y, S:D614G, S:H655Y, N:R203K, N:G204R | S:G142D, S:T478K, S:D614G, S:P681R, S:D950N, M:I82T, N:D63G, ORF3a:S26L | S:G142D, S:T478K, S:D614G, S:P681R, S:D950N, ORF1a:P1640L, ORF1a:V3718A, ORF1a:T3750I, M:I82T, N:D63G, ORF3a:S26L | S:D614G, S:P681R, ORF3a:S26L | S:D614G | S:A67V, S:D614G, M:I82T | S:T95I, S:D614G | S:D614G, ORF1a:T3255I, N:P13L, N:R203K, N:G204R | S:A67V, S:T95I, S:G339D, S:S373P, S:S375F, S:K417N, S:S477N, S:T478K, S:E484A, S:Q493R, S:Q498R, S:N501Y, S:T547K, S:D614G, S:H655Y, S:N679K, S:P681H, S:N764K, S:D796Y, S:N856K, S:Q954H, S:N969K, S:L981F, N:P13L, N:R203K, N:G204R, ORF1a:K856R, ORF1a:A2710T, ORF1a:T3255I, ORF1a:P3395H, ORF1a:I3758V, E:T9I, M:D3G, M:Q19E, M:A63T | S:T19I, S:G142D, S:V213G, S:G339D, S:S371F, S:S373P, S:S375F, S:T376A, S:D405N, S:R408S, S:K417N, S:S477N, S:T478K, S:E484A, S:Q493R, S:Q498R, S:N501Y, S:D614G, S:H655Y, S:N679K, S:P681H, S:N764K, S:D796Y, S:Q954H, S:N969K, N:P13L, N:R203K, N:G204R, N:S413R, ORF1a:S135R, ORF1a:T842I, ORF1a:G1307S, ORF1a:L3027F, ORF1a:T3090I, ORF1a:T3255I, ORF1a:P3395H, ORF3a:T223I, E:T9I, M:Q19E, M:A63T | S:T19I, S:G142D, S:V213G, S:G339D, S:S371F, S:S373P, S:S375F, S:T376A, S:D405N, S:R408S, S:K417N, S:S477N, S:T478K, S:E484A, S:Q498R, S:N501Y, S:D614G, S:H655Y, S:N679K, S:P681H, S:N764K, S:D796Y, S:Q954H, S:N969K, N:P13L, N:R203K, N:G204R, N:S413R, ORF1a:S135R, ORF1a:T842I, ORF1a:G1307S, ORF1a:L3027F, ORF1a:T3090I, ORF1a:T3255I, ORF1a:P3395H, ORF3a:T223I, E:T9I, M:Q19E, M:A63T | S:T19I, S:G142D, S:V213G, S:G339D, S:S371F, S:S373P, S:S375F, S:T376A, S:D405N, S:R408S, S:K417N, S:S477N, S:T478K, S:E484A, S:Q498R, S:N501Y, S:D614G, S:H655Y, S:N679K, S:P681H, S:N764K, S:D796Y, S:Q954H, S:N969K, N:P13L, N:R203K, N:G204R, N:S413R, ORF1a:S135R, ORF1a:T842I, ORF1a:G1307S, ORF1a:L3027F, ORF1a:T3090I, ORF1a:T3255I, ORF1a:P3395H, ORF3a:T223I, E:T9I, M:Q19E, M:A63T | S:T19I, S:G142D, S:V213G, S:G339D, S:S371F, S:S373P, S:S375F, S:T376A, S:D405N, S:R408S, S:K417N, S:S477N, S:T478K, S:E484A, S:Q493R, S:Q498R, S:N501Y, S:D614G, S:H655Y, S:N679K, S:P681H, S:N764K, S:D796Y, S:Q954H, S:N969K, N:P13L, N:R203K, N:G204R, N:S413R, ORF1a:S135R, ORF1a:T842I, ORF1a:G1307S, ORF1a:L3027F, ORF1a:T3090I, ORF1a:L3201F, ORF1a:T3255I, ORF1a:P3395H, ORF3a:T223I, E:T9I, M:Q19E, M:A63T | S:T19I, S:G142D, S:V213G, S:S371F, S:S373P, S:S375F, S:T376A, S:D405N, S:R408S, S:K417N, S:S477N, S:T478K, S:E484A, S:Q498R, S:N501Y, S:D614G, S:H655Y, S:N679K, S:P681H, S:N764K, S:D796Y, S:Q954H, S:N969K, N:P13L, N:R203K, N:G204R, N:S413R, ORF1a:S135R, ORF1a:T842I, ORF1a:G1307S, ORF1a:L3027F, ORF1a:T3090I, ORF1a:L3201F, ORF1a:T3255I, ORF1a:P3395H, ORF3a:T223I, E:T9I, M:Q19E, M:A63T |
| PR-54 | 17-Jan-22 | S:N501Y, S:D614G, S:P681H, N:R203K, N:G204R | S:K417N, S:N501Y, S:D614G | S:N501Y, S:D614G, S:H655Y, N:R203K, N:G204R | S:G142D, S:L452R, S:D614G, S:P681R, S:D950N, M:I82T, N:D63G, ORF3a:S26L | S:G142D, S:L452R, S:D614G, S:P681R, S:D950N, M:I82T, N:D63G, ORF3a:S26L | S:L452R, S:D614G, S:P681R, ORF3a:S26L | S:L452R, S:D614G | S:A67V, S:D614G, M:I82T | S:T95I, S:D614G | S:D614G, ORF1a:T3255I, N:P13L, N:R203K, N:G204R | S:A67V, S:T95I, S:G339D, S:S373P, S:S375F, S:K417N, S:Q498R, S:N501Y, S:Y505H, S:T547K, S:D614G, S:H655Y, S:N679K, S:P681H, S:N764K, S:D796Y, S:N856K, S:Q954H, S:N969K, S:L981F, N:P13L, N:R203K, N:G204R, ORF1a:K856R, ORF1a:L2084I, ORF1a:A2710T, ORF1a:T3255I, ORF1a:P3395H, ORF1a:I3758V, E:T9I, M:D3G, M:Q19E, M:A63T | S:T19I, S:G142D, S:V213G, S:G339D, S:S371F, S:S373P, S:S375F, S:T376A, S:D405N, S:R408S, S:K417N, S:Q498R, S:N501Y, S:Y505H, S:D614G, S:H655Y, S:N679K, S:P681H, S:N764K, S:D796Y, S:Q954H, S:N969K, N:P13L, N:R203K, N:G204R, ORF1a:S135R, ORF1a:T842I, ORF1a:G1307S, ORF1a:L3027F, ORF1a:T3090I, ORF1a:T3255I, ORF1a:P3395H, ORF3a:T223I, E:T9I, M:Q19E, M:A63T | S:T19I, S:G142D, S:V213G, S:G339D, S:S371F, S:S373P, S:S375F, S:T376A, S:D405N, S:R408S, S:K417N, S:L452R, S:Q498R, S:N501Y, S:Y505H, S:D614G, S:H655Y, S:N679K, S:P681H, S:N764K, S:D796Y, S:Q954H, S:N969K, N:P13L, N:R203K, N:G204R, ORF1a:S135R, ORF1a:T842I, ORF1a:G1307S, ORF1a:L3027F, ORF1a:T3090I, ORF1a:T3255I, ORF1a:P3395H, ORF3a:T223I, E:T9I, M:Q19E, M:A63T | S:T19I, S:G142D, S:V213G, S:G339D, S:S371F, S:S373P, S:S375F, S:T376A, S:D405N, S:R408S, S:K417N, S:L452R, S:Q498R, S:N501Y, S:Y505H, S:D614G, S:H655Y, S:N679K, S:P681H, S:N764K, S:D796Y, S:Q954H, S:N969K, N:P13L, N:R203K, N:G204R, ORF1a:S135R, ORF1a:T842I, ORF1a:G1307S, ORF1a:L3027F, ORF1a:T3090I, ORF1a:T3255I, ORF1a:P3395H, ORF3a:T223I, E:T9I, M:Q19E, M:A63T | S:T19I, S:G142D, S:V213G, S:G339D, S:S371F, S:S373P, S:S375F, S:T376A, S:D405N, S:R408S, S:K417N, S:Q498R, S:N501Y, S:Y505H, S:D614G, S:H655Y, S:N679K, S:P681H, S:N764K, S:D796Y, S:Q954H, S:N969K, N:P13L, N:R203K, N:G204R, ORF1a:S135R, ORF1a:T842I, ORF1a:G1307S, ORF1a:L3027F, ORF1a:T3090I, ORF1a:L3201F, ORF1a:T3255I, ORF1a:P3395H, ORF3a:T223I, E:T9I, M:Q19E, M:A63T | S:T19I, S:G142D, S:V213G, S:S371F, S:S373P, S:S375F, S:T376A, S:D405N, S:R408S, S:K417N, S:Q498R, S:N501Y, S:Y505H, S:D614G, S:H655Y, S:N679K, S:P681H, S:N764K, S:D796Y, S:Q954H, S:N969K, N:P13L, N:R203K, N:G204R, ORF1a:S135R, ORF1a:T842I, ORF1a:G1307S, ORF1a:L3027F, ORF1a:T3090I, ORF1a:L3201F, ORF1a:T3255I, ORF1a:P3395H, ORF3a:T223I, E:T9I, M:Q19E, M:A63T |
| PR-55 | 20-Jan-22 | S:N501Y, S:D614G, S:P681H, N:R203K, N:G204R | S:K417N, S:N501Y, S:D614G | S:N501Y, S:D614G, S:H655Y, N:R203K, N:G204R | S:G142D, S:T478K, S:D614G, S:D950N, N:D63G | S:G142D, S:T478K, S:D614G, S:D950N, N:D63G | S:D614G | S:D614G | S:A67V, S:D614G | S:T95I, S:D614G | S:D614G, ORF1a:T3255I, N:P13L, N:R203K, N:G204R | S:A67V, S:T95I, S:G339D, S:S373P, S:S375F, S:K417N, S:S477N, S:T478K, S:E484A, S:Q493R, S:Q498R, S:N501Y, S:Y505H, S:T547K, S:D614G, S:H655Y, S:N679K, S:P681H, S:N764K, S:D796Y, S:Q954H, S:N969K, S:L981F, N:P13L, N:R203K, N:G204R, ORF1a:K856R, ORF1a:A2710T, ORF1a:T3255I, ORF1a:P3395H, E:T9I, M:Q19E, M:A63T | S:T19I, S:G142D, S:V213G, S:G339D, S:S371F, S:S373P, S:S375F, S:T376A, S:D405N, S:R408S, S:K417N, S:S477N, S:T478K, S:E484A, S:Q493R, S:Q498R, S:N501Y, S:Y505H, S:D614G, S:H655Y, S:N679K, S:P681H, S:N764K, S:D796Y, S:Q954H, S:N969K, N:P13L, N:R203K, N:G204R, N:S413R, ORF1a:S135R, ORF1a:T842I, ORF1a:G1307S, ORF1a:L3027F, ORF1a:T3090I, ORF1a:T3255I, ORF1a:P3395H, ORF3a:T223I, E:T9I, M:Q19E, M:A63T | S:T19I, S:G142D, S:V213G, S:G339D, S:S371F, S:S373P, S:S375F, S:T376A, S:D405N, S:R408S, S:K417N, S:S477N, S:T478K, S:E484A, S:Q498R, S:N501Y, S:Y505H, S:D614G, S:H655Y, S:N679K, S:P681H, S:N764K, S:D796Y, S:Q954H, S:N969K, N:P13L, N:R203K, N:G204R, N:S413R, ORF1a:S135R, ORF1a:T842I, ORF1a:G1307S, ORF1a:L3027F, ORF1a:T3090I, ORF1a:T3255I, ORF1a:P3395H, ORF3a:T223I, E:T9I, M:Q19E, M:A63T | S:T19I, S:G142D, S:V213G, S:G339D, S:S371F, S:S373P, S:S375F, S:T376A, S:D405N, S:R408S, S:K417N, S:S477N, S:T478K, S:E484A, S:Q498R, S:N501Y, S:Y505H, S:D614G, S:H655Y, S:N679K, S:P681H, S:N764K, S:D796Y, S:Q954H, S:N969K, N:P13L, N:R203K, N:G204R, N:S413R, ORF1a:S135R, ORF1a:T842I, ORF1a:G1307S, ORF1a:L3027F, ORF1a:T3090I, ORF1a:T3255I, ORF1a:P3395H, ORF3a:T223I, E:T9I, M:Q19E, M:A63T | S:T19I, S:G142D, S:V213G, S:G339D, S:S371F, S:S373P, S:S375F, S:T376A, S:D405N, S:R408S, S:K417N, S:S477N, S:T478K, S:E484A, S:Q493R, S:Q498R, S:N501Y, S:Y505H, S:D614G, S:H655Y, S:N679K, S:P681H, S:N764K, S:D796Y, S:Q954H, S:N969K, N:P13L, N:R203K, N:G204R, N:S413R, ORF1a:S135R, ORF1a:T842I, ORF1a:G1307S, ORF1a:L3027F, ORF1a:T3090I, ORF1a:L3201F, ORF1a:T3255I, ORF1a:P3395H, ORF3a:T223I, E:T9I, M:Q19E, M:A63T | S:T19I, S:G142D, S:V213G, S:S371F, S:S373P, S:S375F, S:T376A, S:D405N, S:R408S, S:K417N, S:S477N, S:T478K, S:E484A, S:Q498R, S:N501Y, S:Y505H, S:D614G, S:H655Y, S:N679K, S:P681H, S:N764K, S:D796Y, S:Q954H, S:N969K, N:P13L, N:R203K, N:G204R, N:S413R, ORF1a:S135R, ORF1a:T842I, ORF1a:G1307S, ORF1a:L3027F, ORF1a:T3090I, ORF1a:L3201F, ORF1a:T3255I, ORF1a:P3395H, ORF3a:T223I, E:T9I, M:Q19E, M:A63T |
| PR-56 | 24-Jan-22 | S:N501Y, S:D614G, S:P681H, N:R203K, N:G204R | S:K417N, S:N501Y, S:D614G | S:N501Y, S:D614G, S:H655Y, N:R203K, N:G204R | S:G142D, S:T478K, S:D614G, S:D950N | S:G142D, S:T478K, S:D614G, S:D950N | S:D614G | S:D614G | S:A67V, S:D614G | S:T95I, S:D614G | S:D614G, ORF1a:T3255I, N:P13L, N:R203K, N:G204R | S:A67V, S:T95I, S:G339D, S:S373P, S:S375F, S:K417N, S:S477N, S:T478K, S:E484A, S:Q493R, S:Q498R, S:N501Y, S:Y505H, S:T547K, S:D614G, S:H655Y, S:N679K, S:P681H, S:N764K, S:D796Y, S:N856K, S:Q954H, S:N969K, S:L981F, N:P13L, N:R203K, N:G204R, ORF1a:A2710T, ORF1a:T3255I, ORF1a:P3395H, ORF1a:I3758V, E:T9I, M:Q19E, M:A63T | S:T19I, S:G142D, S:V213G, S:G339D, S:S371F, S:S373P, S:S375F, S:T376A, S:D405N, S:R408S, S:K417N, S:S477N, S:T478K, S:E484A, S:Q493R, S:Q498R, S:N501Y, S:Y505H, S:D614G, S:H655Y, S:N679K, S:P681H, S:N764K, S:D796Y, S:Q954H, S:N969K, N:P13L, N:R203K, N:G204R, N:S413R, ORF1a:S135R, ORF1a:T842I, ORF1a:G1307S, ORF1a:L3027F, ORF1a:T3090I, ORF1a:T3255I, ORF1a:P3395H, ORF3a:T223I, E:T9I, M:Q19E, M:A63T | S:T19I, S:G142D, S:V213G, S:G339D, S:S371F, S:S373P, S:S375F, S:T376A, S:D405N, S:R408S, S:K417N, S:S477N, S:T478K, S:E484A, S:Q498R, S:N501Y, S:Y505H, S:D614G, S:H655Y, S:N679K, S:P681H, S:N764K, S:D796Y, S:Q954H, S:N969K, N:P13L, N:P151S, N:R203K, N:G204R, N:S413R, ORF1a:S135R, ORF1a:T842I, ORF1a:G1307S, ORF1a:L3027F, ORF1a:T3090I, ORF1a:T3255I, ORF1a:P3395H, ORF3a:T223I, E:T9I, M:Q19E, M:A63T | S:T19I, S:G142D, S:V213G, S:G339D, S:S371F, S:S373P, S:S375F, S:T376A, S:D405N, S:R408S, S:K417N, S:S477N, S:T478K, S:E484A, S:Q498R, S:N501Y, S:Y505H, S:D614G, S:H655Y, S:N679K, S:P681H, S:N764K, S:D796Y, S:Q954H, S:N969K, N:P13L, N:R203K, N:G204R, N:S413R, ORF1a:S135R, ORF1a:T842I, ORF1a:G1307S, ORF1a:L3027F, ORF1a:T3090I, ORF1a:T3255I, ORF1a:P3395H, ORF3a:T223I, E:T9I, M:Q19E, M:A63T | S:T19I, S:G142D, S:V213G, S:G339D, S:S371F, S:S373P, S:S375F, S:T376A, S:D405N, S:R408S, S:K417N, S:S477N, S:T478K, S:E484A, S:Q493R, S:Q498R, S:N501Y, S:Y505H, S:D614G, S:H655Y, S:N679K, S:P681H, S:N764K, S:D796Y, S:Q954H, S:N969K, N:P13L, N:R203K, N:G204R, N:S413R, ORF1a:S135R, ORF1a:T842I, ORF1a:G1307S, ORF1a:L3027F, ORF1a:T3090I, ORF1a:L3201F, ORF1a:T3255I, ORF1a:P3395H, ORF3a:T223I, E:T9I, M:Q19E, M:A63T | S:T19I, S:G142D, S:V213G, S:S371F, S:S373P, S:S375F, S:T376A, S:D405N, S:R408S, S:K417N, S:S477N, S:T478K, S:E484A, S:Q498R, S:N501Y, S:Y505H, S:D614G, S:H655Y, S:N679K, S:P681H, S:N764K, S:D796Y, S:Q954H, S:N969K, N:P13L, N:R203K, N:G204R, N:S413R, ORF1a:S135R, ORF1a:T842I, ORF1a:G1307S, ORF1a:L3027F, ORF1a:T3090I, ORF1a:L3201F, ORF1a:T3255I, ORF1a:P3395H, ORF3a:T223I, E:T9I, M:Q19E, M:A63T |
| PR-57 | 28-Jan-22 | S:N501Y, S:D614G, S:P681H, N:R203K, N:G204R | S:K417N, S:N501Y, S:D614G | S:N501Y, S:D614G, S:H655Y, N:R203K, N:G204R | S:G142D, S:T478K, S:D614G | S:G142D, S:T478K, S:D614G | S:D614G | S:D614G | S:D614G | S:D614G | S:D614G, ORF1a:T3255I, N:P13L, N:R203K, N:G204R | S:G339D, S:S373P, S:S375F, S:K417N, S:S477N, S:T478K, S:E484A, S:Q493R, S:Q498R, S:N501Y, S:Y505H, S:T547K, S:D614G, S:H655Y, S:N679K, S:P681H, S:N764K, S:D796Y, S:Q954H, S:N969K, N:P13L, N:R203K, N:G204R, ORF1a:T3255I, ORF1a:P3395H, E:T9I, M:Q19E, M:A63T | S:T19I, S:G142D, S:V213G, S:G339D, S:S371F, S:S373P, S:S375F, S:T376A, S:D405N, S:R408S, S:K417N, S:S477N, S:T478K, S:E484A, S:Q493R, S:Q498R, S:N501Y, S:Y505H, S:D614G, S:H655Y, S:N679K, S:P681H, S:N764K, S:D796Y, S:Q954H, S:N969K, N:P13L, N:R203K, N:G204R, N:S413R, ORF1a:S135R, ORF1a:T842I, ORF1a:G1307S, ORF1a:L3027F, ORF1a:T3090I, ORF1a:T3255I, ORF1a:P3395H, ORF3a:T223I, E:T9I, M:Q19E, M:A63T | S:T19I, S:G142D, S:V213G, S:G339D, S:S371F, S:S373P, S:S375F, S:T376A, S:D405N, S:R408S, S:K417N, S:S477N, S:T478K, S:E484A, S:Q498R, S:N501Y, S:Y505H, S:D614G, S:H655Y, S:N679K, S:P681H, S:N764K, S:D796Y, S:Q954H, S:N969K, N:P13L, N:P151S, N:R203K, N:G204R, N:S413R, ORF1a:S135R, ORF1a:T842I, ORF1a:G1307S, ORF1a:L3027F, ORF1a:T3090I, ORF1a:T3255I, ORF1a:P3395H, ORF3a:T223I, E:T9I, M:Q19E, M:A63T | S:T19I, S:G142D, S:V213G, S:G339D, S:S371F, S:S373P, S:S375F, S:T376A, S:D405N, S:R408S, S:K417N, S:S477N, S:T478K, S:E484A, S:Q498R, S:N501Y, S:Y505H, S:D614G, S:H655Y, S:N679K, S:P681H, S:N764K, S:D796Y, S:Q954H, S:N969K, N:P13L, N:R203K, N:G204R, N:S413R, ORF1a:S135R, ORF1a:T842I, ORF1a:G1307S, ORF1a:L3027F, ORF1a:T3090I, ORF1a:T3255I, ORF1a:P3395H, ORF3a:T223I, E:T9I, M:Q19E, M:A63T | S:T19I, S:G142D, S:V213G, S:G339D, S:S371F, S:S373P, S:S375F, S:T376A, S:D405N, S:R408S, S:K417N, S:S477N, S:T478K, S:E484A, S:Q493R, S:Q498R, S:N501Y, S:Y505H, S:D614G, S:H655Y, S:N679K, S:P681H, S:N764K, S:D796Y, S:Q954H, S:N969K, N:P13L, N:R203K, N:G204R, N:S413R, ORF1a:S135R, ORF1a:T842I, ORF1a:G1307S, ORF1a:L3027F, ORF1a:T3090I, ORF1a:L3201F, ORF1a:T3255I, ORF1a:P3395H, ORF3a:T223I, E:T9I, M:Q19E, M:A63T | S:T19I, S:G142D, S:V213G, S:S371F, S:S373P, S:S375F, S:T376A, S:D405N, S:R408S, S:K417N, S:S477N, S:T478K, S:E484A, S:Q498R, S:N501Y, S:Y505H, S:D614G, S:H655Y, S:N679K, S:P681H, S:N764K, S:D796Y, S:Q954H, S:N969K, N:P13L, N:R203K, N:G204R, N:S413R, ORF1a:S135R, ORF1a:T842I, ORF1a:G1307S, ORF1a:L3027F, ORF1a:T3090I, ORF1a:L3201F, ORF1a:T3255I, ORF1a:P3395H, ORF3a:T223I, E:T9I, M:Q19E, M:A63T |
| PR-58 | 31-Jan-22 | S:N501Y, S:D614G, S:P681H, N:R203K, N:G204R | S:K417N, S:N501Y, S:D614G | S:N501Y, S:D614G, S:H655Y, N:R203K, N:G204R | S:G142D, S:D614G | S:G142D, S:D614G | S:D614G | S:D614G | S:D614G | S:D614G | S:D614G, ORF1a:T3255I, N:P13L, N:R203K, N:G204R | S:G339D, S:S373P, S:S375F, S:K417N, S:Q493R, S:Q498R, S:N501Y, S:Y505H, S:T547K, S:D614G, S:H655Y, S:N679K, S:P681H, S:N764K, S:D796Y, S:Q954H, S:N969K, N:P13L, N:R203K, N:G204R, ORF1a:A2710T, ORF1a:T3255I, ORF1a:P3395H, E:T9I, M:Q19E, M:A63T | S:T19I, S:G142D, S:V213G, S:G339D, S:S371F, S:S373P, S:S375F, S:T376A, S:D405N, S:R408S, S:K417N, S:Q493R, S:Q498R, S:N501Y, S:Y505H, S:D614G, S:H655Y, S:N679K, S:P681H, S:N764K, S:D796Y, S:Q954H, S:N969K, N:P13L, N:R203K, N:G204R, N:S413R, ORF1a:S135R, ORF1a:T842I, ORF1a:G1307S, ORF1a:L3027F, ORF1a:T3090I, ORF1a:T3255I, ORF1a:P3395H, ORF3a:T223I, E:T9I, M:Q19E, M:A63T | S:T19I, S:G142D, S:V213G, S:G339D, S:S371F, S:S373P, S:S375F, S:T376A, S:D405N, S:R408S, S:K417N, S:Q498R, S:N501Y, S:Y505H, S:D614G, S:H655Y, S:N679K, S:P681H, S:N764K, S:D796Y, S:Q954H, S:N969K, N:P13L, N:R203K, N:G204R, N:S413R, ORF1a:S135R, ORF1a:T842I, ORF1a:G1307S, ORF1a:L3027F, ORF1a:T3090I, ORF1a:T3255I, ORF1a:P3395H, ORF3a:T223I, E:T9I, M:Q19E, M:A63T | S:T19I, S:G142D, S:V213G, S:G339D, S:S371F, S:S373P, S:S375F, S:T376A, S:D405N, S:R408S, S:K417N, S:Q498R, S:N501Y, S:Y505H, S:D614G, S:H655Y, S:N679K, S:P681H, S:N764K, S:D796Y, S:Q954H, S:N969K, N:P13L, N:R203K, N:G204R, N:S413R, ORF1a:S135R, ORF1a:T842I, ORF1a:G1307S, ORF1a:L3027F, ORF1a:T3090I, ORF1a:T3255I, ORF1a:P3395H, ORF3a:T223I, E:T9I, M:Q19E, M:A63T | S:T19I, S:G142D, S:V213G, S:G339D, S:S371F, S:S373P, S:S375F, S:T376A, S:D405N, S:R408S, S:K417N, S:Q493R, S:Q498R, S:N501Y, S:Y505H, S:D614G, S:H655Y, S:N679K, S:P681H, S:N764K, S:D796Y, S:Q954H, S:N969K, N:P13L, N:R203K, N:G204R, N:S413R, ORF1a:S135R, ORF1a:T842I, ORF1a:G1307S, ORF1a:L3027F, ORF1a:T3090I, ORF1a:L3201F, ORF1a:T3255I, ORF1a:P3395H, ORF3a:T223I, E:T9I, M:Q19E, M:A63T | S:T19I, S:G142D, S:V213G, S:S371F, S:S373P, S:S375F, S:T376A, S:D405N, S:R408S, S:K417N, S:Q498R, S:N501Y, S:Y505H, S:D614G, S:H655Y, S:N679K, S:P681H, S:N764K, S:D796Y, S:Q954H, S:N969K, N:P13L, N:R203K, N:G204R, N:S413R, ORF1a:S135R, ORF1a:T842I, ORF1a:G1307S, ORF1a:L3027F, ORF1a:T3090I, ORF1a:L3201F, ORF1a:T3255I, ORF1a:P3395H, ORF3a:T223I, E:T9I, M:Q19E, M:A63T |
| PR-59 | 03-Feb-22 | S:N501Y, S:D614G, S:P681H, N:R203K, N:G204R | S:K417N, S:N501Y, S:D614G | S:N501Y, S:D614G, S:H655Y, N:R203K, N:G204R | S:G142D, S:T478K, S:D614G | S:G142D, S:T478K, S:D614G | S:D614G | S:D614G | S:D614G | S:L5F, S:D614G | S:D614G, ORF1a:T3255I, N:P13L, N:R203K, N:G204R | S:G339D, S:S373P, S:S375F, S:K417N, S:N440K, S:S477N, S:T478K, S:E484A, S:Q493R, S:Q498R, S:N501Y, S:Y505H, S:T547K, S:D614G, S:H655Y, S:N679K, S:P681H, S:N764K, S:D796Y, S:Q954H, S:N969K, N:P13L, N:R203K, N:G204R, ORF1a:T3255I, ORF1a:P3395H, E:T9I, M:Q19E, M:A63T | S:T19I, S:G142D, S:V213G, S:G339D, S:S371F, S:S373P, S:S375F, S:T376A, S:D405N, S:R408S, S:K417N, S:N440K, S:S477N, S:T478K, S:E484A, S:Q493R, S:Q498R, S:N501Y, S:Y505H, S:D614G, S:H655Y, S:N679K, S:P681H, S:N764K, S:D796Y, S:Q954H, S:N969K, N:P13L, N:R203K, N:G204R, N:S413R, ORF1a:S135R, ORF1a:T842I, ORF1a:G1307S, ORF1a:L3027F, ORF1a:T3090I, ORF1a:T3255I, ORF1a:P3395H, ORF3a:T223I, E:T9I, M:Q19E, M:A63T | S:T19I, S:G142D, S:V213G, S:G339D, S:S371F, S:S373P, S:S375F, S:T376A, S:D405N, S:R408S, S:K417N, S:N440K, S:S477N, S:T478K, S:E484A, S:Q498R, S:N501Y, S:Y505H, S:D614G, S:H655Y, S:N679K, S:P681H, S:N764K, S:D796Y, S:Q954H, S:N969K, N:P13L, N:R203K, N:G204R, N:S413R, ORF1a:S135R, ORF1a:T842I, ORF1a:G1307S, ORF1a:L3027F, ORF1a:T3090I, ORF1a:T3255I, ORF1a:P3395H, ORF3a:T223I, E:T9I, M:Q19E, M:A63T | S:T19I, S:G142D, S:V213G, S:G339D, S:S371F, S:S373P, S:S375F, S:T376A, S:D405N, S:R408S, S:K417N, S:N440K, S:S477N, S:T478K, S:E484A, S:Q498R, S:N501Y, S:Y505H, S:D614G, S:H655Y, S:N679K, S:P681H, S:N764K, S:D796Y, S:Q954H, S:N969K, N:P13L, N:R203K, N:G204R, N:S413R, ORF1a:S135R, ORF1a:T842I, ORF1a:G1307S, ORF1a:L3027F, ORF1a:T3090I, ORF1a:T3255I, ORF1a:P3395H, ORF3a:T223I, E:T9I, M:Q19E, M:A63T | S:T19I, S:G142D, S:V213G, S:G339D, S:S371F, S:S373P, S:S375F, S:T376A, S:D405N, S:R408S, S:K417N, S:N440K, S:S477N, S:T478K, S:E484A, S:Q493R, S:Q498R, S:N501Y, S:Y505H, S:D614G, S:H655Y, S:N679K, S:P681H, S:N764K, S:D796Y, S:Q954H, S:N969K, N:P13L, N:R203K, N:G204R, N:S413R, ORF1a:S135R, ORF1a:T842I, ORF1a:G1307S, ORF1a:L3027F, ORF1a:T3090I, ORF1a:L3201F, ORF1a:T3255I, ORF1a:P3395H, ORF3a:T223I, E:T9I, M:Q19E, M:A63T | S:T19I, S:G142D, S:V213G, S:S371F, S:S373P, S:S375F, S:T376A, S:D405N, S:R408S, S:K417N, S:N440K, S:S477N, S:T478K, S:E484A, S:Q498R, S:N501Y, S:Y505H, S:D614G, S:H655Y, S:N679K, S:P681H, S:N764K, S:D796Y, S:Q954H, S:N969K, N:P13L, N:R203K, N:G204R, N:S413R, ORF1a:S135R, ORF1a:T842I, ORF1a:G1307S, ORF1a:L3027F, ORF1a:T3090I, ORF1a:L3201F, ORF1a:T3255I, ORF1a:P3395H, ORF3a:T223I, E:T9I, M:Q19E, M:A63T |
| PR-60 | 07-Feb-22 | S:D614G, S:P681H, N:R203K, N:G204R | S:K417N, S:D614G | S:D614G, S:H655Y, ORF1a:S1188L, N:R203K, N:G204R | S:G142D, S:D614G | S:G142D, S:D614G | S:D614G | S:D614G | S:D614G | S:D614G | S:D614G, ORF1a:T3255I, N:P13L, N:R203K, N:G204R | S:G339D, S:S373P, S:S375F, S:K417N, S:D614G, S:H655Y, S:N679K, S:P681H, S:N764K, S:D796Y, S:Q954H, S:N969K, S:L981F, N:P13L, N:R203K, N:G204R, ORF1a:A2710T, ORF1a:T3255I, ORF1a:P3395H, E:T9I, M:Q19E, M:A63T | S:G142D, S:V213G, S:G339D, S:S371F, S:S373P, S:S375F, S:T376A, S:D405N, S:R408S, S:K417N, S:D614G, S:H655Y, S:N679K, S:P681H, S:N764K, S:D796Y, S:Q954H, S:N969K, N:P13L, N:R203K, N:G204R, ORF1a:S135R, ORF1a:T842I, ORF1a:G1307S, ORF1a:L3027F, ORF1a:T3090I, ORF1a:T3255I, ORF1a:P3395H, ORF3a:T223I, E:T9I, M:Q19E, M:A63T | S:G142D, S:V213G, S:G339D, S:S371F, S:S373P, S:S375F, S:T376A, S:D405N, S:R408S, S:K417N, S:D614G, S:H655Y, S:N679K, S:P681H, S:N764K, S:D796Y, S:Q954H, S:N969K, N:P13L, N:R203K, N:G204R, ORF1a:S135R, ORF1a:T842I, ORF1a:G1307S, ORF1a:L3027F, ORF1a:T3090I, ORF1a:T3255I, ORF1a:P3395H, ORF3a:T223I, E:T9I, M:Q19E, M:A63T | S:G142D, S:V213G, S:G339D, S:S371F, S:S373P, S:S375F, S:T376A, S:D405N, S:R408S, S:K417N, S:D614G, S:H655Y, S:N679K, S:P681H, S:N764K, S:D796Y, S:Q954H, S:N969K, N:P13L, N:R203K, N:G204R, ORF1a:S135R, ORF1a:T842I, ORF1a:G1307S, ORF1a:L3027F, ORF1a:T3090I, ORF1a:T3255I, ORF1a:P3395H, ORF3a:T223I, E:T9I, M:Q19E, M:A63T | S:G142D, S:V213G, S:G339D, S:S371F, S:S373P, S:S375F, S:T376A, S:D405N, S:R408S, S:K417N, S:D614G, S:H655Y, S:N679K, S:P681H, S:N764K, S:D796Y, S:Q954H, S:N969K, N:P13L, N:R203K, N:G204R, ORF1a:S135R, ORF1a:T842I, ORF1a:G1307S, ORF1a:L3027F, ORF1a:T3090I, ORF1a:L3201F, ORF1a:T3255I, ORF1a:P3395H, ORF3a:T223I, E:T9I, M:Q19E, M:A63T | S:G142D, S:V213G, S:S371F, S:S373P, S:S375F, S:T376A, S:D405N, S:R408S, S:K417N, S:D614G, S:H655Y, S:N679K, S:P681H, S:N764K, S:D796Y, S:Q954H, S:N969K, N:P13L, N:R203K, N:G204R, ORF1a:S135R, ORF1a:T842I, ORF1a:G1307S, ORF1a:L3027F, ORF1a:T3090I, ORF1a:L3201F, ORF1a:T3255I, ORF1a:P3395H, ORF3a:T223I, E:T9I, M:Q19E, M:A63T |
| PR-61 | 10-Feb-22 | S:D614G, S:P681H, N:R203K, N:G204R | S:K417N, S:D614G | S:D614G, S:H655Y, ORF1a:S1188L, N:R203K, N:G204R | S:G142D, S:D614G | S:G142D, S:D614G | S:D614G | S:D614G | S:D614G | S:L5F, S:D614G | S:D614G, ORF1a:T3255I, N:P13L, N:R203K, N:G204R | S:G339D, S:S373P, S:S375F, S:K417N, S:D614G, S:H655Y, S:N679K, S:P681H, S:N764K, S:D796Y, S:Q954H, S:N969K, N:P13L, N:R203K, N:G204R, ORF1a:T3255I, ORF1a:P3395H, E:T9I, M:Q19E, M:A63T | S:T19I, S:G142D, S:V213G, S:G339D, S:S371F, S:S373P, S:S375F, S:T376A, S:D405N, S:R408S, S:K417N, S:D614G, S:H655Y, S:N679K, S:P681H, S:N764K, S:D796Y, S:Q954H, S:N969K, N:P13L, N:R203K, N:G204R, ORF1a:S135R, ORF1a:T842I, ORF1a:G1307S, ORF1a:L3027F, ORF1a:T3090I, ORF1a:T3255I, ORF1a:P3395H, ORF3a:T223I, E:T9I, M:Q19E, M:A63T | S:T19I, S:G142D, S:V213G, S:G339D, S:S371F, S:S373P, S:S375F, S:T376A, S:D405N, S:R408S, S:K417N, S:D614G, S:H655Y, S:N679K, S:P681H, S:N764K, S:D796Y, S:Q954H, S:N969K, N:P13L, N:R203K, N:G204R, ORF1a:S135R, ORF1a:T842I, ORF1a:G1307S, ORF1a:L3027F, ORF1a:T3090I, ORF1a:T3255I, ORF1a:P3395H, ORF3a:T223I, E:T9I, M:Q19E, M:A63T | S:T19I, S:G142D, S:V213G, S:G339D, S:S371F, S:S373P, S:S375F, S:T376A, S:D405N, S:R408S, S:K417N, S:D614G, S:H655Y, S:N679K, S:P681H, S:N764K, S:D796Y, S:Q954H, S:N969K, N:P13L, N:R203K, N:G204R, ORF1a:S135R, ORF1a:T842I, ORF1a:G1307S, ORF1a:L3027F, ORF1a:T3090I, ORF1a:T3255I, ORF1a:P3395H, ORF3a:T223I, E:T9I, M:Q19E, M:A63T | S:T19I, S:G142D, S:V213G, S:G339D, S:S371F, S:S373P, S:S375F, S:T376A, S:D405N, S:R408S, S:K417N, S:D614G, S:H655Y, S:N679K, S:P681H, S:N764K, S:D796Y, S:Q954H, S:N969K, N:P13L, N:R203K, N:G204R, ORF1a:S135R, ORF1a:T842I, ORF1a:G1307S, ORF1a:L3027F, ORF1a:T3090I, ORF1a:L3201F, ORF1a:T3255I, ORF1a:P3395H, ORF3a:T223I, E:T9I, M:Q19E, M:A63T | S:T19I, S:G142D, S:V213G, S:S371F, S:S373P, S:S375F, S:T376A, S:D405N, S:R408S, S:K417N, S:D614G, S:H655Y, S:N679K, S:P681H, S:N764K, S:D796Y, S:Q954H, S:N969K, N:P13L, N:R203K, N:G204R, ORF1a:S135R, ORF1a:T842I, ORF1a:G1307S, ORF1a:L3027F, ORF1a:T3090I, ORF1a:L3201F, ORF1a:T3255I, ORF1a:P3395H, ORF3a:T223I, E:T9I, M:Q19E, M:A63T |
| PR-62 | 14-Feb-22 | S:N501Y, S:D614G, S:P681H, N:R203K, N:G204R | S:K417N, S:N501Y, S:D614G | S:N501Y, S:D614G, S:H655Y, N:R203K, N:G204R | S:G142D, S:T478K, S:D614G | S:G142D, S:T478K, S:D614G | S:D614G | S:D614G | S:D614G | S:D614G | S:D614G, ORF1a:T3255I, N:P13L, N:R203K, N:G204R | S:G339D, S:S373P, S:S375F, S:K417N, S:S477N, S:T478K, S:E484A, S:Q493R, S:Q498R, S:N501Y, S:Y505H, S:T547K, S:D614G, S:H655Y, S:N679K, S:P681H, S:N764K, S:D796Y, S:Q954H, S:N969K, N:P13L, N:R203K, N:G204R, ORF1a:T3255I, ORF1a:P3395H, E:T9I, M:Q19E, M:A63T | S:T19I, S:G142D, S:V213G, S:G339D, S:S371F, S:S373P, S:S375F, S:T376A, S:D405N, S:R408S, S:K417N, S:S477N, S:T478K, S:E484A, S:Q493R, S:Q498R, S:N501Y, S:Y505H, S:D614G, S:H655Y, S:N679K, S:P681H, S:N764K, S:D796Y, S:Q954H, S:N969K, N:P13L, N:R203K, N:G204R, N:S413R, ORF1a:S135R, ORF1a:T842I, ORF1a:G1307S, ORF1a:L3027F, ORF1a:T3090I, ORF1a:T3255I, ORF1a:P3395H, ORF3a:T223I, E:T9I, M:Q19E, M:A63T | S:T19I, S:G142D, S:V213G, S:G339D, S:S371F, S:S373P, S:S375F, S:T376A, S:D405N, S:R408S, S:K417N, S:S477N, S:T478K, S:E484A, S:Q498R, S:N501Y, S:Y505H, S:D614G, S:H655Y, S:N679K, S:P681H, S:N764K, S:D796Y, S:Q954H, S:N969K, N:P13L, N:R203K, N:G204R, N:S413R, ORF1a:S135R, ORF1a:T842I, ORF1a:G1307S, ORF1a:L3027F, ORF1a:T3090I, ORF1a:T3255I, ORF1a:P3395H, ORF3a:T223I, E:T9I, M:Q19E, M:A63T | S:T19I, S:G142D, S:V213G, S:G339D, S:S371F, S:S373P, S:S375F, S:T376A, S:D405N, S:R408S, S:K417N, S:S477N, S:T478K, S:E484A, S:Q498R, S:N501Y, S:Y505H, S:D614G, S:H655Y, S:N679K, S:P681H, S:N764K, S:D796Y, S:Q954H, S:N969K, N:P13L, N:R203K, N:G204R, N:S413R, ORF1a:S135R, ORF1a:T842I, ORF1a:G1307S, ORF1a:L3027F, ORF1a:T3090I, ORF1a:T3255I, ORF1a:P3395H, ORF3a:T223I, E:T9I, M:Q19E, M:A63T | S:T19I, S:G142D, S:V213G, S:G339D, S:S371F, S:S373P, S:S375F, S:T376A, S:D405N, S:R408S, S:K417N, S:S477N, S:T478K, S:E484A, S:Q493R, S:Q498R, S:N501Y, S:Y505H, S:D614G, S:H655Y, S:N679K, S:P681H, S:N764K, S:D796Y, S:Q954H, S:N969K, N:P13L, N:R203K, N:G204R, N:S413R, ORF1a:S135R, ORF1a:T842I, ORF1a:G1307S, ORF1a:L3027F, ORF1a:T3090I, ORF1a:L3201F, ORF1a:T3255I, ORF1a:P3395H, ORF3a:T223I, E:T9I, M:Q19E, M:A63T | S:T19I, S:G142D, S:V213G, S:S371F, S:S373P, S:S375F, S:T376A, S:D405N, S:R408S, S:K417N, S:S477N, S:T478K, S:E484A, S:Q498R, S:N501Y, S:Y505H, S:D614G, S:H655Y, S:N679K, S:P681H, S:N764K, S:D796Y, S:Q954H, S:N969K, N:P13L, N:R203K, N:G204R, N:S413R, ORF1a:S135R, ORF1a:T842I, ORF1a:G1307S, ORF1a:L3027F, ORF1a:T3090I, ORF1a:L3201F, ORF1a:T3255I, ORF1a:P3395H, ORF3a:T223I, E:T9I, M:Q19E, M:A63T |
| PR-63 | 18-Feb-22 | S:D614G | S:K417N, S:D614G | S:D614G | S:G142D, S:D614G | S:G142D, S:D614G | S:D614G | S:D614G | S:D614G | S:D614G | S:D614G, ORF1a:T3255I, N:P13L | S:G339D, S:S373P, S:S375F, S:K417N, S:D614G, S:N764K, S:D796Y, S:Q954H, S:N969K, S:L981F, N:P13L, ORF1a:T3255I, ORF1a:P3395H, E:T9I, M:Q19E, M:A63T | S:G142D, S:G339D, S:S371F, S:S373P, S:S375F, S:T376A, S:D405N, S:R408S, S:K417N, S:D614G, S:N764K, S:D796Y, S:Q954H, S:N969K, N:P13L, ORF1a:S135R, ORF1a:L3027F, ORF1a:T3090I, ORF1a:T3255I, ORF1a:P3395H, ORF3a:T223I, E:T9I, M:Q19E, M:A63T | S:G142D, S:G339D, S:S371F, S:S373P, S:S375F, S:T376A, S:D405N, S:R408S, S:K417N, S:D614G, S:N764K, S:D796Y, S:Q954H, S:N969K, N:P13L, ORF1a:S135R, ORF1a:L3027F, ORF1a:T3090I, ORF1a:T3255I, ORF1a:P3395H, ORF3a:T223I, E:T9I, M:Q19E, M:A63T | S:G142D, S:G339D, S:S371F, S:S373P, S:S375F, S:T376A, S:D405N, S:R408S, S:K417N, S:D614G, S:N764K, S:D796Y, S:Q954H, S:N969K, N:P13L, ORF1a:S135R, ORF1a:L3027F, ORF1a:T3090I, ORF1a:T3255I, ORF1a:P3395H, ORF3a:T223I, E:T9I, M:Q19E, M:A63T | S:G142D, S:G339D, S:S371F, S:S373P, S:S375F, S:T376A, S:D405N, S:R408S, S:K417N, S:D614G, S:N764K, S:D796Y, S:Q954H, S:N969K, N:P13L, ORF1a:S135R, ORF1a:L3027F, ORF1a:T3090I, ORF1a:L3201F, ORF1a:T3255I, ORF1a:P3395H, ORF3a:T223I, E:T9I, M:Q19E, M:A63T | S:G142D, S:S371F, S:S373P, S:S375F, S:T376A, S:D405N, S:R408S, S:K417N, S:D614G, S:N764K, S:D796Y, S:Q954H, S:N969K, N:P13L, ORF1a:S135R, ORF1a:L3027F, ORF1a:T3090I, ORF1a:L3201F, ORF1a:T3255I, ORF1a:P3395H, ORF3a:T223I, E:T9I, M:Q19E, M:A63T |
| PR-64 | 21-Feb-22 | S:D614G, S:P681H, N:R203K, N:G204R | S:K417N, S:D614G | S:D614G, S:H655Y, N:R203K, N:G204R | S:G142D, S:D614G | S:G142D, S:D614G | S:D614G | S:D614G | S:D614G | S:D614G | S:D614G, ORF1a:T3255I, N:P13L, N:R203K, N:G204R | S:G339D, S:S373P, S:S375F, S:K417N, S:D614G, S:H655Y, S:N679K, S:P681H, S:N764K, S:D796Y, S:Q954H, S:N969K, N:P13L, N:R203K, N:G204R, ORF1a:T3255I, ORF1a:P3395H, E:T9I, M:Q19E, M:A63T | S:T19I, S:G142D, S:V213G, S:G339D, S:S371F, S:S373P, S:S375F, S:T376A, S:D405N, S:R408S, S:K417N, S:D614G, S:H655Y, S:N679K, S:P681H, S:N764K, S:D796Y, S:Q954H, S:N969K, N:P13L, N:R203K, N:G204R, ORF1a:S135R, ORF1a:T842I, ORF1a:G1307S, ORF1a:L3027F, ORF1a:T3090I, ORF1a:T3255I, ORF1a:P3395H, ORF3a:T223I, E:T9I, M:Q19E, M:A63T | S:T19I, S:G142D, S:V213G, S:G339D, S:S371F, S:S373P, S:S375F, S:T376A, S:D405N, S:R408S, S:K417N, S:D614G, S:H655Y, S:N679K, S:P681H, S:N764K, S:D796Y, S:Q954H, S:N969K, N:P13L, N:R203K, N:G204R, ORF1a:S135R, ORF1a:T842I, ORF1a:G1307S, ORF1a:L3027F, ORF1a:T3090I, ORF1a:T3255I, ORF1a:P3395H, ORF3a:T223I, E:T9I, M:Q19E, M:A63T | S:T19I, S:G142D, S:V213G, S:G339D, S:S371F, S:S373P, S:S375F, S:T376A, S:D405N, S:R408S, S:K417N, S:D614G, S:H655Y, S:N679K, S:P681H, S:N764K, S:D796Y, S:Q954H, S:N969K, N:P13L, N:R203K, N:G204R, ORF1a:S135R, ORF1a:T842I, ORF1a:G1307S, ORF1a:L3027F, ORF1a:T3090I, ORF1a:T3255I, ORF1a:P3395H, ORF3a:T223I, E:T9I, M:Q19E, M:A63T | S:T19I, S:G142D, S:V213G, S:G339D, S:S371F, S:S373P, S:S375F, S:T376A, S:D405N, S:R408S, S:K417N, S:D614G, S:H655Y, S:N679K, S:P681H, S:N764K, S:D796Y, S:Q954H, S:N969K, N:P13L, N:R203K, N:G204R, ORF1a:S135R, ORF1a:T842I, ORF1a:G1307S, ORF1a:L3027F, ORF1a:T3090I, ORF1a:L3201F, ORF1a:T3255I, ORF1a:P3395H, ORF3a:T223I, E:T9I, M:Q19E, M:A63T | S:T19I, S:G142D, S:V213G, S:S371F, S:S373P, S:S375F, S:T376A, S:D405N, S:R408S, S:K417N, S:D614G, S:H655Y, S:N679K, S:P681H, S:N764K, S:D796Y, S:Q954H, S:N969K, N:P13L, N:R203K, N:G204R, ORF1a:S135R, ORF1a:T842I, ORF1a:G1307S, ORF1a:L3027F, ORF1a:T3090I, ORF1a:L3201F, ORF1a:T3255I, ORF1a:P3395H, ORF3a:T223I, E:T9I, M:Q19E, M:A63T |
| PR-65 | 25-Feb-22 | S:D614G | S:K417N, S:D614G | S:D614G | S:G142D, S:D614G | S:G142D, S:D614G | S:D614G | S:D614G | S:D614G | S:D614G | S:D614G, ORF1a:T3255I, N:P13L | S:G339D, S:S373P, S:S375F, S:K417N, S:T547K, S:D614G, S:N764K, S:D796Y, S:Q954H, S:N969K, N:P13L, ORF1a:L2084I, ORF1a:T3255I, E:T9I, M:Q19E, M:A63T | S:T19I, S:G142D, S:G339D, S:S371F, S:S373P, S:S375F, S:T376A, S:D405N, S:R408S, S:K417N, S:D614G, S:N764K, S:D796Y, S:Q954H, S:N969K, N:P13L, ORF1a:S135R, ORF1a:L3027F, ORF1a:T3090I, ORF1a:T3255I, ORF3a:T223I, E:T9I, M:Q19E, M:A63T | S:T19I, S:G142D, S:G339D, S:S371F, S:S373P, S:S375F, S:T376A, S:D405N, S:R408S, S:K417N, S:D614G, S:N764K, S:D796Y, S:Q954H, S:N969K, N:P13L, ORF1a:S135R, ORF1a:L3027F, ORF1a:T3090I, ORF1a:T3255I, ORF3a:T223I, E:T9I, M:Q19E, M:A63T | S:T19I, S:G142D, S:G339D, S:S371F, S:S373P, S:S375F, S:T376A, S:D405N, S:R408S, S:K417N, S:D614G, S:N764K, S:D796Y, S:Q954H, S:N969K, N:P13L, ORF1a:S135R, ORF1a:L3027F, ORF1a:T3090I, ORF1a:T3255I, ORF3a:T223I, E:T9I, M:Q19E, M:A63T | S:T19I, S:G142D, S:G339D, S:S371F, S:S373P, S:S375F, S:T376A, S:D405N, S:R408S, S:K417N, S:D614G, S:N764K, S:D796Y, S:Q954H, S:N969K, N:P13L, ORF1a:S135R, ORF1a:L3027F, ORF1a:T3090I, ORF1a:L3201F, ORF1a:T3255I, ORF3a:T223I, E:T9I, M:Q19E, M:A63T | S:T19I, S:G142D, S:S371F, S:S373P, S:S375F, S:T376A, S:D405N, S:R408S, S:K417N, S:D614G, S:N764K, S:D796Y, S:Q954H, S:N969K, N:P13L, ORF1a:S135R, ORF1a:L3027F, ORF1a:T3090I, ORF1a:L3201F, ORF1a:T3255I, ORF3a:T223I, E:T9I, M:Q19E, M:A63T |
| PR-66 | 28-Feb-22 |  |  |  |  |  |  |  |  |  |  | S:G339D, S:Q954H, S:N969K, E:T9I, M:Q19E, M:A63T | S:G339D, S:Q954H, S:N969K, ORF1a:S135R, ORF3a:T223I, E:T9I, M:Q19E, M:A63T | S:G339D, S:Q954H, S:N969K, ORF1a:S135R, ORF3a:T223I, E:T9I, M:Q19E, M:A63T | S:G339D, S:Q954H, S:N969K, ORF1a:S135R, ORF3a:T223I, E:T9I, M:Q19E, M:A63T | S:G339D, S:Q954H, S:N969K, ORF1a:S135R, ORF3a:T223I, E:T9I, M:Q19E, M:A63T | S:Q954H, S:N969K, ORF1a:S135R, ORF3a:T223I, E:T9I, M:Q19E, M:A63T |
| PR-67 | 03-Mar-22 | S:P681H, N:R203K, N:G204R |  | S:H655Y, N:R203K, N:G204R | S:G142D | S:G142D |  |  |  |  | ORF1a:T3255I, N:P13L, N:R203K, N:G204R | S:H655Y, S:N679K, S:P681H, S:N764K, S:D796Y, N:P13L, N:R203K, N:G204R, ORF1a:T3255I, ORF1a:P3395H, E:T9I, M:Q19E, M:A63T | S:G142D, S:H655Y, S:N679K, S:P681H, S:N764K, S:D796Y, N:P13L, N:R203K, N:G204R, ORF1a:S135R, ORF1a:T3090I, ORF1a:T3255I, ORF1a:P3395H, ORF3a:T223I, E:T9I, M:Q19E, M:A63T | S:G142D, S:H655Y, S:N679K, S:P681H, S:N764K, S:D796Y, N:P13L, N:R203K, N:G204R, ORF1a:S135R, ORF1a:T3090I, ORF1a:T3255I, ORF1a:P3395H, ORF3a:T223I, E:T9I, M:Q19E, M:A63T | S:G142D, S:H655Y, S:N679K, S:P681H, S:N764K, S:D796Y, N:P13L, N:R203K, N:G204R, ORF1a:S135R, ORF1a:T3090I, ORF1a:T3255I, ORF1a:P3395H, ORF3a:T223I, E:T9I, M:Q19E, M:A63T | S:G142D, S:H655Y, S:N679K, S:P681H, S:N764K, S:D796Y, N:P13L, N:R203K, N:G204R, ORF1a:S135R, ORF1a:T3090I, ORF1a:L3201F, ORF1a:T3255I, ORF1a:P3395H, ORF3a:T223I, E:T9I, M:Q19E, M:A63T | S:G142D, S:H655Y, S:N679K, S:P681H, S:N764K, S:D796Y, N:P13L, N:R203K, N:G204R, ORF1a:S135R, ORF1a:T3090I, ORF1a:L3201F, ORF1a:T3255I, ORF1a:P3395H, ORF3a:T223I, E:T9I, M:Q19E, M:A63T |
| PR-68 | 07-Mar-22 |  |  |  |  |  |  |  |  |  |  | M:Q19E, M:A63T | M:Q19E, M:A63T | M:Q19E, M:A63T | M:Q19E, M:A63T | M:Q19E, M:A63T | M:Q19E, M:A63T |
| PR-69 | 10-Mar-22 | S:D614G, S:P681H | S:K417N, S:D614G | S:D614G, S:H655Y | S:G142D, S:D614G | S:G142D, S:D614G | S:D614G | S:D614G | S:D614G | S:T95I, S:D614G | S:D614G, N:P13L | S:T95I, S:G339D, S:S373P, S:S375F, S:K417N, S:T547K, S:D614G, S:H655Y, S:N679K, S:P681H, N:P13L, E:T9I | S:G142D, S:G339D, S:S371F, S:S373P, S:S375F, S:T376A, S:D405N, S:R408S, S:K417N, S:D614G, S:H655Y, S:N679K, S:P681H, N:P13L, N:S413R, ORF1a:T3090I, E:T9I | S:G142D, S:G339D, S:S371F, S:S373P, S:S375F, S:T376A, S:D405N, S:R408S, S:K417N, S:D614G, S:H655Y, S:N679K, S:P681H, N:P13L, N:S413R, ORF1a:T3090I, E:T9I | S:G142D, S:G339D, S:S371F, S:S373P, S:S375F, S:T376A, S:D405N, S:R408S, S:K417N, S:D614G, S:H655Y, S:N679K, S:P681H, N:P13L, N:S413R, ORF1a:T3090I, E:T9I | S:G142D, S:G339D, S:S371F, S:S373P, S:S375F, S:T376A, S:D405N, S:R408S, S:K417N, S:D614G, S:H655Y, S:N679K, S:P681H, N:P13L, N:S413R, ORF1a:T3090I, E:T9I | S:G142D, S:S371F, S:S373P, S:S375F, S:T376A, S:D405N, S:R408S, S:K417N, S:D614G, S:H655Y, S:N679K, S:P681H, N:P13L, N:S413R, ORF1a:T3090I, E:T9I |
| PR-70 | 14-Mar-22 | S:D614G | S:K417N, S:D614G | S:D614G | S:D614G | S:D614G | S:D614G | S:D614G | S:D614G | S:D614G | S:D614G, N:P13L | S:G339D, S:S373P, S:S375F, S:K417N, S:D614G, N:P13L | S:G339D, S:S371F, S:S373P, S:S375F, S:T376A, S:D405N, S:R408S, S:K417N, S:D614G, N:P13L, ORF1a:G1307S, ORF1a:T3090I | S:G339D, S:S371F, S:S373P, S:S375F, S:T376A, S:D405N, S:R408S, S:K417N, S:D614G, N:P13L, ORF1a:G1307S, ORF1a:T3090I | S:G339D, S:S371F, S:S373P, S:S375F, S:T376A, S:D405N, S:R408S, S:K417N, S:D614G, N:P13L, ORF1a:G1307S, ORF1a:T3090I | S:G339D, S:S371F, S:S373P, S:S375F, S:T376A, S:D405N, S:R408S, S:K417N, S:D614G, N:P13L, ORF1a:G1307S, ORF1a:T3090I | S:S371F, S:S373P, S:S375F, S:T376A, S:D405N, S:R408S, S:K417N, S:D614G, N:P13L, ORF1a:G1307S, ORF1a:T3090I |
| PR-71 | 21-Mar-22 |  |  |  |  |  |  |  |  |  |  | E:T9I | E:T9I | E:T9I | E:T9I | E:T9I | E:T9I |
| PR-72 | 28-Mar-22 | S:P681H | S:K417N | S:H655Y |  |  |  |  |  |  | N:P13L | S:G339D, S:S373P, S:S375F, S:K417N, S:H655Y, S:N679K, S:P681H, N:P13L | S:G339D, S:S371F, S:S373P, S:S375F, S:T376A, S:D405N, S:R408S, S:K417N, S:H655Y, S:N679K, S:P681H, N:P13L, ORF1a:L3027F, ORF1a:T3090I | S:G339D, S:S371F, S:S373P, S:S375F, S:T376A, S:D405N, S:R408S, S:K417N, S:H655Y, S:N679K, S:P681H, N:P13L, ORF1a:L3027F, ORF1a:T3090I | S:G339D, S:S371F, S:S373P, S:S375F, S:T376A, S:D405N, S:R408S, S:K417N, S:H655Y, S:N679K, S:P681H, N:P13L, ORF1a:L3027F, ORF1a:T3090I | S:G339D, S:S371F, S:S373P, S:S375F, S:T376A, S:D405N, S:R408S, S:K417N, S:H655Y, S:N679K, S:P681H, N:P13L, ORF1a:L3027F, ORF1a:T3090I | S:S371F, S:S373P, S:S375F, S:T376A, S:D405N, S:R408S, S:K417N, S:H655Y, S:N679K, S:P681H, N:P13L, ORF1a:L3027F, ORF1a:T3090I |
| PR-73 | 04-Apr-22 |  |  |  |  |  |  |  | S:A67V | S:T95I | N:P13L | S:A67V, S:T95I, N:P13L, ORF1a:L2084I, M:Q19E, M:A63T | N:P13L, M:Q19E, M:A63T | N:P13L, M:Q19E, M:A63T | N:P13L, M:Q19E, M:A63T | N:P13L, M:Q19E, M:A63T | N:P13L, M:Q19E, M:A63T |
| PR-74 | 11-Apr-22 | S:D614G | S:D614G | S:D614G | S:D614G | S:D614G | S:D614G | S:D614G | S:D614G | S:D614G | S:D614G | S:D614G, S:Q954H, S:N969K | S:D614G, S:Q954H, S:N969K, ORF1a:S135R | S:D614G, S:Q954H, S:N969K, ORF1a:S135R | S:D614G, S:Q954H, S:N969K, ORF1a:S135R | S:D614G, S:Q954H, S:N969K, ORF1a:S135R | S:D614G, S:Q954H, S:N969K, ORF1a:S135R |
| PR-75 | 18-Apr-22 | S:P681H |  | S:H655Y |  |  |  |  |  |  |  | S:H655Y, S:N679K, S:P681H, E:T9I, M:Q19E, M:A63T | S:H655Y, S:N679K, S:P681H, ORF3a:T223I, E:T9I, M:Q19E, M:A63T | S:H655Y, S:N679K, S:P681H, ORF3a:T223I, E:T9I, M:Q19E, M:A63T | S:H655Y, S:N679K, S:P681H, ORF3a:T223I, E:T9I, M:Q19E, M:A63T | S:H655Y, S:N679K, S:P681H, ORF3a:T223I, E:T9I, M:Q19E, M:A63T | S:H655Y, S:N679K, S:P681H, ORF3a:T223I, E:T9I, M:Q19E, M:A63T |
| PR-76 | 25-Apr-22 | S:P681H | S:K417N | S:H655Y |  |  |  |  |  |  | N:P13L | S:G339D, S:S373P, S:S375F, S:K417N, S:H655Y, S:N679K, S:P681H, S:Q954H, N:P13L, M:Q19E, M:A63T | S:G339D, S:S371F, S:S373P, S:S375F, S:T376A, S:D405N, S:R408S, S:K417N, S:H655Y, S:N679K, S:P681H, S:Q954H, N:P13L, N:S413R, M:Q19E, M:A63T | S:G339D, S:S371F, S:S373P, S:S375F, S:T376A, S:D405N, S:R408S, S:K417N, S:H655Y, S:N679K, S:P681H, S:Q954H, N:P13L, N:S413R, M:Q19E, M:A63T | S:G339D, S:S371F, S:S373P, S:S375F, S:T376A, S:D405N, S:R408S, S:K417N, S:H655Y, S:N679K, S:P681H, S:Q954H, N:P13L, N:S413R, M:Q19E, M:A63T | S:G339D, S:S371F, S:S373P, S:S375F, S:T376A, S:D405N, S:R408S, S:K417N, S:H655Y, S:N679K, S:P681H, S:Q954H, N:P13L, N:S413R, M:Q19E, M:A63T | S:S371F, S:S373P, S:S375F, S:T376A, S:D405N, S:R408S, S:K417N, S:H655Y, S:N679K, S:P681H, S:Q954H, N:P13L, N:S413R, M:Q19E, M:A63T |

Number of mutations

| SARS-CoV-2 virus genome | Synonymous | Non-synonymous | Deletions |
| --- | --- | --- | --- |
| 5'UTR | 0 | 0 | 0 |
| 3'UTR | 0 | 0 | 3 |
| ORF 1a | 201 | 740 | 51 |
| ORF 1ab | 115 | 404 | 22 |
| Spike(S) | 55 | 264 | 22 |
| ORF3a | 21 | 67 | 6 |
| Intergenic region | 0 | 0 | 2 |
| E | 5 | 14 | 0 |
| M | 9 | 39 | 3 |
| ORF6 | 5 | 5 | 0 |
| ORF7a | 0 | 19 | 1 |
| ORF7b | 2 | 13 | 1 |
| ORF8 | 5 | 12 | 0 |
| N | 26 | 75 | 3 |
| ORF10 | 4 | 4 | 0 |

**Supplementary Figure 1:** Overall mutation profile of SARS-CoV-2 genomes from sewage water samples when compared to the SARS-CoV-2 reference genome (MN908947.3).

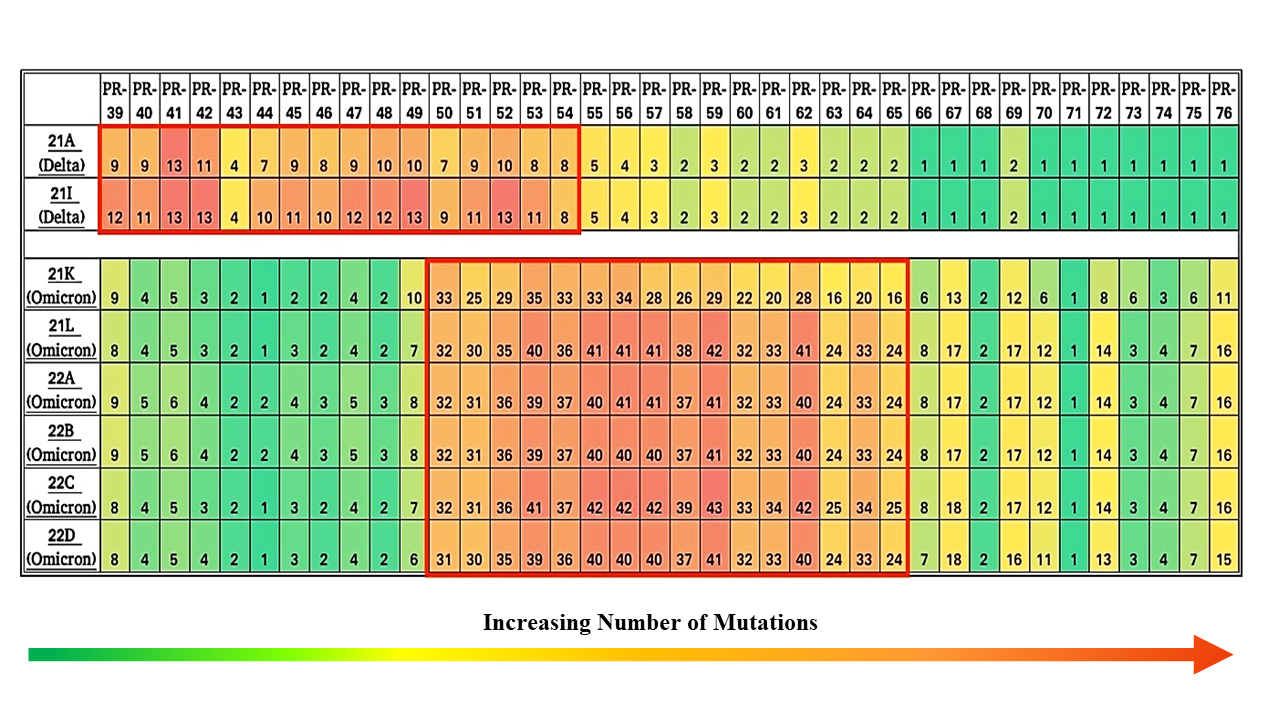

**Supplementary Figure 2:** Amino acid mutation count of omicron and delta VOC per sample

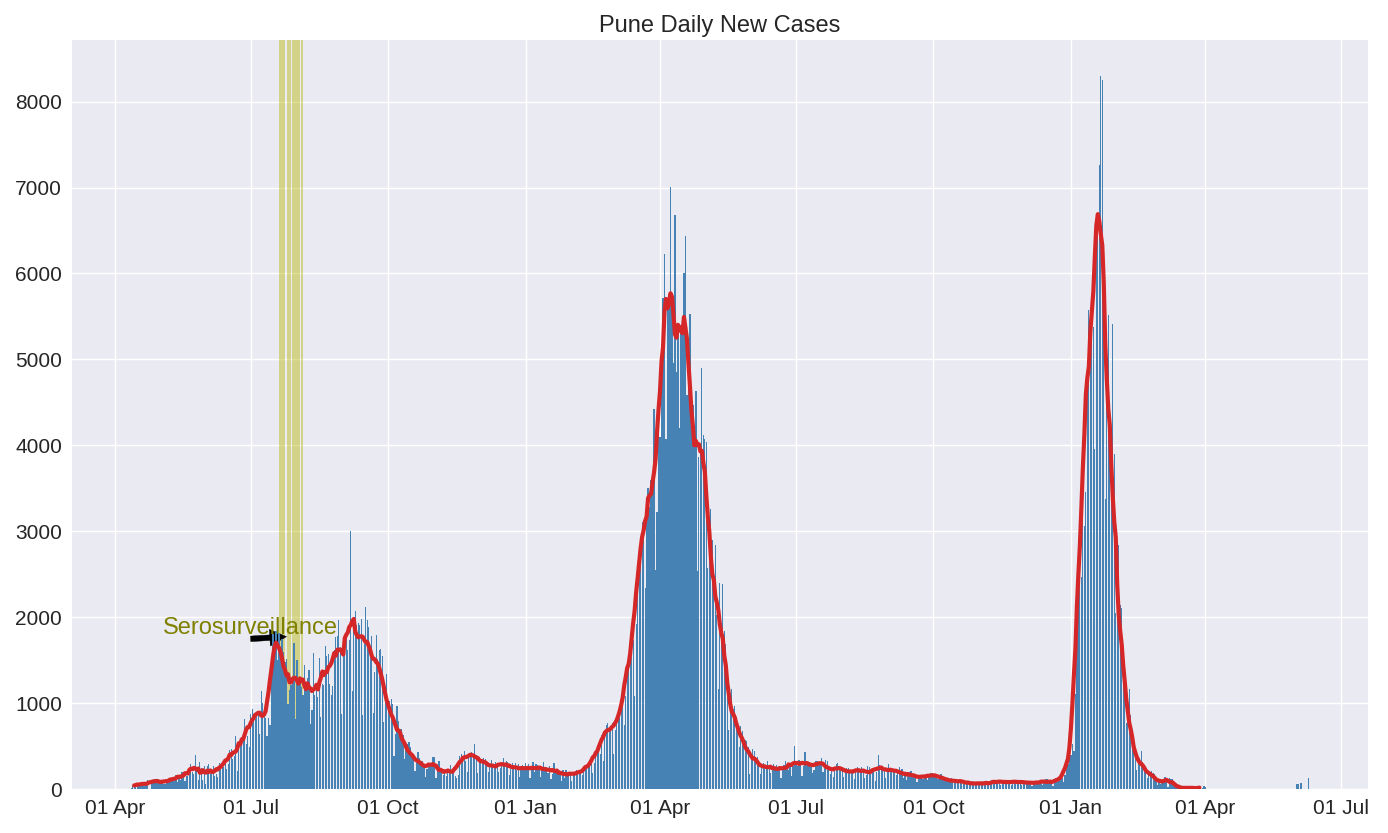

**Supplementary Figure 3:** Daily new COVID-19 positive case in Pune City from April, 2020 to July, 2022 (Note: This image has been obtained from <http://cms.unipune.ac.in/~bspujari/Covid19/Pune2/>)
